# An Updated Systematic Review and Meta-Analysis of Mortality, Need for ICU admission, Use of Mechanical Ventilation, Adverse effects and other Clinical Outcomes of Ivermectin Treatment in COVID-19 Patients

**DOI:** 10.1101/2021.04.30.21256415

**Authors:** Smruti Karale, Vikas Bansal, Janaki Makadia, Muhammad Tayyeb, Hira Khan, Shree Spandana Ghanta, Romil Singh, Aysun Tekin, Abhishek Bhurwal, Hemant Mutneja, Ishita Mehra, Rahul Kashyap

## Abstract

**Importance:** Repurposing Ivermectin, a known anti-parasitic agent, for treating COVID-19 has demonstrated positive results in several studies. We aim to evaluate the benefit and risk of Ivermectin in COVID-19.

**Methods:** We conducted a systematic search for full-text manuscripts published from February 1, 2020, to August 15th, 2021 focusing on Ivermectin therapy against COVID-19. The primary outcomes were mortality, need for intensive care unit (ICU) admission; secondary outcomes were - adverse effects, need for mechanical ventilation, viral clearance, time to viral clearance, need for hospitalization, and length of hospital stay. Random-effects models were used for all analyses.

**Results:** We included a total of 52 studies (n=17561) in the qualitative analysis, out of these, 44 studies (n=14019) were included in the meta-analysis. In the mortality meta-analysis (N=29), odds of death were lower in the Ivermectin-arm compared to control (OR 0.54, p=0.009). Although lower odds of mortality were observed in various subgroup analyses of RCTs, they did not reach statistical significance: therapeutic RCTs: mild-moderate COVID-19 (OR 0.31, p=0.06), therapeutic RCTs: severe/critical COVID-19 (OR 0.86, p=0.56), inpatient RCTs: mild-moderate COVID-19 (OR 0.18, p=0.08), inpatient RCTs: severe/critical COVID-19 (OR 0.86, p=0.56). Ivermectin, mostly as adjuvant therapy, was associated with higher odds of viral clearance (N=22) (OR 3.52, p=0.0002), shorter duration to achieve viral clearance (N=8) (MD - 4.12, p=0.02), reduced need for hospitalization (N=6) (OR 0.34, p=008).

**Conclusion:** Our meta-analysis suggests that the mortality benefit of Ivermectin in COVID-19 is uncertain. But as adjuvant therapy, Ivermectin may improve viral clearance and reduce the need for hospitalization.

**Highlights:** *What We Already Know about This Topic:*

1. COVID-19 is an ongoing global pandemic, for which Ivermectin has been tried on a therapeutic and prophylactic basis.
2. Results from several clinical trials and observational studies suggest that Ivermectin may improve survival and clinical outcomes with a good safety profile when compared with other treatments; however, the current evidence is limited..

*What This Article Tells Us That Is New:*

1. This systematic review and meta-analysis provide a summary of the latest literature on the efficacy and safety of Ivermectin use for COVID-19.
2. Based on our analysis of the latest evidence, we found that Ivermectin’s benefit in reducing mortality cannot be concluded with confidence. However, as an adjuvant therapy it may help reduce the need for hospitalization, duration for viral clearance while increasing the likelihood of achieving viral clearance.
3. We need more high-quality data for conclusive evidence regarding the benefit of Ivermectin in reducing the need for ICU admissions, mechanical ventilation and duration of hospital stay in COVID-19 patients.

## Introduction

On March 11, 2020, the World Health Organization (WHO) declared COVID-19 disease a global pandemic^1^. Today, on September 9,2021, there are about >222 million confirmed cases worldwide and >4.6 million deaths due to COVID-19^2^. Patients infected with SARS-COV-2, the causative Coronavirus, exhibit a spectrum of clinical presentations ranging from asymptomatic to severely critical and multiple risk factors are involved in the prognosis of the disease^3-8^. To guide decision-making for this multi-system disease^8-17^, a variety of treatment modalities have been proposed and evaluated^18-23^. However, the evidence for the risk-benefit ratio of most of these treatments remains unclear.

Ivermectin has held an excellent safety record in humans as an antiparasitic agent for over three decades^24^. Apart from its established activity against several parasites, Ivermectin has demonstrated antiviral activity against many RNA and DNA viruses in vivo and against a few in vitro by targeting specific proteins^25^. One such plausible mechanism against RNA viruses is the inhibition of importin (IMP) α/β Integrase, thus blocking viral entry into the nucleus and the ensuing suppression of the cell’s anti-viral response^25-28^. Moreover, it exhibited anti-bacterial, anti-inflammatory, and anti-cancer effects^24,29,30^. An Australian study demonstrated the effectiveness of Ivermectin in inhibiting SARS-COV2 in-vitro^31^. Subsequently, several Ivermectin-based clinical trials and observational studies conducted globally showed mostly positive results^32-35^. Some countries have incorporated this therapy in their COVID-19 guidelines^36-39^. Only a month after this authorization, Peru witnessed a record drop of 25% in COVID-19 related mortality^40^.

Dr. Satoshi Ōmura, who was among the scientists that won the Nobel prize for the discovery of Ivermectin, highlighted the need to develop a drug that arrests the early stage of viral replication in COVID-19^41^. Repurposing Ivermectin, a low-cost drug may help bypass the time and funds needed to develop and test novel therapies. The lack of robust evidence has been a major hindrance in the large-scale approval of Ivermectin. The available studies are methodologically diverse and mostly under-powered while few report conflicting results. Therefore, we intend to systematically review the latest literature and plan to perform a meta-analysis to overcome some of the individual study biases.

## Methods

### Search Method and strategy

We conducted a comprehensive literature search for studies mentioning the use of Ivermectin in COVID-19 from February 1, 2020 till August 15, 2021 (Figure 1). We screened all titles and abstracts identified by preliminary search for eligible studies and manually searched references of included articles for additional studies. Then we analyzed full-text manuscripts of included studies according to the protocol of Systematic Review and Meta-Analysis (PRISMA) guidelines.

**Figure 1:**
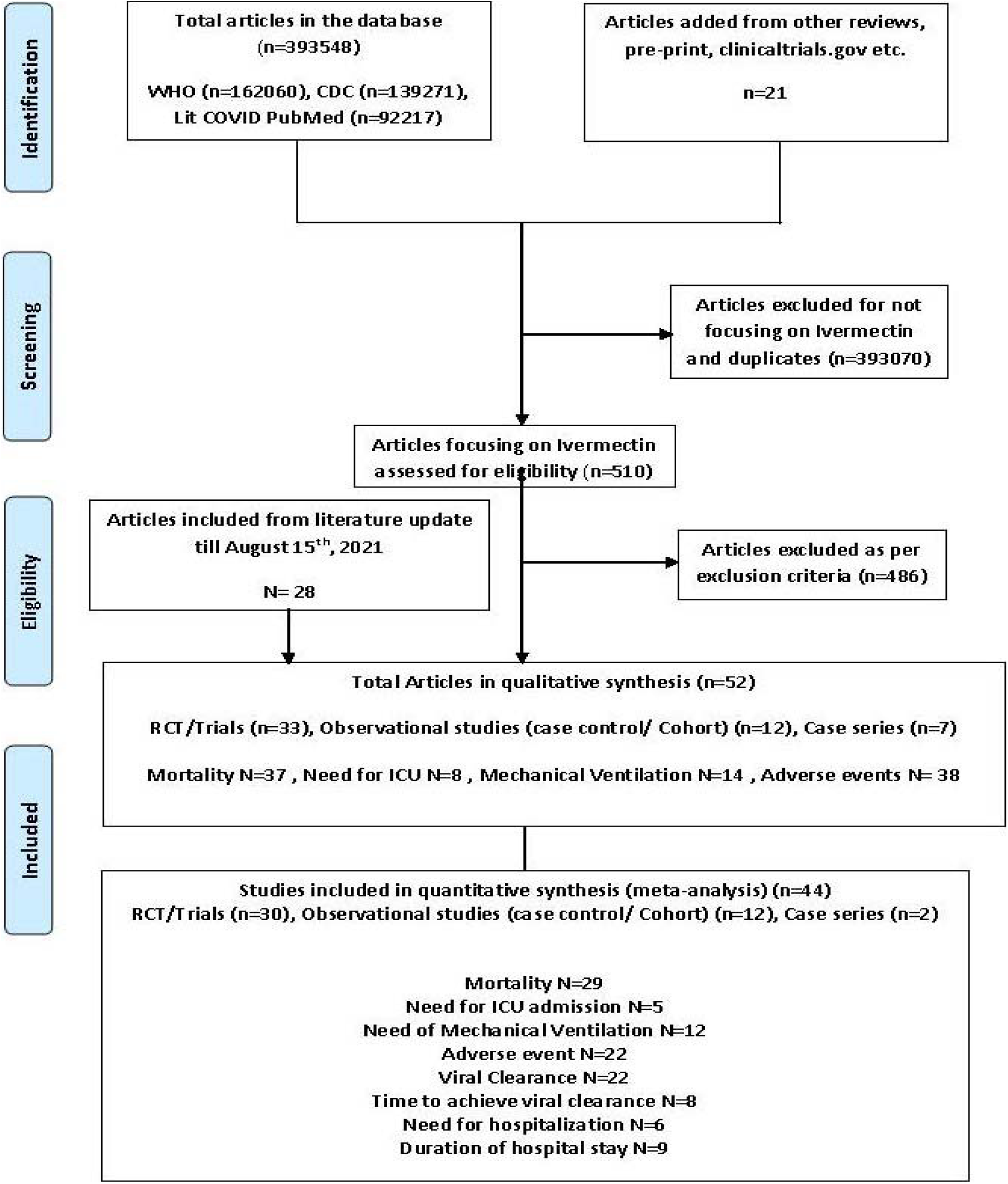
Preferred Reporting Items for Systematic Reviews and Meta-Analyses flow— study inclusion. ICU: Intensive Care Unit, CDC: Centers for Disease Control and Prevention, RCT: Randomized controlled trial, WHO: World Health Organization.

### Eligibility Criteria

We included studies reporting outcomes of mortality, ICU admission, mechanical ventilation, adverse-effects, viral clearance, hospitalization with therapeutic Ivermectin treatment against COVID-19. We excluded studies focusing on pregnant females, in-vitro studies, meta-analyses, case reports and case series <5 patients.

### Study selection and data extraction

The extracted data was tabularized in Microsoft Excel with following parameters: author, country of study, study design, number of patients, Ivermectin regimen, concomitant treatment, efficacy outcomes and adverse effects. The included data was checked for accuracy by all authors and disagreements were resolved through consensus and after input from a third reviewer. IRB approval was exempted because data was extracted from publicly available studies. We also analyzed raw data files or supplemental files of studies by Cadegiani^42^ and Lima-Morales et al^43^ for outcomes of cases receiving Ivermectin, Niaee et al^44^ for clinical severity based mortality data and duration of hospital stay, Choudhary et al^35^ for time for viral clearance, Mahmud et al^45^ for need for hospitalization, Ghooi et al^46^ for mortality in Ivermectin receiving patients, Samaha et al^47^ for viral clearance. In Cadegiani et al’s study^42^, we considered ‘CG1’ as control group because only this sample was demographically comparable to the treatment group. Additionally, we compiled all ongoing clinical trials on Ivermectin from clinicaltrials.gov (e-Table-8). We excluded the study by Elgazzar et al^48^ because it was retracted.

### Outcomes

The primary outcome was defined as mortality benefit with Ivermectin therapy in COVID-19 and need for ICU admission. The secondary outcomes were need for mechanical ventilation, adverse effects of Ivermectin, viral clearance and time to achieve viral clearance, need for hospitalization and duration of hospital stay.

### Statistical analysis

Primary and Secondary outcomes were quantitatively analyzed by Review Manager (RevMan) Version 5.4 for windows^49^ and Comprehensive Meta-Analysis software package (Biostat, Englewood, NJ, USA)^50^ was used for pooled-analysis. Random-effects model was used for both^51^. Raw data for outcomes and non-events from each study were used to calculate crude odds ratio (OR) with 95% confidence intervals (CI) for each study. The Cochrane Q and I^2^ statistics were calculated to assess heterogeneity between studies^51^. I^2^ <25% were interpreted as low-level heterogeneity^51^. We performed subgroup analysis by study design to decrease inherent selection bias in observational studies^51^. Probability of publication bias was assessed with funnel plot using Egger’s tests^51^ (e-Figure 15-31). If there was statistical heterogeneity in results, further sensitivity analysis was conducted to determine the source of heterogeneity by excluding each study. After significant heterogeneity was excluded, random-effects model was used for meta-analysis. For sensitivity analysis, if we found heterogeneity to be similar with different studies’ exclusion, we chose to present the result with lowest odd’s ratio. P-value <0.05 (2-sided) was considered statistically significant^51^.

### Risk of Bias and Quality assessment

Clinical trials were evaluated using Cochrane risk of bias tool^52^ (e-Table 2) and correlation of quality measures with estimates of treatment effects in meta-analyses of randomized controlled trials tool^53^ (e-Table 3) was used for quality assessment. We used NIH quality assessment Tool for case series^54^ (e-Table 4), case-control (e-Table 5), or cohort studies (e-Table 6). NIH quality assessment tools were based on quality assessment methods, concepts, and other tools developed by researchers in the Agency for Healthcare Research and Quality (AHRQ), Cochrane Collaboration, USPSTF, Scottish Intercollegiate Guidelines Network, and National Health Service Centre for Reviews and Dissemination, consulting epidemiologists and evidence-based medicine experts, with adaptations by methodologists and NHLBI staff. We assessed the certainty of evidence by using GRADE pro profiler^55^ (GRADE working group, McMaster University and Evidence Prime Inc) at the outcome level (e-Table 7).

## Results

### Study Characteristics

A total of 17561 patients from 52 articles^32-35,42-47,56-97^ (Ivermectin n=7509, No Ivermectin n= 10052) were included in the pooled analysis, 14019 patients from 44 studies (Ivermectin n=3913, No Ivermectin n=10106) were included in quantitative synthesis (e-Table 1, Figure 1). Out of 33 trials, 26 were true RCTs (e-Table 1, Figure 1). The remaining 19 studies were observational and included 6 case-series (e-Table 1, Figure 1). Out of 52, 13 articles were available as preprints, Huvemec trial^76^ details were obtained online and clinicaltrials.gov was used for data extraction for 1 trial^65^ (e-Table 1, Figure 1). Due to absence of informed consent, blinding and randomization, we considered Veerapaneni et al’s^93^ study as case control study.

### Primary Outcomes

#### (A) Mortality

##### Meta-analysis Overall

Total 29 studies reported mortality rate in patients receiving Ivermectin (279/3240) vs no-Ivermectin therapy (1347/9407). The odds of mortality in the Ivermectin group were lower compared to control group but the heterogeneity was high (OR 0.54, 95% CI 0.34-0.86, p=0.009; I^2^=75%) (Figure 2A). Evidence was graded very low (e-table 7). In sensitivity analysis, we observed similar mortality benefit with moderate heterogeneity after excluding study by Soto-Becerra et al^92^ (OR 0.49, 95% CI 0.32-0.75, p=0.001; I^2^=52%) (Figure 2B).

**Figure 2:**
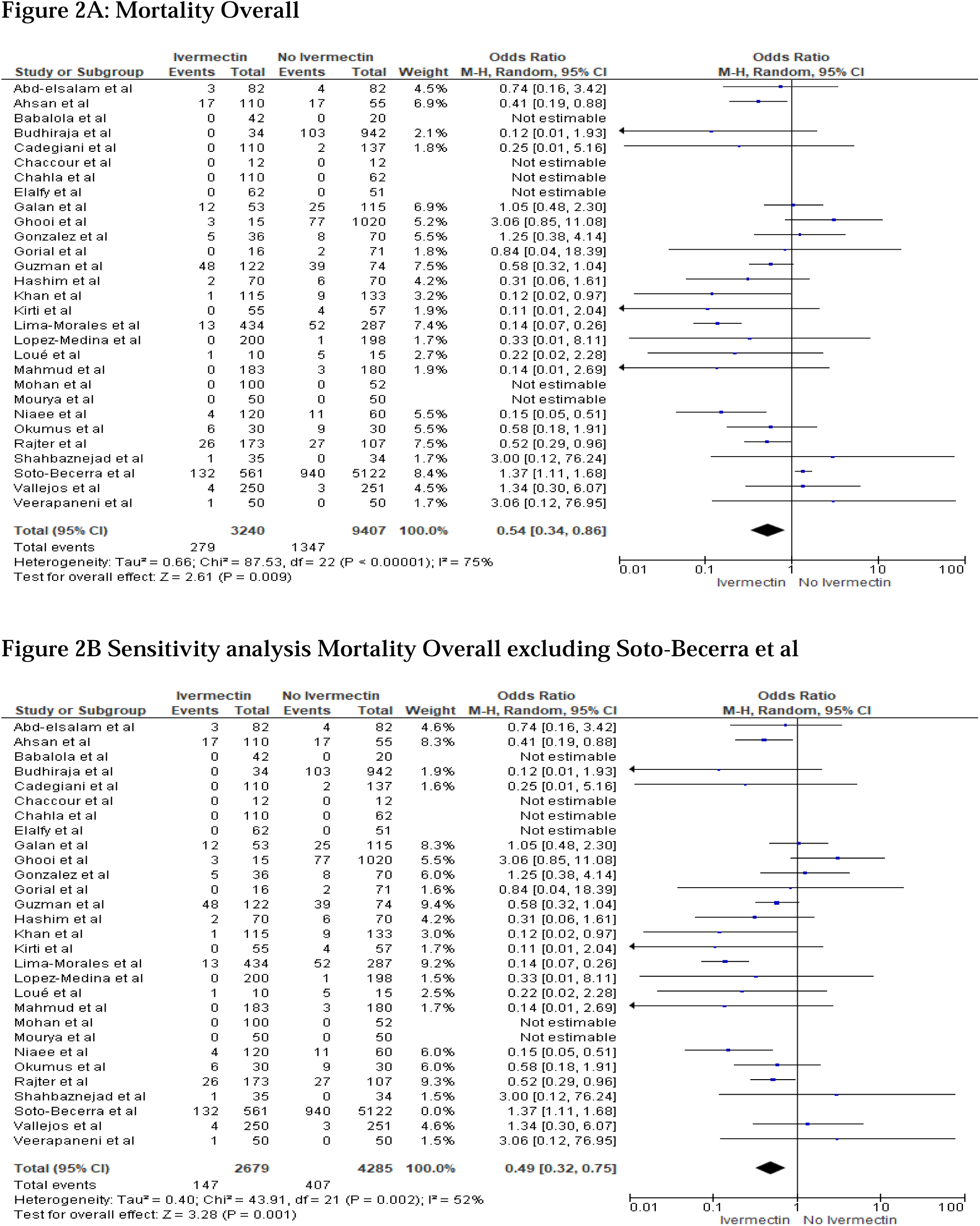
Meta-analysis for Ivermectin use and overall Mortality.

##### Subgroup analysis by study design

###### Subgroup: Clinical trials

We performed subgroup analysis of 18 clinical trials (RCTs N=15, Non-RCTs N=3) and observed similar mortality benefit (OR 0.47, 95% CI 0.25-0.90, p=0.02; I^2^=59%) (Figure 3A) but evidence was graded low (e-table 7). On excluding Lima-Morales et al^43^ for sensitivity analysis, benefit was observed with low heterogeneity but was not statistically significant (OR 0.61, 95% CI 0.37-1.02, p=0.06; I^2^=19%) (e-Figure 1).

**Figure 3:**
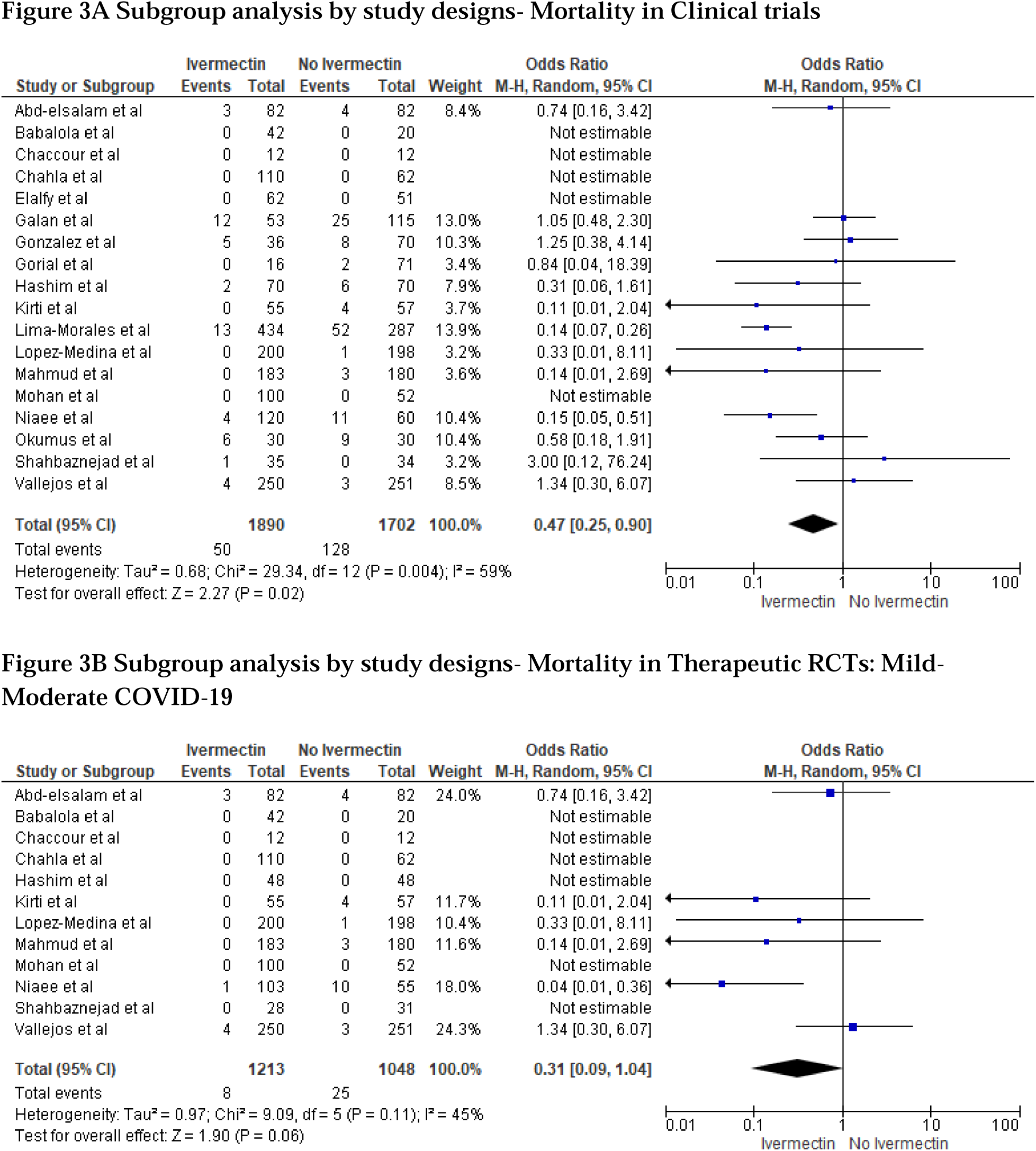
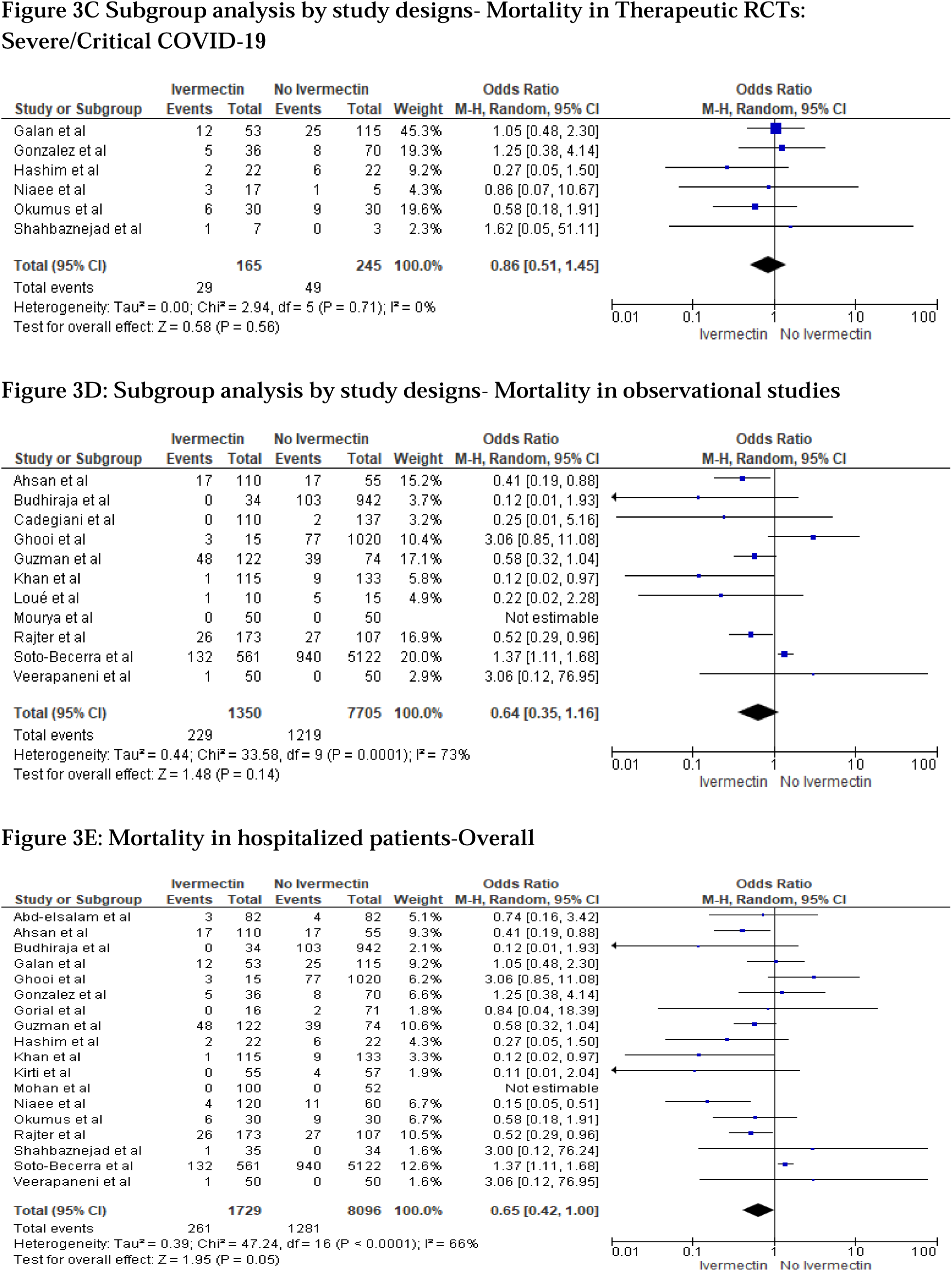
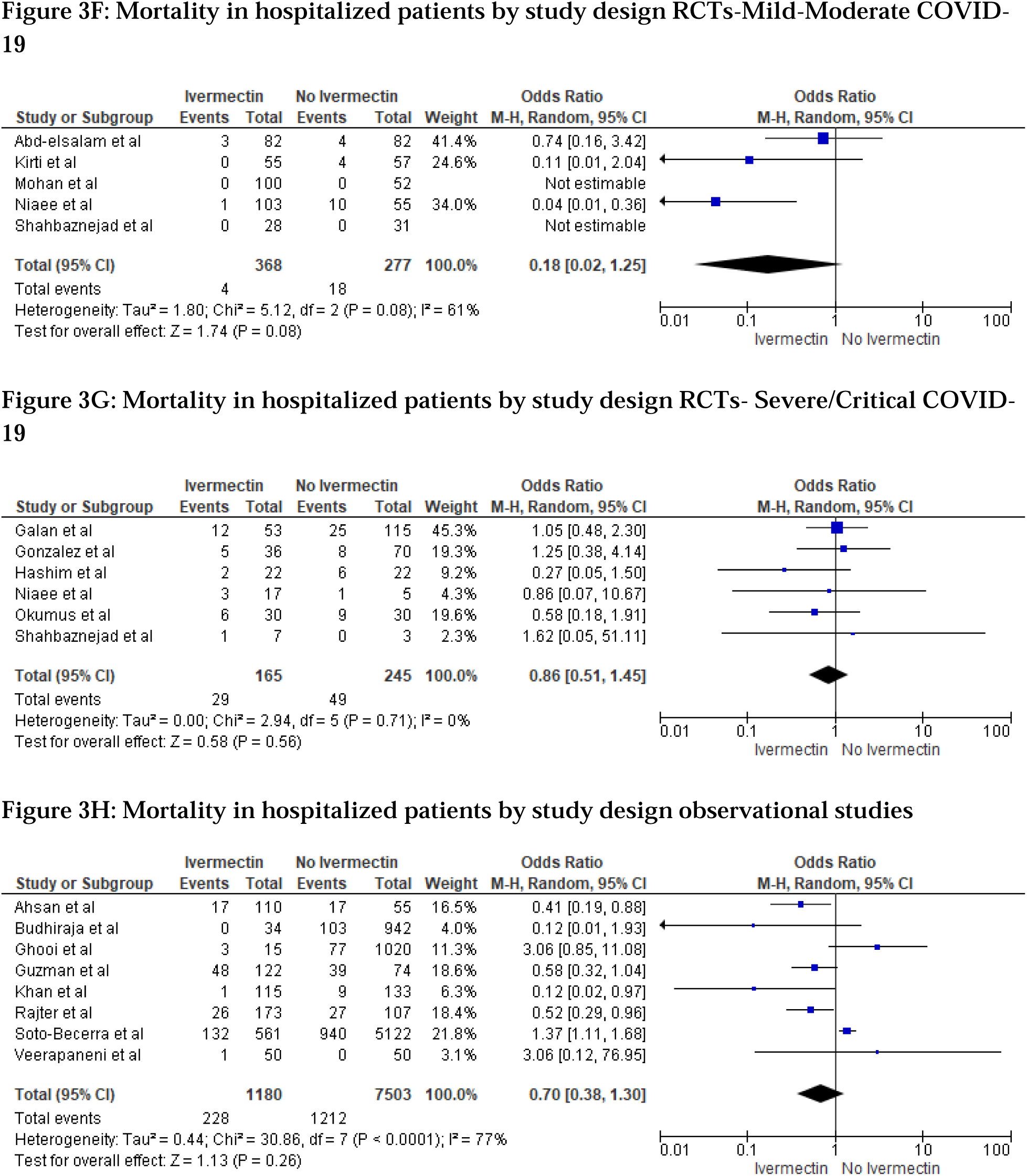
Subgroup analysis for Ivermectin use and mortality by study design and hospitalized patients.

###### Subgroup: RCTs

We analyzed clinical severity-based data in 15 RCTs which included patients treated with Ivermectin on inpatient or outpatient basis. In the mild/moderate subgroup, the odds of mortality were lower with moderate heterogeneity but did not reach statistical significance (OR 0.31, 95% CI 0.09-1.04, p=0.06; I^2^=45%) (Figure 3B). Similarly, mortality benefit was not statistically significant in the severe/critical subgroup (OR 0.86, 95% CI 0.51-1.45, p=0. 56; I^2^=0%) (Figure 3C). Certainty of evidence was low in the mild/moderate and severe/critical subgroup (e-table 7).

In sensitivity analysis, after excluding Vallejos et al^96^ from the mild-moderate subgroup, the odds of mortality decreased with low heterogeneity and reached statistical significance (OR 0.21, 95% CI 0.06-0.69, p=0.01; I^2^=24%) (e-Figure 2). Therefore, in mild/moderate COVID-19 patients, Ivermectin may possibly be useful for reducing mortality when we used as an adjuvant therapy.

##### Observational studies

Mortality benefit was not statistically significant in subgroup analysis of 11 observational studies (OR 0.64, 95% CI 0.35-1.16, p=0.14; I^2^=73%) (Figure 3D) and evidence was graded very low (e-table 7). After excluding Soto-Becerra et al^92^ in sensitivity analysis, we observed statistically significant mortality benefit with moderate heterogeneity (OR 0.54, 95% CI 0.32- 0.92, p=0.02; I^2^=38%) (e-Figure 3).

##### Subgroup analysis for hospitalized patients

###### Overall

In the subgroup analysis for hospitalized patients (studies N=18) mortality benefit was not statistically significant (OR 0.65, 95% CI 0.42-1.00, p=0.05; I^2^=66%) (Figure 3E). Evidence was very low (e-table 7). After excluding Soto-Becerra et al^92^ in sensitivity analysis, we observed statistically significant lower odds of mortality with moderate heterogeneity (OR 0.58, 95% CI 0.39-0.87, p=0.008; I^2^=37%) (e-Figure 4).

###### Subgroup: Inpatient RCTs

We analyzed clinical severity-based data in 9 RCTs with only hospitalized patients. It showed mortality benefit that was not statistically significant in the mild-moderate COVID-19 subgroup (OR 0.18, 95% CI 0.02-1.25, p=0.08; I^2^=61%) (Figure 3F) and in the severe/critical COVID-19 subgroup (OR 0.86, 95%CI 0.51-1.45, p=0.56; I^2^=0%) (Figure 3G). Evidence was graded low in both the subgroup analyses (e-table 7). After excluding Abd-elsalam et al^94^ in sensitivity analysis for mild-moderate COVID-19, mortality benefit was statistically significant (OR 0.06, 95% CI 0.01-0.33, p=0.001; I^2^= 0%) (e-Figure 5).

###### Subgroup: Inpatient Observational studies

In the analysis of 8 observational studies with only hospitalized patients, significant mortality benefit was not seen, and heterogeneity was high (OR 0.70, 95% CI 0.38-1.30, p=0.26; I^2^=77%) (Figure 3H). Evidence was graded very low (e-table 7). In sensitivity analysis, after excluding Soto-Becerra et al^92^ no significant mortality benefit was seen and heterogeneity was moderate (OR 0.58, 95% CI 0.32-1.05, p=0.07; I^2^=50%) (e-Figure 6).

#### (B) Effect of Ivermectin monotherapy compared with Placebo/Standard of Care as per Cochrane’s criteria

We identified 7 RCTs where they investigated the effect of ivermectin monotherapy with or without standard of care compared to placebo or standard of care. Overall, no significant mortality benefit was observed (OR 0.75, 95%CI 0.35-1.63, p=0.47; I^2^=0%) (Figure 4). In severity based subgroup analysis, the result remained same for mild/moderate subgroup (OR 0.71, 95% CI 0.27-1.87, p=0.49; I^2^=0%) and severe/critical subgroup (OR 0.83, 95% CI 0.23- 3.02, p=0.78; I^2^=NA) (Figure 4). Certainty of evidence was moderate overall and in the mild/moderate subgroup, however it was graded low in severe/critical subgroup due to small sample size (e-table 7).

**Figure 4.**
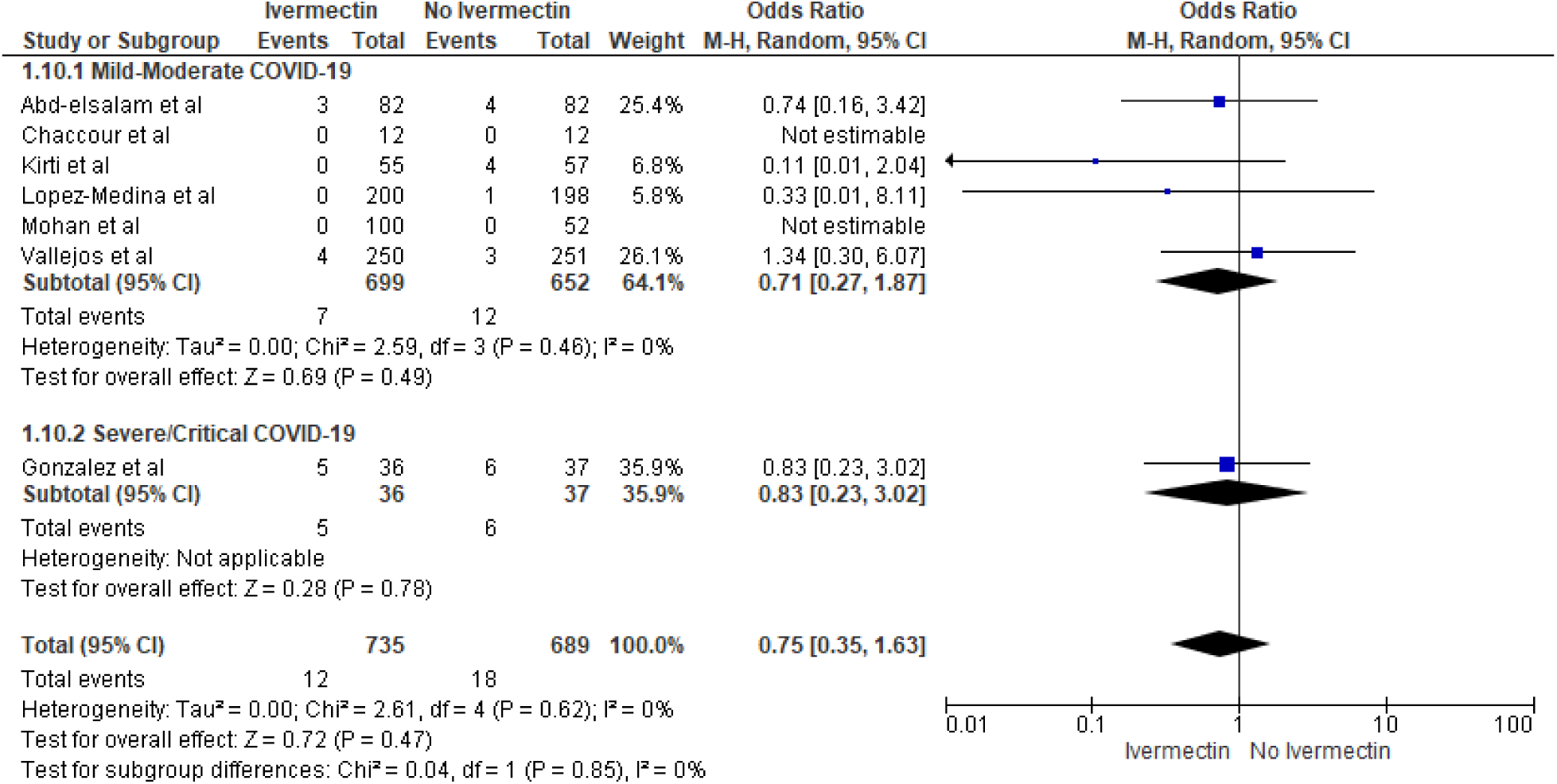
Effect of Ivermectin Monotherapy compared with Placebo/Standard of Care in RCTs as per Cochrane’s criteria.

##### Pooled Analysis for mortality

Pooled analysis of 37 studies out of 52 studies, yielded a rate of 3.6%, 95% CI 2.1-6.3; I^2^=93.87% (e-Figure 7).

#### (C) Need for ICU admission

##### Meta-analysis

In 5 studies (out of 52), 27/263 in the Ivermectin arm, 52/322 in the control arm needed ICU admission. Benefit with Ivermectin was not statistically significant (OR 0.48, 95% CI 0.17- 1.37, p=0.17; I^2^=59%) (Figure 5A). Evidence was graded very low (e-table 7). In sensitivity analysis, after excluding Galan et al^71^ we observed statistically significant benefit (OR 0.32, 95% CI 0.11-0.92, p=0.03; I^2^=27%) (Figure 5B). Based on low grading of evidence and observed low-moderate heterogeneity in analysis, Ivermectin may be helpful in combination therapy in decreasing need for ICU admission. Further validation with well-designed pragmatic platform trials is needed.

**Figure 5:**
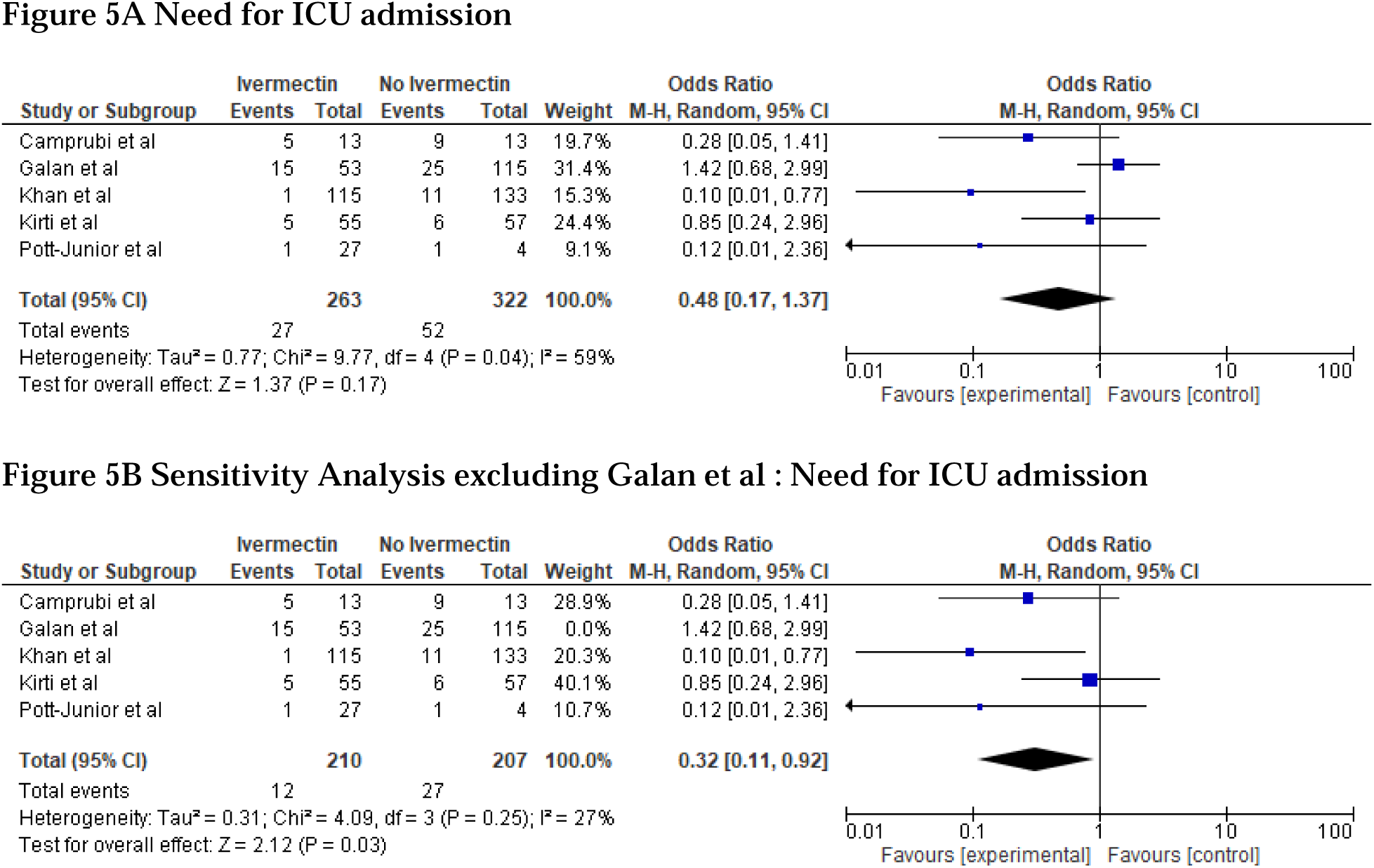
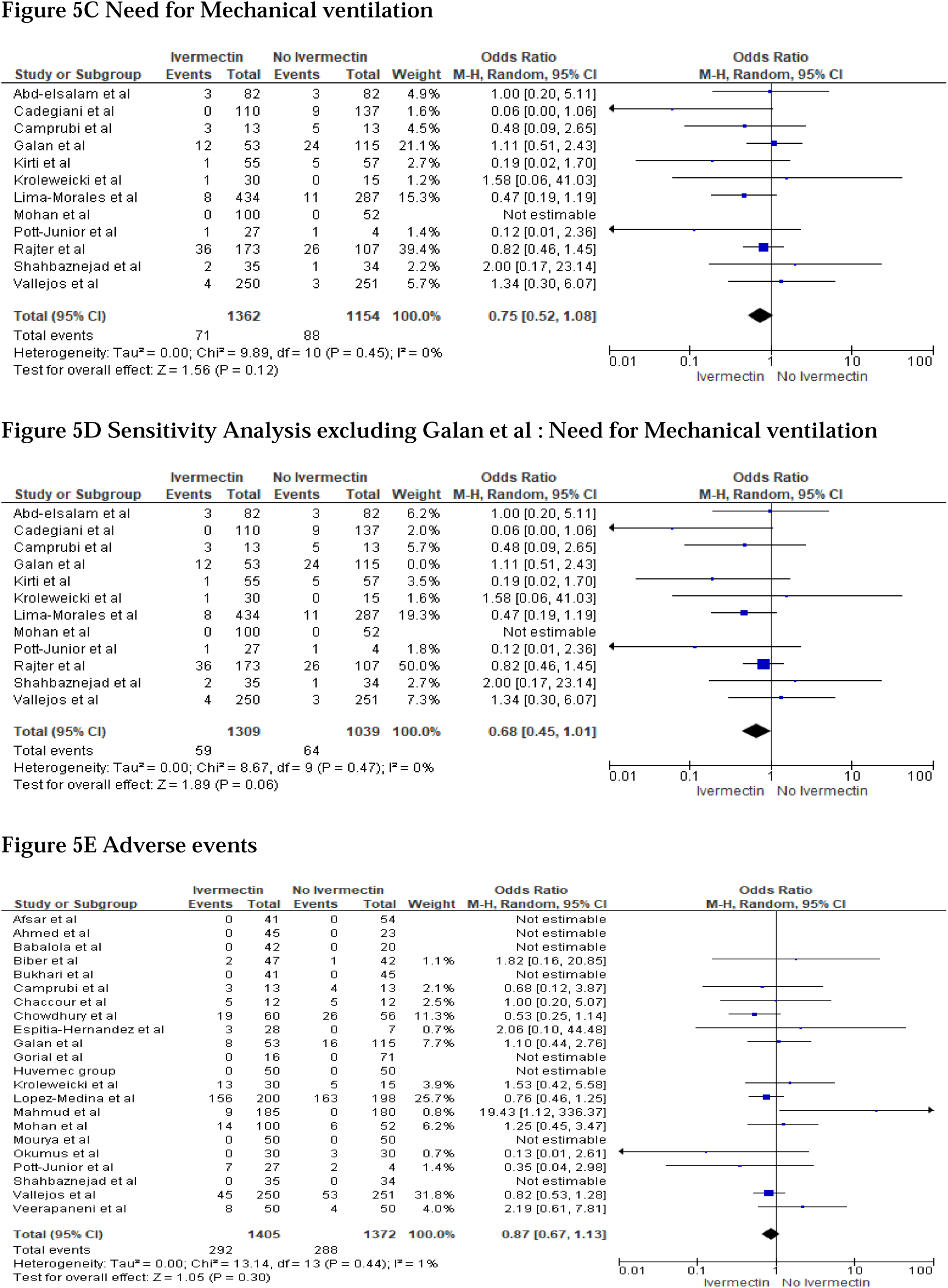
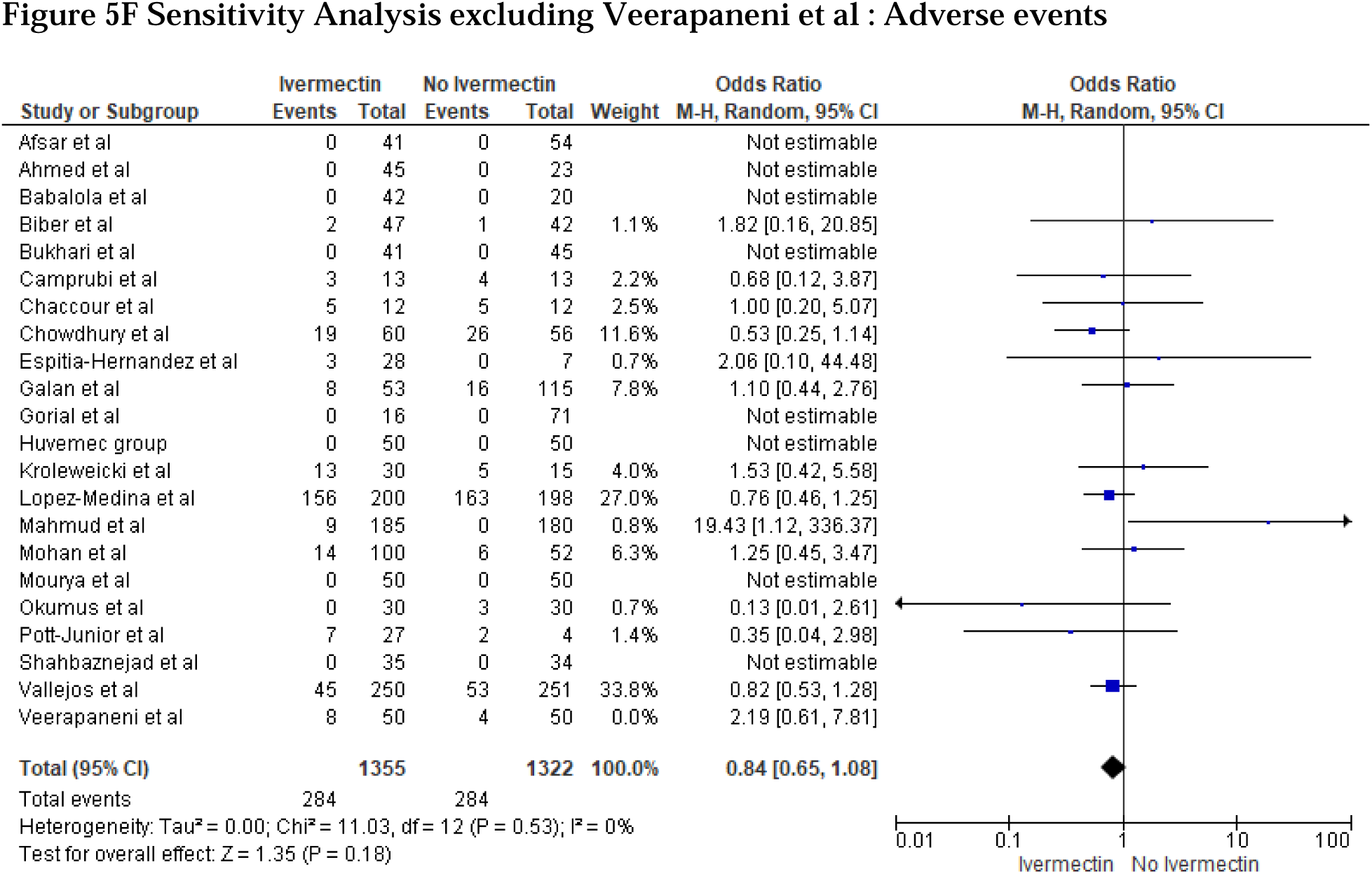
Meta-analysis for Ivermectin use and need for ICU admission, mechanical ventilation and Adverse events.

##### Pooled Analysis

Pooled ICU admission rate obtained from 8 studies out of 52 studies was 5.4%, 95% CI 1.9-14.7; I^2^=91.9% (e-Figure 8).

### Secondary Outcomes

#### (A) Need for Mechanical Ventilation

##### Meta-analysis

Overall, 12 studies documented the need for mechanical ventilation. 71/1362 patients receiving Ivermectin and 88/1154 patients in the control arm required mechanical ventilation. The benefit of Ivermectin was not statistically significant (OR=0.75, 95%CI 0.52-1.08, p=0.12; I^2^=0%) (Figure 5C). Evidence was graded very low (e-table 7). On excluding Galan et al^71^ in sensitivity analysis, benefit observed was not statistically significant and showed no heterogeneity (OR 0.68, 95% CI 0.45-1.01, p=0.06; I^2^=0%) (Figure 5D).

##### Pooled Analysis

The rate of need for mechanical ventilation based on 14 studies (out of 52 studies) was 3.9%, 95% CI 1.7-8.6; I^2^=87.2% (e-Figure 9).

#### (B) Adverse events

##### Meta-analysis

A total of 22 studies reported data for rate of adverse-events in the Ivermectin-arm (292/1405) vs control group (288/1372). We did not find an association between Ivermectin and rate of adverse events as compared to controls (OR 0.87, 95% CI 0.67- 1.13, p=0.30; I^2^=1%). (Figure 5E) but evidence was graded very low (e-table 7). In sensitivity analysis, after excluding Veerapaneni et al^93^ incidence of adverse effects with Ivermectin was similar to that of the control group but was not conclusive (OR 0.84, 95% CI 0.65-1.08, p=0.18; I^2^=0%) (Figure 5F).

##### Pooled Analysis

A total of 38 studies (out of 52 studies) yielded a pooled rate of 6.6%, 95% CI 3.9-11.1; I^2^=92.2% (e-Figure 10).

#### (C) Viral Clearance

Twenty-two studies reported data for patients achieving viral clearance: 1036/1327 in the Ivermectin arm while 790/1206 patients in the non-Ivermectin arm tested negative for RT-PCR. The analysis indicated that patients in the Ivermectin-arm showed higher odds of achieving viral clearance when compared to the control group at a given point in time (OR 3.52,95%CI 1.81- 6.86, p<0.01; I^2^=85%) (figure 6A). Evidence level was low (e-table 7). When we excluded Mourya et al^85^ in the sensitivity analysis, similar benefit was observed but heterogeneity remained high (OR 2.79, 95%CI 1.52-5.10, p<0.01; I^2^=81%) (e-Figure 11).

**Figure 6:**
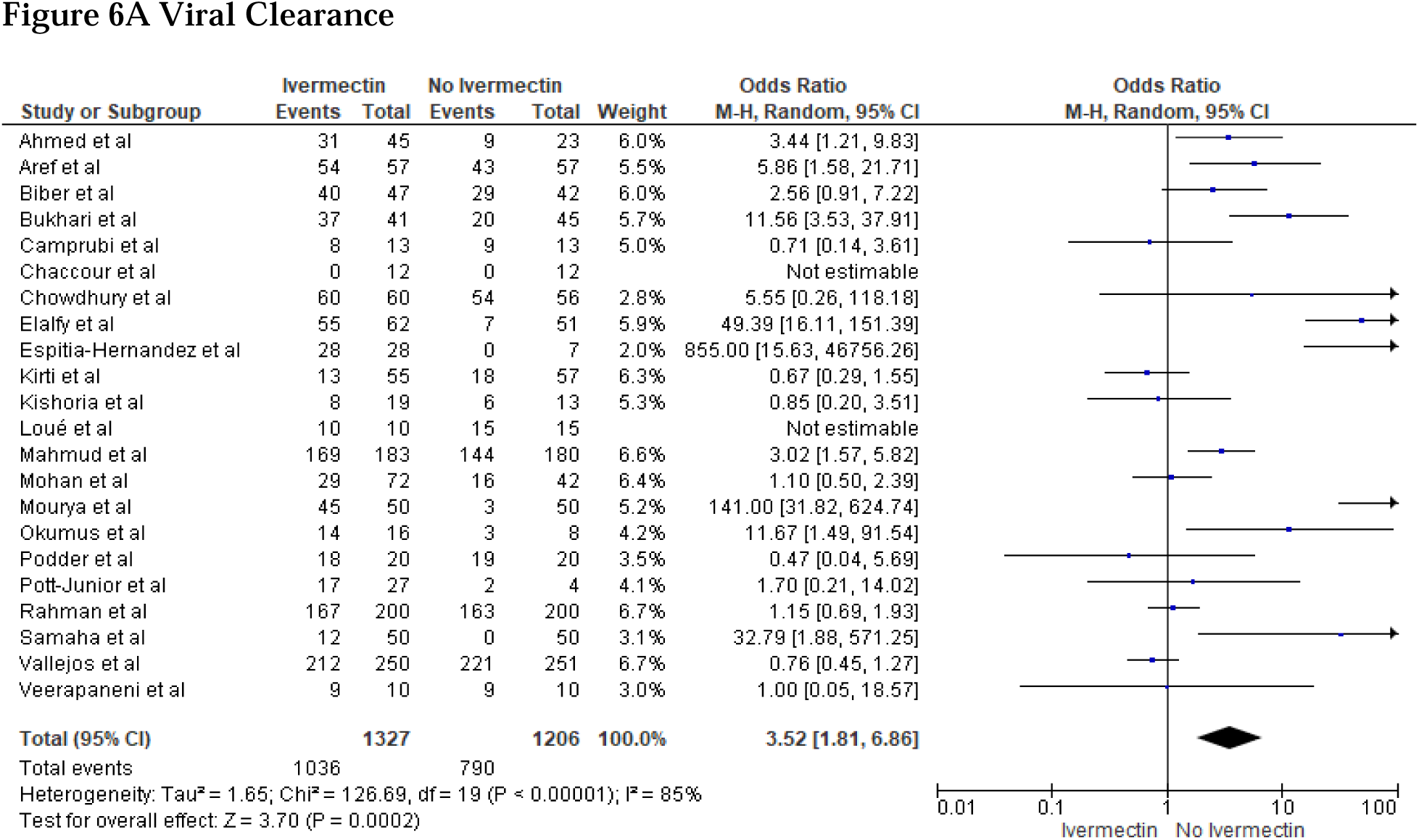
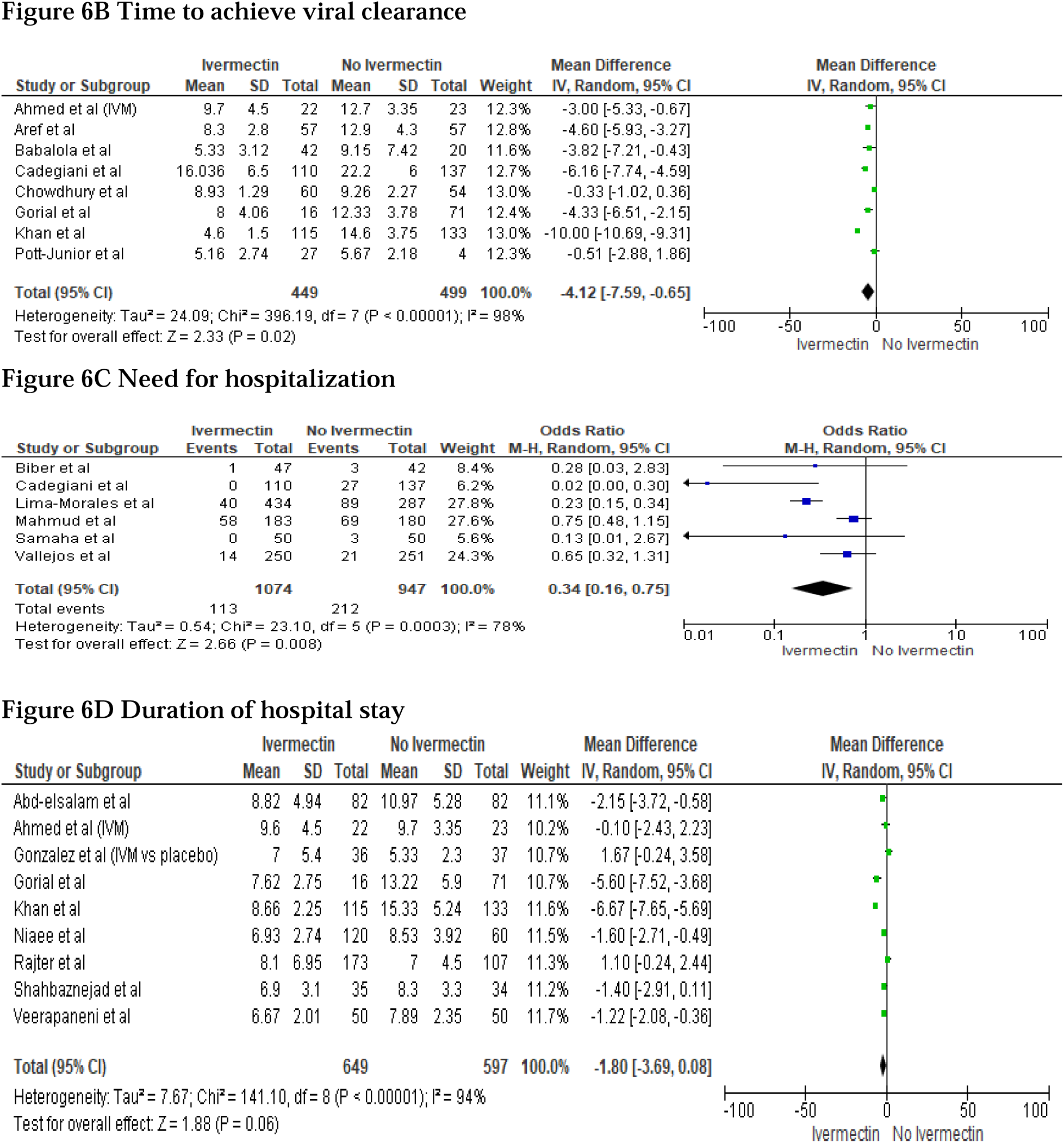
Meta-analysis for Ivermectin use and need for Viral Clearance, Time to achieve viral clearance, Need for hospitalization and Duration of hospital stay.

#### (D) Time to achieve Viral Clearance

Eight studies presented data for the time needed for viral clearance in the Ivermectin receiving arm vs that in the non-Ivermectin arm. The results show that the time needed to achieve viral clearance was shorter by over 4 days in the Ivermectin group as compared to the non-Ivermectin group, but with high heterogeneity (MD -4.12,95%CI -7.59,-0.65, p=0.02; I^2^=98%) (figure 6B). Evidence level was low (e-table 7). In sensitivity analysis, benefit was seen but heterogeneity remained high after excluding Khan et al’s study^77^ (MD -3.24, 95%CI -5.37,- 1.10, p=0.003; I^2^=92%) (e-Figure 12).

#### (D) Need for hospitalization

Six studies described information for the number of patients requiring hospitalization when receiving Ivermectin (113/1074) as compared to those not receiving it (212/947). As per the analysis, Ivermectin group showed lower need for hospitalization as compared to controls but with high heterogeneity (OR 0.34, 95%CI 0.16-0.75, p=0.008; I^2^=78%) (Figure 6C). Evidence was low grade (e-table 7). On excluding Lima Morales et al^43^ in sensitivity analysis, similar benefit was seen with moderate heterogeneity but without statistical significance (OR 0.43, 95% CI 0.18-1.01, p=0.05; I^2^=59%) (e-Figure 13).

#### (F) Length of hospital stay

We analyzed data from 9 studies regarding the duration of hospital stay in the Ivermectin vs control arm. In this meta-analysis, we found that the benefit of Ivermectin in reducing the duration of hospital stay was not statistically significant and heterogeneity was high (MD -1.80, 95% CI -3.69,0.08, p= 0.06; I^2^=94%) (Figure 6D). Evidence was low grade (e-table 7). In the sensitivity analysis, we excluded Khan et al’s study^72^ and the duration of hospital stay was shorter by one day in the Ivermectin arm compared to non-Ivermectin arm with high heterogeneity and no statistical significance (MD -1.16, 95%CI -2.42,-0.1, p=0.07; I^2^=84%) (e-Figure 14).

## Discussion

Our meta-analysis suggests that the role of Ivermectin in improving patient survival in COVID-19 is uncertain. However, Ivermectin might help more patients achieve viral clearance, in a shorter duration of time and may also help reduce the need for hospitalization when used in combination therapy. Its benefit in reducing the need for admission to the ICU, mechanical ventilation and the duration of hospital stay cannot be concluded with confidence. Its relation to the incidence of adverse effects is inconclusive. Further, our pooled analysis indicates that Ivermectin treatment may be linked to lower death rate and better clinical outcomes with a low incidence of adverse effects. The inherent methodological limitations and risk of bias conferred variable evidence grading in the findings in various patient outcomes.

### Mortality

Several systematic review and meta-analysis were performed to investigate the efficacy of ivermectin for the treatment and prevention of COVID-19, with mixed results in meta-analyses and conclusions. Meta-analyses done by Padhy et al ^98^ (studies N=3), Hill et al^99^ (studies N=6) (**retracted**^100^), Lawrie et al^101^ (studies N=5), Nardelli et al^102^ (studies N=7), Kow et al^103^ (studies N=6), Bryant et al^104^ (studies N=13), Kory et al^105^ (studies N=10), Hariyanto et al^106^ (studies N=8) and the BIRD group^107^ (studies N=13) found Ivermectin to be conclusively linked to lower mortality when compared to usual treatment. A network meta-analysis^108^ (studies N=2) reported that Ivermectin was linked to lower mortality with a very close statistical significance. Casteneda-Sabogal et al’s meta-analysis^109^ (studies N=6) and Roman et al^110^ (studies N=5) found no conclusive association of Ivermectin with reduced mortality. Of note, the above three studies^108-110^ reported low or very low certainty of evidence. However, most of the above analyses have included the retracted study by Elgazzar et al^48^. Recently published Cochrane’s review^111^ of highly selected low-risk bias studies aimed at evaluating the true-effect of Ivermectin in COVID-19. They expressed uncertainty regarding the effect of Ivermectin on COVID-19 mortality based on 2 outpatient and 2 inpatient RCTs^111^. Our sample size exceeds (studies N=29) that of all the aforementioned studies. We observed mortality overall (N=29) and in the clinical trials subgroup (N=18) meta-analysis. However, no mortality benefit was observed in therapeutic RCTs (mild-moderate N=12, severe/critical N=6), inpatient RCTs (mild-moderate N=5, severe/critical N=6) and observational studies (overall N=11, Inpatient N=8). Using the selection criteria used in Cochrane’s meta-analysis, we did not observe a mortality benefit when we limit the analysis for Ivermectin monotherapy in comparison to standard of care or placebo^111^. We would like to emphasize the fact that the observed mortality benefit in some analyses was frequently seen when Ivermectin was used in combination therapy especially with vitamins, trace elements, other antimicrobials, and steroids. Beneficial effect of steroids has been already been established, which was frequently used as part of standard of care^112^. As we know proper nutrition and activation of the immune system are two crucial elements in fighting the COVID-19 infection^113^ therefore it is possible that supplemental vitamins and minerals may help in COVID-19 infection. However, clinical evidence does not currently support routine use of any of these agents for the prevention or treatment of COVID-19, unless a patient has a confirmed or suspected micronutrient deficiency^114^. We speculate that the observed mortality benefit of Ivermectin was influenced by the use of these supplements, either through prescription or patient’s own decision for over the counter use.

Our pooled analysis resulting in a low mortality rate of 3.6% with use of Ivermectin, mostly as an adjuvant therapy, is suggestive of its positive impact in lowering mortality in COVID-19 patients. For a valid comparison, it is important to factor in the time window during which the individual studies were conducted. Most studies recorded their data in the II&III quarter of 2020 when mortality rates with various treatments were often higher in respective countries^115^. For instance, the mortality benefit with Ivermectin has been observed on a magnified basis in a population study of 25 Peruvian states. The analysis of this mass administration data revealed a 14 fold drop in deaths a couple months after starting of large-scale Ivermectin use and a 13 fold increase since limiting its use following change in Peru’s political scenario^116^.

### Need for ICU admission and Mechanical Ventilation

Due to the small number of studies and their small individual sample size, benefit with Ivermectin in reducing ICU admissions or mechanical ventilation cannot be concluded with confidence. In our pooled analysis, we observed significantly lower rates for ICU admission (5.4%) and mechanical ventilation (3.9%) in Ivermectin-treated patients, mostly as an adjuvant therapy, when compared to other treatments^117-119^. It must be noted that our pooled analysis mostly included mild/moderate patients, which may skew the results in favor of Ivermectin.

### Adverse events

Our analysis for adverse events was inconclusive. We noticed that most adverse events were non-serious: nausea, vomiting, abdominal pain, diarrhea, pruritus, lethargy, vertigo, tingling, numbness, anxiety, mild hyperglycemia etc. Yet, very few patients reported serious events like organizing pneumonia, hyponatremia, erosive esophagitis, infections/infestations. Erosive esophagitis can be attributed to concomitant use of Doxycycline^120^; hyponatremia^80^ was likely from SIADH-a rare COVID-19 complication^121^. Only Lopez-Medina et al^81^ (Ivermectin dose = 1500ug/Kg over 5 days) and Mahmud et al^45,122^ (Ivermectin dose =12mg twice daily for 5 days) reported drug discontinuation due to adverse-reactions. Given the non-serious nature of most adverse-reactions, and a low pooled rate of 6.6% in comparison to other available treatments^23,123,124^, we suggest that Ivermectin was generally well tolerated.

### Viral Clearance and time to achieve Viral Clearance

We observed that patients who received Ivermectin in combination with other drugs were far more likely to test negative for RT-PCR as compared to the control group. Hariyanto et al’s meta-analysis^106^ (N=9) supports our findings. Contrary to our results, Roman et al^110^ (N=4) and Cochrane’s review^111^ found no such difference.

Similarly, when used in combination therapy, Ivermectin appeared to have significantly shortened the time to achieve viral clearance when compared to the non-Ivermectin arm. This finding was also observed in the analysis of Lawrie et al^101^ (N=2), Hariyanto et al^106^ (N=6) and the BIRD group^107^ (N=4), however, Lawrie et al’s^101^ results were not statistically significant. It must be noted that these outcomes are subject to technical variations because of the different laboratory parameters as well as the day at which viral clearance was measured.

### Need for hospitalization and duration of hospital stay

Our analysis suggests that Ivermectin treatment may help reduce the need for hospitalization. Another study by the BIRD group^107^ (N=2) also found this association, yet their analysis did not reach statistical significance.

In our analysis, we found trends of shorter duration of hospital stay but our results were not conclusive, hence more data is needed to confirm this finding. Three other studies^101,106,107^ also found similar benefit, only one^107^ was not statistically significant. While Roman et al^110^ observed that Ivermectin was associated with a longer hospital stay; but without statistical significance. Recently, Cochrane’s review^111^ also did not find an association between Ivermectin and duration of hospitalization. We must acknowledge the high variation in clinical decision making as well as the availability of resources with respect to hospitalization outcomes.

Our analysis for the true effect of Ivermectin is inconclusive, which supports the conclusion of Cochrane’s review^111^ that Ivermectin may or may not be effective by itself. COVID-19, being a complex, multi-stage, and multi-system disease, requires a multi-pronged approach. Hazan et al^97^ enumerated several microbial diseases currently being successfully treated by a combination of multiple drugs with different individual mechanisms of actions. They highlighted another benefit of multidrug therapy: to avert cases of drug-resistance such as those occurring with Ivermectin-monotherapy against parasitic infections. This is crucial since the newly emerging SARS-CoV2 strains have perpetuated the pandemic in spite of desperate measures taken world-wide. Also, this approach may address concerns regarding the adverse effects associated with high dose of Ivermectin. Because, drug combinations can possibly boost drug potency such as with the combination used by Hazan et al^97^, thus rendering moderate doses to be more effective. Therefore, it is imperative that we thoroughly explore Ivermectin as a part of a multidrug regimen.

On the starting point of the COVID-19 disease spectrum is the acute infectious phase that lasts for up to 2 weeks and is characterized by viral replication^125^. This is followed by the hyper-inflammatory state marked by rise in inflammatory markers^125^. Those surviving this phase may experience long-term multi-organ sequelae for up to six months or even a year^126,127^. Patients progressing to the severe stage require hospitalization and mortality rates among hospitalized COVID-19 patients can be as high as 25%^128^. Dwindling economies, overburdened healthcare systems and the consequent dearth of resources further worsen the prognosis of hospitalized patients, especially in low-income countries. Once progressed to the critical stage, mortality rate of around 50% has been recorded^129^. The management of severe and critical patients mainly consists of supportive treatment for complications of severe COVID-19^130^. The available evidence of the efficacy and safety of the recommended drugs for severe and critical COVID-19 patients is not promising^131-134^. Even before the pandemic, much emphasis has been laid on the importance of preventive and early treatment strategies for ARDS and sepsis to achieve a higher survival rate in patients^135-138^. Drawing upon these recommendations, we hypothesize that employing Ivermectin as an adjuvant therapy for early treatment of COVID-19 cases may allow the body to mount an adequate and well-regulated immune response, thus leading to better outcomes. Considering the bleak prognosis of COVID-19 at the advanced stage, we postulate that we may potentially improve patient outcomes by actively arresting its progression to severe stages with effective and safe early adjuvant treatments like Ivermectin. Finally, we suggest that understanding the course of this disease-both pathophysiological and symptomatic, and leveraging it is key to achieving the best outcomes at both-patient and population level.

### Strengths

To our knowledge, this is the largest patient sample size for a systematic review and meta-analysis on the use of Ivermectin for COVID-19. This study integrates data of approximately 17,000 patients from 52 studies-both trials and observational, conducted globally. Additionally, we obtained robust evidence by conducting various subgroup analyses. One noteworthy Ivermectin study, a large clinical trial^48^, was recently withdrawn on grounds of ethical misconduct. Its strongly positive findings accounted for significant effect size in the meta-analyses published so far. This has led us to question the existing conclusions regarding the efficacy of this drug. Ours would be the first meta-analysis that excluded the flawed study to clear the air regarding the effect of Ivermectin in COVID-19 for a stronger empirical evidence of Ivermectin’s efficacy. We also excluded studies which did not meet the rigor of true RCTs in the ‘RCTs’ subgroup. We used GRADE-pro tool^55^ to rate certainty of evidence for all outcomes, which was not done in past to provide more insight. The GRADE framework is known to be the most widely used assessment to rate quality of evidence^55^. Sensitivity analyses were performed with the aim of testing the robustness of results. Moreover, we performed pooled analyses to incorporate data from uncontrolled studies.

### Limitations

Our results are to be interpreted carefully bearing in mind that we included several non-peer-reviewed articles to obtain the latest data. We cannot preclude the possibility of unreported biases and confounders that could have been recognized by a peer-review process. Furthermore, there was significant heterogeneity in treatment protocols used for dosage, duration, route of administration in interventional and control group. Our analyses may not have accounted for confounding by concomitant therapy. We only analyzed the mortality outcome as per Cochrane’s strict selection criteria for Ivermectin monotherapy compared to standard of care/placebo. Patient demographics and outcome measures varied among different studies. Few studies documented the inclusion of patients <18 years of age. However, the mean/median and dispersion values consistently suggested a predominantly adult population. The clinical classification for severity was inconsistent across the included studies. Most studies were conducted on patients with mild /moderate presentation, which limits the generalizability of our results.

Apart from this, there was insufficient data regarding the time of treatment in the course of the disease, hence inferences could not be made in that regard. We could not confirm adequate control-matching in some observational studies. We advise discretion in interpretation of our results for adverse-event rate since most of the reported adverse-effects could not be measured objectively by investigators. Because most of the studies were conducted in developing nations, not everyone underwent RT-PCR testing owing to limited resources; clinical diagnosis was used instead. Even though RT-PCR is the gold standard for diagnosis, it is subject to its limitations. By virtue of its design, we postulate that this meta-analysis formulated with a random-effects model may have addressed some of the heterogeneity stemming from the above factors. Lastly, we must acknowledge the subjective nature of the GRADE tool used to evaluate the level of evidence^55^.

### Current status of Ivermectin

In February 2021, the NIH declared that it neither recommends for or against using Ivermectin for COVID-19^139^ due to inadequate data. For further guidance, they require evidence from larger, well-designed and well-executed trials. As of March 5,2021, the FDA has warned people against using Ivermectin for COVID-19^140^, specifically against self-medicating with veterinary Ivermectin formulations^140,141^. They are yet to review the evidence but have initiated a preliminary research. In late March, the EMA (European Medicines Agency) stated that Ivermectin cannot be recommended for the prevention or treatment of COVID-19 outside clinical trials^142^. Similarly, the WHO suggested limiting Ivermectin use to clinical trials until more data is available^143^. A large phase 3 clinical trial evaluating the use of Ivermectin in COVID-19 patients with mild to moderate symptoms is yet to be concluded^144^. The NIH, FDA, CDC, EMA and several other important government and private sector bodies will collaborate on this “Master protocol” titled ACTIV-6 that focuses on rigorous testing of up to seven repurposed drugs^145^. Recently, the CDC issued warning against the use of non-prescription Ivermectin as well as the ingestion of Ivermectin-containing topical or veterinary products and advised seeking immediate medical care for any adverse effects arising out of such events^146^.

## Conclusion

In summary, based on updated evidence, benefit of Ivermectin as monotherapy or in combination treatment for lowering the COVID-19 associated mortality is inconclusive due to lack of high-quality RCTs. Ivermectin combination therapy may play a role in controlling the viral replication and preventing the need for hospitalization. Using well-designed larger observational studies^147,148^ and clinical trials, we need to further investigate its efficacy, its ideal dosage and timing in the disease course, drug interactions and possible synergistic drug combinations to achieve maximum benefit. We also need to evaluate the benefit of Ivermectin for treating the various emerging strains as well as its role in vaccinated individuals. We propose pragmatic practice embedded platform trials^149^ to test this and other re-purposed and novel therapies specifically in severe COVID-19 patients and other critically ill patients. We recommend the medical practitioners to exercise caution until further evidence is available and use their best clinical judgement while prescribing Ivermectin in COVID-19 patients.

## Data Availability

The data is available on our site.

## Acknowledgment

We acknowledge Dr. Anant Mohan, Dr. Asma Asghar, Dr. Flavio Cadegiani, Dr. Houssam Raad, Dr. José Morgenstern, Dr. Morteza Niaee, Dr. Nasir Afsar, Dr. Nurullah Okumus, Dr. Olufemi Babalola, Dr. Pablo Méndez-Hernández, Dr. Ravi Kirti, Dr.Ravindra Ghooi, Dr. Reaz Mahmud and Dr. Tasnim Ahsan for their correspondence and contribution.

## List of abbreviations

ARDS: Acute respiratory distress syndrome
CDC: Centers for Disease Control and Prevention
COVID-19: Coronavirus Disease 2019
DNA: Deoxyribonucleic Acid
EMA: European Medicines Agency
FDA: Food and Drug Administration
ICU: Intensive Care Unit
IRB: Institutional Review Board
NIH: National Institutes of Health
RCT: Randomized controlled trial
RNA: Ribonucleic Acid
RT-PCR: Reverse Transcription Polymerase Chain Reaction
IMP: Importin
SARS-CoV-2: Severe Acute Respiratory Syndrome Coronavirus 2
WHO: World Health Organization

## Figure and Table Legends

**e-Figure 1 Sensitivity analysis excluding Lima-morales et al: Mortality in Clinical trials**

**e-Figure 2 Sensitivity analysis excluding Vallejos et al: Mortality in Therapeutic RCTs: Mild-Moderate COVID-19**

**e-Figure 3 Sensitivity analysis excluding Soto-Becerra et al: Mortality in observational studies**

**e-Figure 4 Sensitivity analysis excluding Soto-Becerra et al: Mortality in hospitalized patients-Overall**

**e-Figure 5 Sensitivity analysis excluding Abd-elsalam et al: Mortality in hospitalized patients by study design RCTs: Mild-Moderate COVID-19**

**e-Figure 6 Sensitivity Analysis excluding Soto-Becerra et al: Mortality in inpatient observational studies**

**e-Figure 7 Pooled Analysis for Mortality**

**e-Figure 8 Pooled analysis of need for ICU admission**

**e-Figure 9 Pooled analysis of Mechanical Ventilation**

**e-Figure 10 Pooled Analysis of Adverse events**

**e-Figure 11 Sensitivity Analysis excluding Maurya et al : Viral Clearance**

**e-Figure 12 Sensitivity Analysis excluding Khan et al : Time to achieve Viral Clearance**

**e-Figure 13 Sensitivity Analysis excluding Mahmud et al : Need for hospitalization**

**e-Figure 14 Sensitivity Analysis excluding Gonzalez et al : Duration of hospital stay**

**e-Figure 15 Funnel Plot overall mortality**

**e-Figure 16 Funnel Plot Mortality in Clinical Trials**

**e-Figure 17 Funnel Plot Mortality in RCTs: Mild/Moderate COVID-19**

**e-Figure 18 Funnel Plot Mortality in RCTs: Severe/Critical COVID-19**

**e-Figure 19 Funnel Plot Mortality in Observational studies**

**e-Figure 20 Funnel Plot Mortality in Hospitalized patient overall**

**e-Figure 21 Funnel Plot Mortality in Inpatient RCTs: Mild-Moderate COVID-19**

**e-Figure 22 Funnel Plot Mortality in Inpatient RCTs: Severe/Critical COVID-19**

**e-Figure 23 Funnel Plot Mortality in Inpatient Observational studies**

**e-Figure 24 Funnel Plot for Mortality with Ivermectin Monotherapy as per Cochrane’s Criteria**

**e-Figure 25 Funnel Plot Need for ICU Admission**

**e-Figure 26 Funnel Plot Need for Mechanical Ventilation**

**e-Figure 27 Funnel Plot Adverse event**

**e-Figure 28 Funnel Plot Need for Hospitalization**

**e-Figure 29 Funnel Plot duration of Hospital stay**

**e-Figure 30 Funnel Plot Incidence of Viral Clearance**

**e-Figure 31 Funnel Plot Time to achieve viral clearance**

**e-table 1. Study characteristic table for all included studies**

**e-table 2. Cochrane Risk of bias assessment of the trials included in the study**

**e-table 3. Correlation of quality measures with estimates of treatment effects assessment of the trials those were included in the study**

**e-table 4. NIH quality assessment Tool for case series included in the study**

**e-table 5. NIH Quality Assessment of Case-Control Studies**

**e-table 6. NIH Quality Assessment of Observational Cohort and Cross-Sectional Studies**

**e-table 7. Certainty of the evidence (GRADE) Profile at Outcome Level**

**e-table 8. Ongoing clinical trials**

