## Supplemental Table 1 for "An Updated Systematic Review and Meta-Analysis of Mortality, Need for ICU admission, Use of Mechanical Ventilation, Adverse effects and other Clinical Outcomes of Ivermectin Treatment in COVID-19 Patients"

**e-table 1. Study characteristic table for all included studies**

| **Author** | **Inpatient/ Outpatient** | **Type of study design** | **Country** | **Cases** | **Controls** | **Total patients** | **Treatment for controls** | **Treatment for Cases** | **Concomitant treatment for cases** | **Pre-print or peer reviewed** |
| --- | --- | --- | --- | --- | --- | --- | --- | --- | --- | --- |
| Abd-Elsalam et al^94^ | inpatient | 1:1 randomized, open‐label, parallel‐group clinical trial | Egypt | 82 | 82 | 164 | Standard of care | Ivermectin (12 mg every day for 3 days) plus standard of care | Information not available | Peer- reviewed |
| Afsar et al^56^ | Outpatient | Case-control | Pakistan | 54 | 41 | 95 | Azithromycin, Hydroxychloroquine; Corticosteroids, antibacterials, Zinc, low dose Aspirin | Ivermectin 12 mg PO daily for six days | Azithromycin & HCQ; Corticosteroids, antibacterials, zinc, low dose aspirin | Pre-print |
| Ahmed et al^57^ | Inpatient | RCT(randomized, double-blind, placebo-controlled trial) | Bangladesh | 45 | 23 | 68 | Placebo | first arm: oral ivermectin alone 12 mg once daily for 5 days,  second arm: oral ivermectin 12mg single dose | second arm: doxycycline | Peer-reviewed |
| Ahsan et al^58^ | Inpatient | Case series | Pakistan | 110 | 55 | 165 | Doxycycline, famotidine, remdesivir, methylprednisolone, Tocilizumab, CP, enoxaparin, Azithromycin | Ivermectin (150-200 ug/kg/day (on two successive days), | Doxycycline, famotidine, remdesivir, methylprednisolone, Tocilizumab, CP, enoxaparin, Azithromycin | Peer-reviewed |
| Alam et al^59^ | Outpatient | Case series | Bangladesh | 100 | - | 100 | - | Ivermectin 0.2 mg/kg single dose | Doxycycline with supportive treatment | Peer-reviewed |
| Aref et al^95^ | outpatient | prospective clinical trial | Egypt | 57 | 57 | 114 | Standard of care | ivermectin nanosuspension nasal spray twice daily plus the Egyptian protocol of treatment | Egyptian protocol of treatment : 1. Paracetamol 500 mg intravenously every 6 hours.  2. Hydroxychloroquine 500 mg/12 h with close monitoring of liver and kidney functions.  3. Azithromycin 1 g first day, then 500 mg per day for 3 days or clarithromycin 500 mg every 12 hfor 7–14 days.  4. Oseltamivir 150 mg/12 h for 5 days.  5. Ascorbic acid 500 mg/12 h.  6. Cyanocobalamin IV once daily. | Peer- reviewed |
| Babalola et al^60^ | Outpatient +Inpatient | RCT (Randomized, double blinded, placebo controlled trial) | Nigeria | 42 | 20 | 62 | Lopinavir/Ritonavir daily for 2 weeks. Some required: dexamethasone, enoxaparin, and supplemental oxygen; supplements taken by patients: Zinc, ascorbic acid, vitamin D and Azithromycin. | group A: Ivermectin 6mg every 84 hrs for 2 weeks;  Group B: Ivermectin 12mg every 84hrs for 2 weeks | Some required:, dexamethasone, enoxaparin, and supplemental oxygen; supplements taken by patients: Zinc, ascorbic acid, vitamin D and Azithromycin. | Peer-reviewed |
| Bhattacharya et al^34^ | Inpatient | Case series | India | 148 | - | 148 | - | Ivermectin single dose 12mg | Atorvastatin, Inj. N-acetyl cysteine, standard of care | Peer-reviewed |
| Biber et al^61^ | Outpatient | RCT(double blinded) | Israel | 47 | 42 | 89 | Placebo | 0·2 mg/kg for 3 days | - | Pre-print |
| Budhiraja et al^62^ | Inpatient | Case series | India | 34 | 942 | 976 | HCQ+- Azithromycin/ tocilizumab/ Convalescent plasma therapy/ steroids | Ivermectin | HCQ+- Azithromycin/ tocilizumab/ Convalescent plasma therapy/ steroids | Pre-print |
| Bukhari et al ^63^ | Inpatient | RCT(Randomized Controlled Trial with no blinding) | Pakistan | 41 | 45 | 86 | vitamin C, vitamin D3 , and paracetamol SOS | single dose of ivermectin 12 milligrams | vitamin C ,vitamin D3, and paracetamol SOS | Pre-print |
| Cadegiani et al^42^ | Outpatient | Observational case control | Brazil | 110 | 137 | 247 | Supportive care | Ivermectin: 0.2mg/Kg OD for 3 days | Azithromycin with Dutasteride/ Spironolactone / Vitamin D/ Vitamin C/ Zinc/ Apixaban/ Rivaroxaban/ Enoxaparin/ glucocorticoids as per clinical judgement. | Pre-print |
| Camprubi et al ^63^ | Inpatient | Retrospective observational case control | Spain | 13 | 13 | 26 | Azithromycin, HCQ, Lopinavir/ritonavir, Tocilizumab/steroids/anakinra/siltuximab/remdesivir/b-interferon | Ivermectin 200microg/kg single dose | Azithromycin, HCQ, Lopinavir/Ritonavir, +/- Tocilizumab, Steroids, Anakinra, Remdisivir, b Interferon | Peer-reviewed |
| Carvallo1 et al^65^ | Outpatient (135) & Inpatient (32) | Clinical Trial(prospective, single center) | Argentina | 167 | - | 167 | - | Ivermectin 0.6mg/ml solution,  mild: 24 mg on day 0 and 7 moderate: 36 mg on day 0 and 7 severe: 48 mg via gastric cannulae on day 0 and 7 | mild: aspirin   moderate: dexamethasone , aspirin   severe: Dexamethasone, Enoxaparin | Pre-print |
| Chaccour et al^66^ | Outpatient | RCT(pilot, double blinded, placebo-controlled, single center, parallel arm, randomized clinical trial) | Spain | 12 | 12 | 24 | Placebo | Ivermectin at 400ug/Kg single oral dose | - | Peer-reviewed |
| Chachar et al^67^ | Outpatient | RCT(open label randomized control trial) | Pakistan | 25 | 25 | 50 | Symptomatic treatment only | Ivermectin 12mg stat, 12mg after 12 hrs, and 12mg after 24 hrs | symptomatic treatment | Peer-reviewed |
| Chahla et al^68^ |  | RCT | Argentina | 110 | 62 | 172 | symptomatic treatment: paracetamol, aspirin, Ranitidine | Ivermectin orally 4 drops of 6 mg = 24 mg every 7 days for 4 weeks | symptomatic treatment: paracetamol, aspirin, Ranitidine | Pre-print |
| Chowdhary et al^35^ | Outpatient | RCT(Randomized prospective trial, observational) | Bangladesh | 60 | 56 | 116 | Hydroxychloroquine, Azithromycin, symptomatic treatment | Ivermectin 200μgm/kg single dose | Doxycycline, symptomatic treatment | Peer-reviewed |
| Elalfy et al^69^ | Outpatient | Non-randomized controlled trial | Egypt | 62 | 51 | 113 | Paracetamol, Zinc, good nutrition and hydration; Azithromycin as needed. | Ivermectin : <60 kg or 60-90 kg:(200-300 ug /kg); 90-120 kg :(300-400 ug/kg); >120 kg:30 mg fixed dose | Nitazoxanide, Ribavirin, Zinc | Peer-reviewed |
| Espitia-Hernandez et al^70^ | Outpatient | Trial (non randomized, open label, proof of concept study) | Mexico | 28 | 7 | 35 | Standard treatment (self-isolation, proper nutrition, oral hydration and acetaminophen) | Ivermectin (6 mg once daily in day 0,1,7 and 8) | Azithromycin, Cholecalciferol | Peer-reviewed |
| Galan et al^71^ | Inpatient | RCT (double blinded randomized clinical trial) | Brazil | 53 | 115 | 168 | Chloroquine/Hydroxychloroquine; Corticosteroids, Vasoactive drugs, Anticoagulants | Ivermectin: 14mg daily x 3 days  < 55kg: 10mg each dose. | Corticosteroids, Vasoactive drugs, Anticoagulants | Peer- reviewed |
| Ghooi et al^46^ | Inpatient | Cohort study | India | 15 | 1020 | 1035 | Vit C, Enoxaparin. HCQ, Cholecalciferol, Azithromycin, Doxycycline, Remdesivir, ASA, Atorvastatin, iodine, Meropenem, Metformin, TCZ, Heparin | Ivermectin | Vit C, Enoxaparin. HCQ, Cholecalciferol, Azithromycin, Doxycycline, Remdesivir, ASA, Atorvastatin, iodine, Meropenem, Metformin, TCZ, Heparin | Pre-print |
| Gonzalez et al^72^ | Inpatient | RCT- (double blinded randomized clinical trial) | Mexico | 36 | 70 | 106 | Group 1: Hydroxychloroquine, Group 3: Placebo: Calcium citrate,  low molecular weight heparin or unfractionated heparin,  Dexamethasone in patients receiving oxygen | Ivermectin: <80Kg:12 mg, >80Kg: 18 mg | low molecular weight heparin or unfractionated heparin, Dexamethasone in patients receiving oxygen, antibiotic | Pre-print |
| Gorial et al^32^ | Inpatient | Pilot clinical trial(not randomized, interventional, single center, with a synthetic controlled arm) | Iraq | 16 | 71 | 87 | HCQ plus AZT | IVM 200 Mcg single dose | HCQ 400mg BID for the first day then 200mg BID for 5 days plus AZT 500mg single dose in the first day then 250mg for 5 days. | Pre-print |
| Guzman et al^73^ | Inpatient | Retrospective Cohort Analysis | Mexico | 122 | 74 | 196 | Dexamethasone, Antibiotics, Plasma, IVIg, low molecular weight Heparin or unfractionated Heparin, Fentanyl, Propofol, Thiopental, Midazolam, Buprenorphine, Etomidate, Ketamine, Dexmedetomidine, Remifentanil, Vecuronium, Cisatracurium, Rocuronium, Atracurium | Ivermectin: <80Kg:12 mg, >80Kg: 18 mg | Dexamethasone, Antibiotics, Plasma, IVIg, low molecular weight Heparin or unfractionated Heparin, Fentanyl, Propofol, Thiopental, Midazolam, Buprenorphine, Etomidate, Ketamine, Dexmedetomidine, Remifentanil, Vecuronium, Cisatracurium, Rocuronium, Atracurium | Pre-print |
| Hashim et al^74^ | Inpatient(22)&Outpatient(48) | RCT(randomized, Interventional, controlled single blinded) | Iraq | 70 | 70 | 140 | SOC (Acetaminophen as needed, Vitamin C, Zinc, Vitamin D3, Azithromycin, Oxygen therapy/ C-Pap if needed, Dexamethasone or Methylprednisolone if needed | Ivermectin 200ug/kg PO per day for two days, and in some patients, who needed more time to recover, a third dose 200ug/kg PO per day was given 7 days after the first dose. | Doxycycline and SOC as needed | Pre-print |
| Hazan et al^97^ | outpatient | Open Label Trial matched with Real-World Care | USA | 24 | 0 | 24 | Externally Controlled Trial (ECT) Arm | IVM (12mg on day 1, day 4, and day 8) | Doxycycline (100mg twice a day), Zinc (25mg twice a  day), Vitamin D3 (1500 IU twice a day) and Vitamin C (1500mg twice a day) | Pre-print |
| Hussain et al^75^ | Unknown | Case series | Bangladesh | 8 | - | 8 | - | Ivermectin (12 mg for patients with 80 kg weight And 18 mg for those with above 80 kg weight) single dose | Doxycycline | Peer-reviewed |
| Huvemec group^76^ | Inpatient | RCT(RCT(double blinded, multicenter)) | Bulgaria | 50 | 50 | 100 | Placebo | Ivermectin 400 μg / kg for 3 consecutive days | - | - |
| Khan et al^77^ | Inpatient | Retrospective case control study | Bangladesh | 115 | 133 | 248 | SOC(antipyretics, antihistaminics, antibiotics) | IVM one dose of 12mg | SOC | Peer-reviewed |
| Kirti et al^78^ | Inpatient | RCT(parallel, randomized, double blinded, placebo controlled trial) | India | 55 | 57 | 112 | SOC and placebo | Ivermectin 12 mg on days 1 and 2 | SOC | Pre-print |
| Kishoria et al^79^ | Inpatient | RCT(randomized, open label study) | India | 19 | 13 | 32 | Hydroxychloroquine, Paracetamol as required, Vitamin C | Ivermectin 12 mg single dose | HCQ, Paracetamol, Vitamin C | Peer-reviewed |
| Kroleweiki et al^80^ | Inpatient | RCT(pilot, randomized, open label, controlled trial) | Argentina | 30 | 15 | 45 | SOC | IVM 0·6 mg/kg/day orally for 5 days | SOC | Pre-print |
| Lima-Morales et al^43^ | Outpatient | Trial (non-randomized) | Mexico | 434 | 287 | 721 | NSAIDs, Antibiotics, Antivirals, Corticosteroids | Ivermectin 12mg single dose | Azithromycin, Montelukast, Acetylsalicylic . Some took NSAIDs, Antibiotics, Antivirals | Peer-reviewed |
| Lopez-Medina et al^81^ | Outpatient & Inpatient | RCT (double blinded randomized controlled trial) | Colombia | 200 | 198 | 398 | Placebo and NSAIDS, Macrolides, Antipyretics, non-Macrolide Antibiotics, Glucocorticoids, Anticoagulants, other Immunomodulating agents (oral Interferon or Colchicine), Acyclovir, Antidiarrheals, Antiemetics, Antihistamines, Antiparasitics, Antispasmodics, Antitussives, PPIs, Salbutamol | Ivermectin: 300ug/kg for 5 days | NSAIDS, Macrolides, Antipyretics, non-Macrolide Antibiotics, Glucocorticoids, Anticoagulants, other Immunomodulating agents (oral Interferon or Colchicine), Acyclovir, Antidiarrheals, Antiemetics, Antihistamines, Antiparasitics, Antispasmodics, Antitussives, PPIs, Salbutamol | Peer-reviewed |
| Loue et al ^82^ | outpatient | case-control study | France | 10 | 15 | 25 | Prophylactic anticoagulation, antibiotics for lung involvement, oxygen as needed | Ivermectin single dose of 200 micro grams/kg | Prophylactic anticoagulation, antibiotics for lung involvement, oxygen as needed | Peer-reviewed |
| Mahmud et al^45^ | Outpatient & Inpatient | RCT(randomized, double blinded, placebo controlled phase 3 trial) | Bangladesh | 183 | 180 | 363 | SOC(Paracetamol, Vitamin D, Oxygen if indicated, Low molecular weight Heparin, Dexamethasone if indicated) | Ivermectin 6 mg 2 tab stat | Doxycycline and SOC | Peer-reviewed |
| Mohan et al^83^ | Inpatient | RCT(double blinded, randomized, placebo controlled trial) | India | 80 | 45 | 125 | placebo +Standard hospital protocol | I arm: 12mg Ivermectin elixir; II arm: 24 mg Ivermectin elixir | Standard hospital protocol | Pre-print |
| Morgenstern et al^84^ | Outpatient(2706) & Inpatient(393) | Observational (descriptive retrospective case series) | Dominican Republic | 3099 | - | 3099 | - | outpatients: Ivermectin at 0.4mg / kg, orally  Hospitalized :Ivermectin PO at 0.3mg / kg, days 1,2,6,7. | outpatients: Azithromycin  Hospitalized : Azithromycin; Enoxaparin and Dexamethasone as needed. | Peer-reviewed |
| Mourya et al^85^ | unknown | Observational cohort study | India | 50 | 50 | 100 | HCQ, Azithromycin | Ivermectin 12 mg once a day for 7 days. | HCQ, Azithromycin | Peer-reviewed |
| Niaee et al^44^ | Inpatient | RCT(randomized, double-blind, placebo-controlled, multicenter, phase 2 clinical trial) | Iran | 60 | 120 | 180 | arm1:Hydroxychloroquine; arm2: placebo plus common regime (HCQ, heparin, supplemental oxygen) | 4arms: 1) single dose Ivermectin (200mcg/Kg, 1 pill per day), 2)three low interval doses of Ivermectin (200, 200, 200 mcg/Kg , 3 pills in 1, 3 and 5 interval days ), 3)single dose Ivermectin (400mcg/Kg, 2 pills per day), 4) three high interval doses of Ivermectin ( 400, 200, 200 mcg/Kg, 4 pills in 1, 3 and 5 interval days). | HCQ, Heparin, Oxygen, Enoxaparin and Dexamethasone as needed. | Pre-print |
| Nunez et al^86^ | Outpatient(24) & Inpatient(83) | Observational (descriptive prospective case series) | Mexico | 107 | - | 107 | - | Outpatients: Ivermectin 12 mg V.O; Repeat doses: 12 mg v.o at 48 hours;  Severe hospitalized patients: Ivermectin: 12 mg V.O. days 1, 3 and 5; Critical hospitalized patients: Ivermectin: 12 mg SNG, days 1, 3 and 5 | outpatients: Acetaminophen, Ketorolac, Atorvastatin; Severe: Azithromycin, Oseltamivir, Acetaminophen, Omeprazole Critical hospitalized patients: Atorvastatin, Azithromycin, Oseltamivir, Enoxaparin, Narcotic, sedation, Muscle relaxant | Peer-reviewed |
| Okumus et al ^88^ | Inpatient | RCT(randomized, open label, controlled phase 3 trial) | Turkey | 30 | 30 | 60 | Hydroxychloroquine, Favipiravir, Azithromycin | Ivermectin 200 mcg/kg/day for five days (9 mg between 36-50 kg, 12 mg between 51-65 kg, 15 mg between 66-79 kg and 200 microgram/kg in > 80 kg) in the form of a solution prepared for enteral use. | Azithromycin, Favipiravir, Hydroxychloroquine | Pre-print |
| Podder et al^87^ | Outpatient | RCT (randomized open-label trial) | Bangladesh | 32 | 30 | 62 | antipyretics, cough suppressants, and doxycycline | Ivermectin 200 micrograms/kg single dose was administered orally | antipyretics, cough suppressants, and doxycycline | Peer-reviewed |
| Pott-Junior et. al^89^ | Inpatient | RCT (randomized open-label trial) | Brazil | 27 | 4 | 31 | SOC, Low-molecular-weight heparin, Antibiotics, Glucocorticoids | Ivermectin: Group B=100 mcg/kg; Group C= 200 mcg/kg; Group D= 400 mcg/kg. | SOC, Low-molecular-weight heparin, Antibiotics, Glucocorticoids | Peer-reviewed |
| Rahman et al^90^ | Inpatient | prospective comparative cohort | Bangladesh | 200 | 200 | 400 | HCQ, Azithromycin | Ivermectin 18 mg on first day | Doxycycline | Peer-reviewed |
| Rajter et al^33^ | Inpatient | Retrospective Cohort study (multi-center) | USA | 173 | 107 | 280 | Hydroxychloroquine, Azithromycin or other medications | Ivermectin at 200 mcg/kg (at least one oral dose ) | HCQ/Azithromycin | Peer- reviewed |
| Samaha et al^47^ | Outpatient | RCT | Lebanon | 50 | 50 | 100 | zinc and vitamin C supplements. | Ivermectin-9 mg PO 45kg <body weight > 64kg, 12mg PO if 65kg <body weight > 84kg and 150mcg/kg if body weight ≥ 85 Kg. | zinc and vitamin C supplements. | Peer- reviewed |
| Shahbaznejad et al^91^ | Inpatient | RCT | Iran | 35 | 34 | 69 | HCQ and/or lopinavir/ritonavir, antibiotics | single oral dose of ivermectin (0.2mg/kg) | HCQ and/or lopinavir/ritonavir, antibiotics | Peer- reviewed |
| Soto-Becerra et al^92^ | Inpatient | Retrospective Cohort study | Peru | 561 | 5122 | 5683 | 1)HCQ alone 2)Azithromycin alone 3)HCQ +Azithromycin | Ivermectin 200mcg/Kg orally | One arm: Standard therapy(antipyretics, hydration) second arm: Standard therapy plus Azithromycin | Pre-print |
| Vallejos et al^96^ | outpatient | randomized, double-blind, placebo-controlled | Argentina | 250 | 251 | 501 | placebo plus standard of care (SOC) | ivermectin weighing up to 80 Kg received 2 tablets of 6 mg (mg) each at inclusion and another 2 tablets of 6 mg each 24 h after the first dose (total 24 mg). Those weighing more than 80 kg and up to 110 kg received 3 tablets of 6 mg each at inclusion and another 3 tablets of 6 mg each 24 h after the first dose (total 36 mg). Those  weighing more than 110 kg received 4 tablets of 6 mg  each at inclusion and another 4 tablets of 6 mg each 24 h after the first dose (total 48 mg). | SOC | Peer- reviewed |
| Veerapaneni et al^93^ | Inpatient | Observational Prospective study, case control | India | 50 | 50 | 100 | Placebo(Vitamin B6) | Ivermectin 200ug/kg single dose | Doxycycline | Peer- reviewed |
