## Supplemental Table 2 for "An Updated Systematic Review and Meta-Analysis of Mortality, Need for ICU admission, Use of Mechanical Ventilation, Adverse effects and other Clinical Outcomes of Ivermectin Treatment in COVID-19 Patients"

**e-table 2. Cochrane Risk of bias assessment of the trials**

| **Reference** | **Sequence generation risk of bias** | **Allocation concealment risk of bias** | **Selective reporting risk of bias** | **Other sources of bias risk of bias** | **Blinding participants and personnel risk of bias** | **Blinding outcome assessors’ risk of bias** | **Incomplete outcome data risk of bias** |
| --- | --- | --- | --- | --- | --- | --- | --- |
| **Ahmed *et al.^57^*** | High | Low | Low | Low | Low | Unclear | Low |
| **Babalola *et al.^60^*** | High | Low | Low | Low | Low | Unclear | Low |
| **Biber *et al^61^*** | Low | Low | Low | Low | Low | Low | High |
| **Bukhari et al.*^63^*** | High | High | Low | High | High | Unclear | Low |
| **Carvallo *et al.^65^*** | High | High | Low | Low | High | High | Low |
| **Chaccour *et al^66^*** | Low | Low | Low | Low | Low | Unclear | Low |
| **Chachar *et al.^67^*** | High | High | Low | Low | High | Unclear | Low |
| **Chala *et al^68^*** | High | High | Low | Low | High | Low | High |
| **Chowdhury *et al.^35^*** | Low | High | Low | Low | High | Unclear | Low |
| **Elalfy et al*^69^*** | High | High | High | Low | High | High | Low |
| **Espitia-Hernandez *et al.^70^*** | High | High | Low | Low | High | High | Low |
| **Galan et al*^71^*** | Low | Low | Low | Low | Low | Unclear | Low |
| **Gonzalez et al*^72^*** | High | Unclear | Low | Low | Low | Unclear | Low |
| **Gorial *et al. ^32^*** | High | High | Low | Low | High | Unclear | Low |
| **Hashim *et al. ^74^*** | High | High | Low | Low | High | Unclear | Low |
| **Huvemec trial*^76^*** | Unclear | Low | Unclear | Unclear | Low | Unclear | Unclear |
| **Krolewiecki *et al.^80^*** | Low | High | Low | Low | High | Low | Low |
| **Kishoria *et al.^79^*** | Low | Low | Low | Low | High | Unclear | Low |
| **Lima-Morales et al.*^43^*** | High | High | Low | Low | High | High | Low |
| **Lopez-Medina et al.*^81^*** | Low | Low | Low | Low | Low | Unclear | Low |
| **Mahmud *et al.^45^*** | Low | Low | Low | Low | Low | Unclear | Low |
| **Mohan *et al.^83^*** | Low | Low | Low | Low | Low | Low | Low |
| **Niaee *et al. ^44^*** | Low | Low | Low | Low | Low | Unclear | Low |
| **Okumus *et al. ^88^*** | High | High | Low | Low | High | Unclear | Low |
| **Podder *et al^87^*** | Low | High | Low | Low | High | Unclear | Low |
| **Pott-Junior *et al.^89^*** | Low | High | Low | Low | High | High | Low |
| **Ravikirti *et al.^78^*** | Low | Low | Low | Low | Low | Unclear | Low |
| **Samaha *et al^47^*** | High | High | Low | Low | High | Unclear | Low |
| **Shahbaznejad *et al^91^*** | Unclear | Low | Low | Low | Low | Unclear | Low |
| **Abd‐Elsalam et al^94^** | Low | High | Low | Low | High | High | Low |
| **Aref et al^95^** | High | High | Low | Unclear | High | High | Low |
| **Vallejos et al^96^** | Low | Low | Low | Low | Low | Low | Low |
| **Hazan et al^97^** | High | High | High | High | High | High | Low |
