## Supplemental Table 3 for "An Updated Systematic Review and Meta-Analysis of Mortality, Need for ICU admission, Use of Mechanical Ventilation, Adverse effects and other Clinical Outcomes of Ivermectin Treatment in COVID-19 Patients"

**e-table 3. Correlation of quality measures with estimates of treatment effects assessment of the trials**

| **Reference** | **Ahmed *et al.^57^*** | **Babalola *et al.^60^*** | **Biber *et al^61^*** | **Bukhari *et al^63^*** | **Carvallo *et al.^65^*** | **Chaccour *et al.^66^*** | **Chachar *et al.^67^*** | **Chahla *et al.^68^*** | **Chowdhury *et al.^35^*** | **Elalfy et al*^69^*** |
| --- | --- | --- | --- | --- | --- | --- | --- | --- | --- | --- |
| Study question well defined in introduction /methods | Yes | Yes | Yes | Yes | Yes | Yes | Yes | Yes | Yes | Yes |
| Study question well defined anywhere in the article | Yes | Yes | Yes | Yes | Yes | Yes | Yes | Yes | Yes | Yes |
| Placebo control | Yes | Yes | Yes | No | No | Yes | No | No | No | No |
| Appropriate outcome studied | Yes | Yes | Yes | Yes | Yes | Yes | Yes | Yes | Yes | Yes |
| Multicenter study | N/D | N/D | Yes | No | No | No | No | Yes | No | No |
| Study country | Bangladesh | Nigeria | Israel | Pakistan | Argentina | Spain | Pakistan | Argentina | Bangladesh | Egypt |
| Adequate selection criteria | Yes | No | Yes | Yes | Yes | Yes | Yes | Yes | Yes | No |
| Randomization methods described | No | No | Yes | Yes | N/A | Yes | Yes | Yes | Yes | N/A |
| Central randomization site | N/D | N/D | Yes | N/A | N/A | Yes | N/A | Yes | N/A | N/A |
| Allocation concealment | Yes | Yes | Yes | No | N/A | Yes | No | No | No | No |
| Patients blinded | Yes | Yes | Yes | No | No | Yes | No | No | No | No |
| Caregivers blinded | Yes | Yes | Yes | No | No | Yes | No | No | No | No |
| Outcome assessors blinded | N/D | N/D | Yes | No | No | N/D | N/D | No | N/D | N/D |
| Data analysts blinded | N/D | N/D | Yes | N/D | No | N/D | N/D | Yes | N/D | N/D |
| Double blinded | Yes | Yes | Yes | No | No | Yes | No | No | No | No |
| Vital statistical measures | Yes | Yes | Yes | Yes | Yes | Yes | Yes | Yes | Yes | Yes |
| Statistician author or acknowledged | No | No | Yes | No | No | No | No | Yes | Yes | Yes |
| Intention-to-treat analysis | No | No | No | No | No | N/A | No | No | No | No |
| Power calculation reported | No | No | Yes | No | No | Yes | No | No | No | No |
| Stopping rules described | No | No | No | No | No | No | No | No | No | No |
| Baseline characteristics reported | Yes | Yes | Yes | Yes | Yes | Yes | Yes | Yes | Yes | Yes |
| Groups similar at baseline | Yes | No | Yes | N/D | N/A | No | No | No | No | No |
| Confounders accounted for | No | Yes | No | No | No | Yes | No | Yes | No | No |
| Percentage dropouts | Yes | Yes | No | Yes | N/A | Yes | N/A | Yes | N/A | Yes |
| Reasons for dropout given | Yes | Yes | No | Yes | N/A | N/A | N/A | N/A | N/A | No |
| Findings support conclusion | Yes | Yes | Yes | Yes | Yes | Yes | Yes | Yes | Yes | Yes |

**N/A, Not applicable; N/D, Not defined**

| **Reference** | **Espitia-Hernandez *et al.^70^*** | **Galan et al*^71^*** | **Gonzalez et al*^72^*** | **Gorial *et al. ^32^*** | **Hashim *et al. ^74^*** | **Huvemec trial*^76^*** | **Krolewiecki *et al.^80^*** | **Kishoria *et al.^79^*** |
| --- | --- | --- | --- | --- | --- | --- | --- | --- |
| Study question well defined in introduction /methods | Yes | Yes | Yes | Yes | Yes | N/A | Yes | Yes |
| Study question well defined anywhere in the article | Yes | Yes | Yes | Yes | Yes | N/A | Yes | Yes |
| Placebo control | No | No | Yes | No | No | Yes | No | No |
| Appropriate outcome studied | Yes | Yes | Yes | Yes | Yes | Yes | Yes | Yes |
| Multicenter study | No | No | No | No | No | Yes | Yes | No |
| Study country | Mexico | Brazil | Mexico | Iraq | Iraq | Bulgaria | Argentina | India |
| Adequate selection criteria | Yes | Yes | Yes | Yes | Yes | N/D | Yes | Yes |
| Randomization methods described | Yes | Yes | Yes | N/D | Yes | No | Yes | Yes |
| Central randomization site | N/A | N/A | N/A | N/A | N/A | N/D | Yes | N/A |
| Allocation concealment | No | Yes | N/D | No | No | Yes | No | Yes |
| Patients blinded | No | Yes | Yes | No | No | Yes | No | No |
| Caregivers blinded | No | Yes | Yes | No | No | Yes | No | No |
| Outcome assessors blinded | No | N/D | N/D | N/D | N/D | N/D | Yes | N/D |
| Data analysts blinded | No | N/D | N/D | N/D | N/D | N/D | N/D | N/D |
| Double blinded | No | Yes | Yes | No | No | Yes | No | No |
| Vital statistical measures | Yes | Yes | Yes | Yes | Yes | No | Yes | Yes |
| Statistician author or acknowledged | No | No | No | No | No | N/A | Yes | No |
| Intention-to-treat analysis | No | Yes | No | No | No | N/D | Yes | No |
| Power calculation reported | No | Yes | Yes | Yes | No | No | Yes | No |
| Stopping rules described | No | No | No | No | No | No | No | Yes |
| Baseline characteristics reported | Yes | Yes | Yes | Yes | Yes | No | Yes | Yes |
| Groups similar at baseline | No | No | No | Yes | Yes | N/D | No | No |
| Confounders accounted for | No | No | No | Yes | Yes | N/D | Yes | No |
| Percentage dropouts | N/A | Yes | N/A | N/A | N/A | No | Yes | Yes |
| Reasons for dropout given | N/A | Yes | N/A | N/A | N/A | No | Yes | N/A |
| Findings support conclusion | Yes | Yes | Yes | Yes | Yes | N/A | Yes | Yes |

**e-table 3. Correlation of quality measures with estimates of treatment effects assessment of the trials continued**

**N/A, Not applicable; N/D, Not defined**

**e-table 3. Correlation of quality measures with estimates of treatment effects assessment of the trials continued**

| **Reference** | **Lima Morales *et al. ^43^*** | **Lopez-Medina *et al.^81^*** | **Mahmud *et al. ^45^*** | **Mohan *et al.^83^*** | **Niaee *et al. ^44^*** | **Okumus *et al. ^88^*** | **Podder et al^87^** | **Pott-Junior *et al.^89^*** | **Ravikirti *et al.^78^*** | **Samaha et al^47^** | **Shahbaznejad *et al.^91^*** |
| --- | --- | --- | --- | --- | --- | --- | --- | --- | --- | --- | --- |
| Study question well defined in introduction /methods | Yes | Yes | Yes | Yes | No | Yes | Yes | Yes | Yes | Yes | Yes |
| Study question well defined anywhere in the article | Yes | Yes | Yes | Yes | Yes | N/A | Yes | Yes | Yes | Yes | Yes |
| Placebo control | No | Yes | Yes | Yes | Yes | No | No | No | Yes | Yes | N/D |
| Appropriate outcome studied | Yes | Yes | Yes | Yes | Yes | Yes | Yes | Yes | Yes | Yes | Yes |
| Multicenter study | Yes* | No | No | No | Yes | Yes | No | No | No | N/D | Yes |
| Study country | Mexico | Colombia | Bangladesh | India | Iran | Turkey | Bangladesh | Brazil | India | Lebanon | Iran |
| Adequate selection criteria | Yes | Yes | Yes | Yes | Yes | Yes | Yes | Yes | Yes | Yes | Yes |
| Randomization methods described | N/A | Yes | Yes | Yes | Yes | Yes | Yes | Yes | Yes | Yes | Yes |
| Central randomization site | N/A | N/A | N/A | N/A | Yes | No | N/A | N/A | N/A | N/D | N/D |
| Allocation concealment | No | Yes | Yes | Yes | Yes | No | No | No | Yes | No | Yes |
| Patients blinded | No | Yes | Yes | Yes | Yes | No | No | No | Yes | No | Yes |
| Caregivers blinded | No | Yes | Yes | Yes | Yes | No | No | No | Yes | N/D | Yes |
| Outcome assessors blinded | N/D | N/D | N/D | Yes | N/D | N/D | N/D | N/D | N/D | N/D | N/D |
| Data analysts blinded | N/D | N/D | N/D | Yes | N/D | N/D | N/D | N/D | N/D | N/D | N/D |
| Double blinded | No | Yes | Yes | Yes | Yes | No | No | No | Yes | No | Yes |
| Vital statistical measures | Yes | Yes | Yes | Yes | Yes | Yes | Yes | Yes | Yes | Yes | Yes |
| Statistician author or acknowledged | No | Yes | No | No | No | No | No | No | Yes | No | Yes |
| Intention-to-treat analysis | No | No | Yes | Yes | Yes | No | Yes | No | No | No | No |
| Power calculation reported | No | Yes | Yes | No | Yes | Yes | No | No | Yes | Yes | No |
| Stopping rules described | No | Yes | No | No | No | No | No | No | No | No | No |
| Baseline characteristics reported | Yes | Yes | Yes | Yes | Yes | Yes | Yes | Yes | Yes | Yes | Yes |
| Groups similar at baseline | No | No | No | Yes | N/D | No | Yes | No | Yes | Yes | No |
| Confounders accounted for | Yes | Yes | N/D | Yes | N/D | N/D | N/D | No | No | Yes | No |
| Percentage dropouts | Yes | N/A | Yes | Yes | N/A | Yes | N/A | Yes | Yes | N/A | Yes |
| Reasons for dropout given | Yes | N/A | Yes | Yes | N/A | No | N/A | No | Yes | N/A | No |
| Findings support conclusion | Yes | Yes | N/A | Yes | Yes | N/A | Yes | Yes | Yes | Yes | Yes |

**N/A, Not applicable; N/D, Not defined**

**e-table 3. Correlation of quality measures with estimates of treatment effects assessment of the trials continued**

| **Reference** | **Abd‐Elsalam *et al^94^*** | **Aref *et al^95^*** | **Vallejos *et al^96^*** | **Hazan *et al^97^*** |
| --- | --- | --- | --- | --- |
| Study question well defined in introduction /methods | Yes | Yes | Yes | Yes |
| Study question well defined anywhere in the article | Yes | Yes | Yes | Yes |
| Placebo control | No | No | Yes | No |
| Appropriate outcome studied | Yes | Yes | Yes | Yes |
| Multicenter study | Yes | No | Yes | No |
| Study country | Egypt | Egypt | Argentina | USA |
| Adequate selection criteria | Yes | Yes | Yes | Yes |
| Randomization methods described | Yes | No | Yes | N/A |
| Central randomization site | No | N/A | Yes | N/A |
| Allocation concealment | Yes | N/D | Yes | No |
| Patients blinded | No | No | Yes | No |
| Caregivers blinded | No | No | Yes | No |
| Outcome assessors blinded | N/D | No | Yes | N/D |
| Data analysts blinded | N/D | N/D | Yes | N/D |
| Double blinded | No | No | Yes | No |
| Vital statistical measures | Yes | Yes | Yes | Yes |
| Statistician author or acknowledged | Yes | No | Yes | No |
| Intention-to-treat analysis | Yes | N/D | Yes | No |
| Power calculation reported | Yes | No | Yes | No |
| Stopping rules described | No | No | Yes | No |
| Baseline characteristics reported | Yes | Yes | Yes | Yes |
| Groups similar at baseline | Yes | No | Yes | N/A |
| Confounders accounted for | N/D | N/D | Yes | No |
| Percentage dropouts | Yes | Yes | Yes | Yes |
| Reasons for dropout given | N/A | N/A | N/A | Yes |
| Findings support conclusion | Yes | Yes | Yes | Yes |

**N/A, Not applicable; N/D, Not defined**
