## Supplemental Table 4 for "An Updated Systematic Review and Meta-Analysis of Mortality, Need for ICU admission, Use of Mechanical Ventilation, Adverse effects and other Clinical Outcomes of Ivermectin Treatment in COVID-19 Patients"

**e-table 4. NIH quality assessment Tool for case series**

| **Reference** | **Alam *et al.^58^*** | **Bhattacharya *et al.^34^*** | **Budhiraja *et al.^61^*** | **Hussain *et al^75^*** | **Morgenstern *et al^84^*** | **Nunez *et al.^86^*** | **Ahsan *et al^57^*** |
| --- | --- | --- | --- | --- | --- | --- | --- |
| 1. Was the study question or objective clearly stated? | Yes | Yes | Yes | Yes | Yes | Yes | Yes |
| 2. Was the study population clearly and fully described, including a case definition? | Yes | Yes | Yes | Yes | Yes | Yes | Yes |
| 3. Were the cases consecutive? | Yes | Yes | Yes | Yes | Yes | Yes | Yes |
| 4. Were the subjects comparable? | N/D | N/A | N/D | N/A | Yes | Yes | No |
| 5. Was the intervention clearly described? | Yes | Yes | No | Yes | Yes | Yes | Yes |
| 6. Were the outcome measures clearly defined, valid, reliable and implemented consistently across all study participants? | Yes | Yes | Yes | Yes | Yes | Yes | No |
| 7. Was the length of follow-up adequate? | No | No | No | No | N/D | Yes | Yes |
| 8. Were the statistical methods well described? | N/A | Yes | Yes | N/A | N/A | No | Yes |
| 9. Were the results well described? | Yes | Yes | Yes | Yes | Yes | Yes | Yes |
| **Quality rating** | **Fair** | **Good** | **Fair** | **Fair** | **Good** | **Good** | **Fair** |

**N/A: Not applicable; N/D, Not defined; CD:cannot determine**
