## Supplemental Table 5 for "An Updated Systematic Review and Meta-Analysis of Mortality, Need for ICU admission, Use of Mechanical Ventilation, Adverse effects and other Clinical Outcomes of Ivermectin Treatment in COVID-19 Patients"

**e-table 5. NIH Quality Assessment of Case-Control Studies**

| **Criteria** | **Afsar et al*^55^*** | **Cadegiani et al*^42^*** | **Camprubi et al*^63^*** | **Khan et al*^77^*** | **Loue *et al^82^*** | **Veerapaneni et al*^93^*** |
| --- | --- | --- | --- | --- | --- | --- |
| **1. Was the research question or objective in this paper clearly stated and appropriate?** | Yes | Yes | Yes | Yes | No | Yes |
| **2. Was the study population clearly specified and defined?** | Yes | Yes | Yes | Yes | Yes | No |
| **3. Did the authors include a sample size justification?** | No | Yes | No | No | No | No |
| **4. Were controls selected or recruited from the same or similar population that gave rise to the cases (including the same timeframe)?** | Yes | Yes | No | Yes | Yes | Yes |
| **5. Were the definitions, inclusion and exclusion criteria, algorithms or processes used to identify or select cases and controls valid, reliable, and implemented consistently across all study participants?** | Yes | Yes | No | No | No | Yes |
| **6. Were the cases clearly defined and differentiated from controls?** | Yes | CD | Yes | Yes | Yes | Yes |
| **7. If less than 100 percent of eligible cases and/or controls were selected for the study, were the cases and/or controls randomly selected from those eligible?** | NA | NA | NA | NR | NA | No |
| **8. Was there use of concurrent controls?** | CD | NR | NR | NR | Yes | No |
| **9. Were the investigators able to confirm that the exposure/risk occurred prior to the development of the condition or event that defined a participant as a case?** | Yes | Yes | Yes | Yes | Yes | Yes |
| **10. Were the measures of exposure/risk clearly defined, valid, reliable, and implemented consistently (including the same time period) across all study participants?** | Yes | No | Yes | Yes | Yes | Yes |
| **11. Were the assessors of exposure/risk blinded to the case or control status of participants?** | CD | NO | NR | CD | NR | No |
| **12. Were key potential confounding variables measured and adjusted statistically in the analyses? If matching was used, did the investigators account for matching during study analysis?** | No | Yes | No | No | No | No |
| **Quality Rating (Good, Fair, or Poor)** | **Low** | **Low** | **Very Low** | **Low** | **Low** | **Low** |
| **Risk of Bias** | **Moderate** | **Moderate** | **High** | **Moderate** | **Moderate** | **Moderate** |

**NA=not applicable, CD=cannot determine,NR=not reported**
