## Supplemental Table 6 for "An Updated Systematic Review and Meta-Analysis of Mortality, Need for ICU admission, Use of Mechanical Ventilation, Adverse effects and other Clinical Outcomes of Ivermectin Treatment in COVID-19 Patients"

**e-table 6. NIH Quality Assessment of Observational Cohort and Cross-Sectional Studies**

| **Criteria** | **Ghooi *et al^46^*** | **Guzman *et al.^73^*** | **Mourya *et al.^85^*** | **Rajter *et al^33^*** | **Soto-Becerra *et al^92^*** | **Rahman *et al^90^*** |
| --- | --- | --- | --- | --- | --- | --- |
| **1. Was the research question or objective in this paper clearly stated?** | Yes | Yes | Yes | Yes | Yes | Yes |
| **2. Was the study population clearly specified and defined?** | No | Yes | Yes | Yes | Yes | Yes |
| **3. Was the participation rate of eligible persons at least 50%?** | CD | No | CD | CD | CD | NR |
| **4. Were all the subjects selected or recruited from the same or similar populations (including the same time period)? Were inclusion and exclusion criteria for being in the study prespecified and applied uniformly to all participants?** | CD | Yes | Yes | Yes | No | NR |
| **5. Was a sample size justification, power description, or variance and effect estimates provided?** | No | No | No | No | CD | No |
| **6. For the analyses in this paper, were the exposure(s) of interest measured prior to the outcome(s) being measured?** | Yes | Yes | Yes | Yes | Yes | Yes |
| **7. Was the timeframe sufficient so that one could reasonably expect to see an association between exposure and outcome if it existed?** | CD | No | NR | No | No | ND |
| **8. For exposures that can vary in amount or level, did the study examine different levels of the exposure as related to the outcome (e.g., categories of exposure, or exposure measured as continuous variable)?** | CD | No | NR | No | No | No |
| **9. Were the exposure measures (independent variables) clearly defined, valid, reliable, and implemented consistently across all study participants?** | Yes | No | Yes | Yes | No | Yes |
| **10. Was the exposure(s) assessed more than once over time?** | No | No | No | No | No | Yes |
| **11. Were the outcome measures (dependent variables) clearly defined, valid, reliable, and implemented consistently across all study participants?** | Yes | Yes | Yes | Yes | Yes | No |
| **12. Were the outcome assessors blinded to the exposure status of participants?** | No | No | No | No | No | No |
| **13. Was loss to follow-up after baseline 20% or less?** | NA | NR | Yes | Yes | NR | Yes |
| **14. Were key potential confounding variables measured and adjusted statistically for their impact on the relationship between exposure(s) and outcome(s)?** | No | NO | No | Yes | Yes | No |
| **Quality Rating (Good, Fair, or Poor)** | Poor | **Very Low** | **Low** | **Low** | **Very Low** | **Very low** |
| **Risk of Bias** | High | **High** | **Moderate** | **Moderate** | **High** | **High risk of bias** |

**NR=not reported, NA=not applicable, CD=cannot determine**
