## Supplemental Table 7 for "An Updated Systematic Review and Meta-Analysis of Mortality, Need for ICU admission, Use of Mechanical Ventilation, Adverse effects and other Clinical Outcomes of Ivermectin Treatment in COVID-19 Patients"

**e-table 7. Certainty of the evidence (GRADE) Profile at Outcome Level**

| **Patient or population: COVID-19**  **Intervention: Ivermectin**  **Comparison: No Ivermectin** | | | | | | |
| --- | --- | --- | --- | --- | --- | --- |
| A. **GRADE for Mortality** | | | | | | |
| Outcomes | **Anticipated absolute effects^*^** (95% CI) | | Relative effect (95% CI) | № of participants  (studies) | Certainty of the evidence (GRADE) | Comments |
|  | **Risk with No Ivermectin** | **Risk with Ivermectin** |  |  |  |  |
| **Mortality (Overall)** | 143 per 1,000 | **83 per 1,000**  **(54 to 126)** | **OR 0.54**  **(0.34 to 0.86)** | 12647  (29 studies) | ⨁◯◯◯ VERY LOW | Downgraded for study design, risk of bias, inconsistency, and publication bias |
| **Mortality (Clinical trials)** | 75 per 1,000 | **37 per 1,000**  **(20 to 68)** | **OR 0.47**  **(0.25 to 0.90)** | 3592  (18 studies) | ⨁⨁◯◯ LOW | Downgraded for study design, risk of bias, inconsistency, and publication bias |
| **Mortality (Therapeutic RCTs Mild-Moderate COVID-19)** | 24 per 1,000 | **8 per 1,000**  **(2 to 25)** | **OR 0.31**  **(0.09 to 1.04)** | 2261  (12 studies) | ⨁⨁◯◯ LOW | Downgraded for risk of bias, mixed effect of Ivermectin and upgraded for very large magnitude of effect |
| **Mortality (Therapeutic RCTs Severe/Critical COVID-19)** | 200 per 1,000 | **177 per 1,000**  **(113 to 266)** | **OR 0.86**  **(0.51 to 1.45)** | 410  (6 studies) | ⨁⨁◯◯ LOW | Downgraded for risk of bias, mixed effect of Ivermectin |
| **Mortality (Observational)** | 158 per 1,000 | **107 per 1,000** (62 to 179) | **OR 0.64** (0.35 to 1.16) | 9055 (11 studies) | ⨁◯◯◯ VERY LOW | Downgraded for study design, risk of bias, inconsistency, and publication bias |
| **Mortality (Inpatient Overall)** | 158 per 1,000 | **109 per 1,000**  **(73 to 158)** | **OR 0.65**  **(0.42 to 1.00)** | 9825  (18 studies) | ⨁◯◯◯ VERY LOW | Downgraded for study design, risk of bias, inconsistency, and publication bias |
| **Mortality (Inpatient RCTs-Severe/Critical COVID-19)** | 65 per 1,000 | **12 per 1,000**  **(1 to 80)** | **OR 0.18**  **(0.02 to 1.25)** | 645  (5 studies) | ⨁⨁◯◯ LOW | Downgraded for risk of bias and inconsistency and upgraded for large magnitude of effect |
| **Mortality (Inpatient RCTs- Mild-Moderate COVID-19)** | 200 per 1,000 | **177 per 1,000**  **(113 to 266)** | **OR 0.86**  **(0.51 to 1.45)** | 410  (6 studies) | ⨁⨁◯◯ LOW | Downgraded for risk of bias and inconsistency and upgraded for large magnitude of effect |
| **Mortality (Observational Inpatient)** | 162 per 1,000 | **119 per 1,000** (68 to 200) | **OR 0.70** (0.38 to 1.30) | 8683 (8 studies) | ⨁◯◯◯ VERY LOW | Downgraded for study design, risk of bias, inconsistency, and publication bias |
| **Mortality (Effect of Ivermectin Monotherapy as per Cochrane Guideline with Placebo/Standard of Care RCTs: Overall** | 26 per 1,000 | 20 per 1,000 | **OR 0.75**  **(0.35 to 1.63)** | 1424  (7 studies) | ⨁⨁⨁◯ MODERATE | Downgraded for risk of bias and inconsistency and upgraded for large magnitude of effect and study design |
| **Mortality (Effect of Ivermectin alone as per Cochrane Guideline with Placebo/Standard of Care RCTs: Mild-Moderate COVID-19)** | 18 per 1,000 | **13 per 1,000**  **(5 to 34)** | **OR 0.71**  **(0.27 to 1.87)** | 1351  (6 studies) | ⨁⨁⨁◯ MODERATE | Downgraded for risk of bias and inconsistency and upgraded for large magnitude of effect and study design |
| **Mortality (Effect of Ivermectin alone as per Cochrane Guideline with Placebo/Standard of Care RCTs- Severe/Critical COVID-19)** | 162 per 1,000 | **138 per 1,000**  **(4**3 **to 3**6**9)** | **OR 0.83**  **(0.23 to 3.02)** | 73  (1 study) | ⨁⨁◯◯ LOW | Downgraded for risk of bias, small sample size and inconsistency and upgraded for study design |
| B. **GRADE for ICU admission** | | | | | | |
| **Need for ICU Admission** | 161 per 1,000 | **85 per 1,000** (32 to 209) | **OR 0.48** (0.17 to 1.37) | 585 (5 studies) | ⨁◯◯◯ VERY LOW | Downgraded for study design, risk of bias and inconsistency |
| C. **GRADE for Mechanical Ventilation** | | | | | | |
| **Need for Mechanical Ventilation** | 76 per 1,000 | **58 per 1,000** (41 to 82) | **OR 0.75** (0.52 to 1.08) | 2516 (12 studies) | ⨁◯◯◯ VERY LOW | Downgraded for study design, risk of bias, inconsistency, and publication bias |
| D. **GRADE for Adverse events** | | | | | | |
| **Adverse Event** | 210 per 1,000 | **188 per 1,000** (151 to 231) | **OR 0.87** (0.67 to 1.13) | 2777 (22 studies) | ⨁◯◯◯ VERY LOW | Downgraded for study design, risk of bias, inconsistency, imprecision, and publication bias |
| E. **Need for Hospitalization** | | | | | | |
| **Need for Hospitalization** | 224 per 1,000 | **89 per 1,000 (44 to 178)** | **OR 0.34 (0.16 to 0.75)** | 2021 (6 studies) | ⨁⨁◯◯ LOW | Downgraded for risk of bias, mixed effect of Ivermectin and upgraded for very large magnitude of effect |
| F. **Duration of Hospital Stay** | | | | | | |
| **Duration of Hospital stay** | The mean duration of Hospital stay was **0** | MD **1.8 lower** (3.69 lower to 0.08 higher) | **-** | 1246  (9 studies) | ⨁⨁◯◯ LOW | Downgraded for risk of bias, mixed effect of Ivermectin and upgraded for very large magnitude of effect |
| G. **Incidence of Viral Clearance** | | | | | | |
| **Incidence of Viral clearance** | 655 per 1,000 | **870 per 1,000** (775 to 929) | **OR 3.52** (1.81 to 6.86) | 2533 (22 studies) | ⨁⨁◯◯ LOW | Downgraded for risk of bias, mixed effect of Ivermectin and upgraded for very large magnitude of effect |
| H. **Time to Achieve Viral Clearance** | | | | | | |
| **Time to Achieve Viral Clearance** | The mean time to achieve Viral clearance was **0** | MD **4.12 lower** (7.59 lower to 0.65 lower) | **-** | 948 (8 studies) | ⨁⨁◯◯ LOW | Downgraded for risk of bias, mixed effect of Ivermectin and upgraded for very large magnitude of effect |
| ***The risk in the intervention group (and its 95% confidence interval) is based on the assumed risk in the comparison group and the relative effect of the intervention (and its 95% CI).**  **CI: Confidence interval; MD: Mean difference** | | | | | | |
| **GRADE Working Group grades of evidence**  **High certainty: We are very confident that the true effect lies close to that of the estimate of the effect**  **Moderate certainty: We are moderately confident in the effect estimate: The true effect is likely to be close to the estimate of the effect, but there is a possibility that it is substantially different**  **Low certainty: Our confidence in the effect estimate is limited: The true effect may be substantially different from the estimate of the effect**  **Very low certainty: We have very little confidence in the effect estimate: The true effect is likely to be substantially different from the estimate of effect** | | | | | | |
