## Supplemental Table 8 for "An Updated Systematic Review and Meta-Analysis of Mortality, Need for ICU admission, Use of Mechanical Ventilation, Adverse effects and other Clinical Outcomes of Ivermectin Treatment in COVID-19 Patients"

**e-table 8. Ongoing clinical trials**

| **Serial**  **number** | **Name of the trial** | **NCT number** | **Sponsor/Collaborators** | **Country** | **Study Design** | **Number enrolled** | **Interventions** | **Outcomes** | **Status** |
| --- | --- | --- | --- | --- | --- | --- | --- | --- | --- |
| 1 | Efficacy of Nano-Ivermectin Impregnated Masks in Prevention of Covid-19 Among Healthy Contacts and Medical Staff | NCT04723459 | South Valley University | Egypt | Interventional | 150 | - Ivermectin mask | - Number of persons in each group who Complain of any suspected Symptoms - Number of persons in each group who are diagnosed as COVID-19 patients | Recruiting |
| 2 | Inhaled Ivermectin and COVID-19(CCOVID-19) | NCT04681053 | Mansoura University | Egypt | Interventional | 80 | - Ivermectin Powder | - Rate of virological cure by RT-PCR - Resolution of pneumonia | Recruiting |
| 3 | Ivermectin in the treatment of COVID-19 patients | NCT04723459 | Ministry of Health and Population, Egypt | Egypt | Interventional | 100 | - Ivermectin | - Rate of viral clearance in comparison to other treatment protocols - Evaluate the role of ivermectin as a line of treatment for COVID-19 | Recruiting |
| 4 | An Outpatient Clinical Trial Using Ivermectin and Doxycycline in COVID-19 Positive Patients at High Risk to Prevent COVID-19 Related Hospitalization | NCT04729140 | Max Health, Subsero Health | USA | Interventional | 150 | - Ivermection, - Doxycycline - Placebo | - Assessment of Hb, WBC, HcT, Platelet Count, Na,Cl,Co2 levels, - Admission rate to the hospital secondary to respiratory illness related to COVID-19, - Total duration of symptoms secondary to respiratory illness related to COVID-19 | Recruiting |
| 5 | COVidIVERmectin: Ivermectin for Treatment of Covid-19 | NCT04438850 | IRCCS Sacro Cuore Don Calabria di Negrar and Istituto Di Ricerche Farmacologiche Mario Negri | Italy and Spain | Interventional | 102 | - Ivermectin - Placebo | - SADR - Viral load - Trend viral load - Clinical resolution - Viral clearance - Virological clearance - Hospitalization rate - Severity score | Recruiting |
| 6 | Ivermectin to Prevent Hospitalizations in COVID-19 | NCT04529525 | Instituto de Cardiología de Corrientes | Argentina | Interventional | 500 | - Ivermectin - Placebo | - Percentage of Hospitalization of medical cause in patients with COVID-19 in each arm - Time to hospitalization - Percentage of use of invasive mechanical support in each arm - Time to invasive mechanical ventilation support - Percentage of dialysis in each arm - All-cause mortality - Negative of the swab at 3±1 days and 12±2 days after entering the study - Incidence of Treatment Emergent Adverse Events | Recruiting |
| 7 | Study in COvid-19 Patients With ivermectin (CORVETTE-01) | NCT04703205 | Kitasato University | Japan | Interventional | 240 | - Ivermectin - Placebo | - Period until the COVID-19 PCR test becomes negative | Recruiting |
| 8 | Evaluation of Prognostic Modification in COVID-19 Patients in Early Intervention Treatment | NCT04673214 | Gilberto Cruz Arteaga | Mexico | Interventional | 62 | - Azithromycin - Ivermectin - Ribaroxaban - Paracetamol | - Estimate clinical symptoms by days of follow-up - To assess adverse drug reactions by days of follow-up | Recruiting |
| 9 | Ivermectin vs. Placebo for the Treatment of Patients With Mild to Moderate COVID-19 | NCT04429711 | Sheba Medical Center | Israel | Interventional | 100 | - Ivermectin | - Viral clearance at day 6 - Viral shedding - Duration of Symptoms - Clearance time | Recruiting |
| 10 | Evaluation of Ivermectin Mucoadhesive Nanosuspension as Nasal Spray in Management of Early Covid-19 | NCT04716569 | South Valley University | Egypt | Interventional | 150 | - Ivermectin intranasal spray | - Progression of covid 19 clinical picture | Recruiting |
| 11 | Efficacy of Ivermectin in outpatients with non-severe COVID-19 | NCT04834115 | Universidad Nacional de Asuncion | Paraguay | Interventional | 400 | - Ivermectin tablets | - Primary-proportion of patients with hospitalization criteria - Secondary- 1)proportion of patients with COVID-19 signs and symptoms 2)proportion of cohabitants who had COVID-19 after the index case 3)drug-related adverse events 4)Levels of IgG for SARS-CoV-2 | Recruiting |
| 12 | Ivermectin in Adults With Severe COVID-19. | NCT04602507 | CES University | Colombia | Interventional | 100 | - Ivermectin plus routine care | - Admission to the intensive care unit - Hospital length of stay - Mortality rate - ICU length of stay - Length of stay in ventilator time - Adverse effects of ivermectin | Recruiting |
| 13 | Ivermectin as a Novel Therapy in COVID-19 Treatment | NCT04403555 | Tanta University | Egypt | Interventional | 160 | - Ivermectin plus standard of care | - The number of patients with improvement or mortality | Recruiting |
| 14 | Ivermectin in Treatment of COVID-19 | NCT04445311 | Zagazig University | Egypt | Interventional | 100 | - Ivermectin | - Time to be symptoms free - Need hospital admission - Need mechanical ventilation - Length of stay - Mortality | Recruiting |
| 15 | Efficacy of Ivermectin in COVID-19 | NCT04392713 | Combined Military Hospital, Pakistan | Pakistan | Interventional | 100 | - Ivermectin | - Negative PCR - Need for mechanical ventilation | Recruiting |
| 16 | The Efficacy of Ivermectin and Nitazoxanide in COVID-19 Treatment | NCT04351347 | Tanta University | Egypt | Interventional | 300 | - Ivermectin plus Nitazoxanide | - Number of patients with improvement or died | Recruiting |
| 17 | Max Ivermectin- COVID 19 Study Versus Standard of Care Treatment for COVID 19 Cases. A Pilot Study | NCT04373824 | Max Healthcare Institute Limited | India | Interventional | 50 | - Ivermectin | - Effect of ivermectin on the eradication of virus | Recruiting |
| 18 | Repurposed Approved Therapies for Outpatient Treatment of Patients With Early-Onset COVID-19 and Mild Symptoms | NCT04727424 | Cardresearch | Brazil | Interventional | 2724 | - Fluvoxamine Maleate 100 MG [Luvox] - Metformin Extended Release Oral Tablet - Ivermectin Tablets - Placebo | - Change in viral load on day 03 and 07 after randomization - Time to > 50% clinical improvement - Time to hospitalization - Days with symptoms - All cause hospitalizations - COVID-19 hospitalizations - All-Cause Death - Cardiovascular death - Respiratory death - Promis Global-10 scale - WHO ordinal scale for clinical improvement - Percentage of adherence on Study drug | Recruiting |
| 19 | Ivermectin-Azithromycin-Cholecalciferol (IvAzCol) Combination Therapy for COVID-19 (IvAzCol) | NCT04399746 | Instituto de Seguridad y Servicios Sociales de los Trabajadores del Estado | Mexico | Interventional | 30 | - Ivermectin - Azithromycin - Cholecalciferol | - Viral clearance - Symptoms duration - SpO2, SpO2/FiO2 | Recruiting |
| 20 | Ivermectin vs Combined Hydroxychloroquine and Antiretroviral Drugs (ART) Among Asymptomatic COVID-19 Infection (IDRA-COVID19) | NCT04435587 | Mahidol  University | Thailand | Interventional | 80 | - Ivermectin Pill - Combined ART/ hydroxychloroquine | - Adverse event rates - Efficacy for shortening duration of SAR-CoV2 detection by PCR - Antibody detection rates | Recruiting |
| 21 | Efficacy of Subcutaneous Ivermectin With or Without Zinc in COVID-19 Patients (SIZI-COVID-PK) | NCT04472585 | Sohaib Ashraf | Pakistan | Interventional | 60 | - Ivermectin Injectable Solution - Injectable Placebo - Zinc - Placebo empty capsule | - Time needed to turn positive COVID-19 PCR to negative - Time taken for alleviation of symptoms - Severity of symptoms - Mortality | Recruiting |
| 22 | Randomized Phase IIA Clinical Trial to Evaluate the Efficacy of Ivermectin to Obtain Negative PCR Results in Patients With Early Phase COVID-19 (SAINT-PERU) | NCT04635943 | Universidad Peruana Cayetano Heredia  Barcelona Institute for Global Health | Peru | Interventional | 68 | - Ivermectin - Placebo | - Proportion of patients with a positive SARS-CoV-2 PCR - Mean viral load - Fever and cough progression - Seroconversion at day 21 - Proportion of drug-related adverse events - Levels of IgG, IgM and IgA - Frequency of innate immune cells - Frequency SARS-CoV-2- specific CD4+ and CD8+ T cells - Results from cytokine Human Magnetic 30-Plex Panel - Presence of intestinal helminths | Recruiting |
| 23 | Early Treatment With Ivermectin and LosarTAN for Cancer Patients With COVID-19 Infection (TITAN) | NCT04447235 | Instituto do Cancer do Estado de São Paulo | Brazil | Interventional | 176 | - Placebo - Ivermectin - Losartan | - Incidence of severe complications due COVID-19 infection - Incidence of Severe Acute Respiratory Syndrome - Adverse events - Overall survival | Recruiting |
| 24 | Comparative therapeutic efficacy and safety of different antiviral and anti inflammatory drugs in COVID-19 patients | NCT04779047 | Beni-Suef University | Egypt | Interventional | 150 | - Remdesivir - Hydroxychloroquine - Tocilizumab - Lopinavir/ Ritonavir - Ivermectin | - Percentage of clinical cure in each arm | Recruiting |
| 25 | Novel Agents for Treatment of High-risk COVID-19 Positive Patients | NCT04374019 | Susanne Arnold  University of Kentucky | United States | Interventional | 240 | - Ivermectin - Camostat Mesilate - Dietary Supplement: Artemesia annua, Artesunate | - Clinical Deterioration - Change in Viral Load - Rate of Organ Failure - Progression to ICU Care or Ventilation - Change in Clinical Status - Mortality - Rate of severe adverse events - Oxygen-free days - Ventilator-free days - Vasopressor-free days - ICU-free days - Hospital-free days - Patients meeting Hy's Law criteria - Liver Function - Heart Function | Recruiting |
| 26 | Comparative Study of Hydroxychloroquine and Ivermectin in COVID-19 Prophylaxis | NCT04384458 | Nucleo De Pesquisa E Desenvolvimento De Medicamentos Da Universidade Federal Do Ceara | Brazil | Interventional | 400 | - Hydroxychloroquine - Ivermectin | - Proportion of participants in whom there was a positivity for SARS CoV-2 - Participants who developed mild, moderate, or severe forms of COVID-19 - Measurement of the QT interval - Widening of the corrected QT interval or with changes in heart rate on the ECG - Comparison of hematological and biochemical parameters - Occurrence of adverse events - Assessment of COVID-19 symptom severity - Proportion of participants who discontinue study intervention - Proportion of participants who required hospital care - Proportion of participants who required mechanical ventilation. | Recruiting |
| 27 | A Study to Compare the Efficacy and Safety of Different Doses of Ivermectin for COVID-19 | NCT04431466 | Universidade Federal de Sao Carlos | Brazil | Interventional | 64 | - Ivermectin - Standard treatment for COVID-19 | - Time to undetectable SARS-CoV-2 viral load in the nasopharyngeal swab - Viral load variation in the nasopharyngeal swab - Proportion of patients with undetectable SARSCoV-2 viral load in the nasopharyngeal swab - Proportion of patients with clinical improvement. | Recruiting |
| 28 | Prevention and Treatment for COVID -19 (Severe Acute Respiratory Syndrome Coronavirus 2 SARS-CoV-2) Associated Severe Pneumonia in the Gambia | NCT04703608 | London School of Hygiene and Tropical Medicine | Gambia | Interventional | 1200 | - Ivermectin - ASP (Aspirin Parallel Assignment) - Placebo | - Cohort 1 Index Case: Percentage of patients with COVID-19 associated mild disease/moderate pneumonia progressing to severe pneumonia. - Cohort 1 Household contacts: Percentage of HH members that get infected with SARS-CoV-2 - Cohort 2: Percentage of COVID-19 associated severe pneumonia patients worsening their condition - Cohort1 Index cases: Days from recruitment to virological clearance - Days from recruitment until clinical recovery - IgG geometric mean titre (GMT) at day 14 and 28 after recruitment -Household contacts IgG geometric mean titre (GMT) at day 14 after recruitment  -Percentage of HH members infected that develop COVID19 symptoms  -Cohort 2 - Hours from recruitment to hospital discharge -Hours of duration on oxygen supplementation - Death ratio during hospitalization - Death ratio at D28 and D90 - Occurrence of clinical thrombotic and embolic events - Occurrence of clinical episodes of gastrointestinal bleeding - Change in CRP and D-Dimer levels - Persisting breathlessness at 28 days and 90 days after - Self-reported health at 28 days and 90 days | Recruiting |
| 29 | Exploratory Ph I Trial of the Active IMP in Healthy Volunteers in Relation to COVID-19 | NCT04632706 | MedinCell S.A  MAC Clinical Research | United Kingdom | Interventional (Randomized, double blinded) | 24 | - Ivermectin - Placebo | • Pharmacokinetic concentrations:  - Maximum Plasma Concentration [Cmax]  - Time to Reach Cmax [Tmax]  - Trough Plasma Concentration [Ctrough]  - Area under the plasma concentration-time curve from zero to 24 hours [AUC0-24h]  - Area under the plasma concentration-time curve from zero to 48 hours [AUC0-48h  - Average Plasma Concentration at steady state [Cavg ss]  - Apparent Terminal Half-Life [T1/2]  - Number of participants with treatment emergent adverse events (TEAEs)  •Safety and Tolerability:  - Number of participants with abnormal electrocardiograms (ECG)  - Number of participants with abnormal clinical neurological exam  - Number of participants with abnormal urine and/or blood test  - Number of participants with abnormal physical exams | Recruiting |
| 30 | Trial of Combination Therapy to Treat COVID-19 Infection | NCT04482686 | ProgenaBiome  Topelia Therapeutics | United States | Interventional | 30 | - Ivermectin - Doxycycline - Hcl - Zinc - Vitamin D3 - Vitamin C | - Time to Non-Infectivity by RT-PCR - Time to Symptom progression in days - Time to Symptom improvement - Efficacy of Treatment as measured by Titer - Efficacy of Treatment as measured by RT-PCR, D-Dimer, ProCalcitonin, CRP, Ferritin, Liver Enzymes, CBC, Electrolyte Levels - Treatment Related Adverse Events | Recruiting |
| 31 | Risk Stratification of COVID-19 Using Urine Biomarkers | NCT04681040 | National Center for Global Health and Medicine, Japan | United States, Brazil, Denmark, Japan, Tokyo, Japan, Philippines | Observational, Prospective Cohort | 1000 | - | - Risk Stratification of COVID-19 Participants Using Urine Biomarkers - Prediction of COVID-19 Treatment by Urine LFABP - Increase of O2 support, hospital days, worsening of chest X-ray and CT, and survival rate, at 14 and/or 30 days - Comparison of Risk Stratification with Other Biomarkers | Recruiting |
| 32 | Ivermectin Nasal spray for COVID-19 patients | NCT04510233 | Tanta University | Egypt | Interventional | 60 | - Ivermectin (oral and nasal) | - PCR of SARS-Cov2 RNA | Not yet recruiting |
| 33 | Outpatient use of Ivermectin | NCT04530474 | Temple University | USA | Interventional | 200 | - Ivermectin | - Clinical improvement | Not yet recruiting |
| 34 | The (HD)IVACOV Trial (The High-Dose Ivermectin Against COVID-19 Trial) | NCT04712279 | Corpometria Institute | Brazil | Interventional | 294 | - Ivermectin - Hydroxychloroquine | - WHO clinical progression scale - Clinical Improvement - Time-to-recovery - Viral load - Positivity rate of rtPCR-SARS-CoV-2 - Duration of fatigue and anosmia - Overall duration of clinical manifestations - Proportion of subjects needing - Additional drugs or interventions - oxygen use - high-flow oxygen therapy or non-invasive ventilation - Proportion of hospitalizations - Proportion of mechanical ventilation use - Proportion of pressors use - Proportion of deaths - Proportion of post-COVID mental and physical symptoms - Proportion of post-COVID overall symptoms - Duration of new oxygen use, hospitalization and mechanical ventilation - Proportion of increased CRP, d-dimer and eosinophils - Proportion of decrease in ESR - Disease duration - Change in viral load from baseline to Day 5 | Not yet recruiting |
| 35 | Ivermectin and Nitazoxanide Combination Therapy for COVID-19 | NCT04360356 | Tanta University | Egypt | Interventional | 100 | - Ivermectin plus Nitazoxanide | - Number of Patients with COVID-19-negative PCR - Number of patients with improved respiratory rate - Number of patients with improved PaO2 - Number of patients with normalized Serum IL6 - Number of patients with normalized Serum TNFα - Number of patients with normalized Serum iron - Number of patients with normalized Serum ferritin - Number of patients with normalized PT-INR - Number of patients with normalized complete blood count, - Mortality rate among treated patients | Not yet recruiting |
| 36 | Safety and Efficacy of Ivermectin and Doxycycline in Treatment of Covid-19 | NCT04551755 | Bangladesh Medical Research Council (BMRC | Bangladesh | Interventional | 188 | - Ivermectin and Doxycycline - Placebo | - Time to outcome measure of fever (<100.40F)and cough - Negative RT-PCR test on day 5 of treatment | Not yet recruiting |
| 37 | Safety and efficacy of low dose aspirin/ivermectin combination therapy for treatment of COVID-19 patients (IVCOM) | NCT04768179 | - Makarere University - Ministry of Health, Uganda - Mbarara University of Science and Technology - Joint Clinical Research Center | Uganda | Interventional | 490 | - Ivermectin - ASA - Standard of therapy | - SARS COV 2 Viral clearance - World Health Organization COVID-19 ordinal improvement score - Clinical recovery - Spectrum and severity of adverse events - Maximum Plasma concentration - Minimum Plasma concentration - Area Under the Curve | Not yet recruiting |
| 38 | Effectiveness and Safety of Ivermectin for the Prevention of Covid 19 Infection in Colombian Health Personnel | NCT04527211 | Javeriana University | Colombia | Interventional | 550 | - Ivermectin | - Clinical development of covid-19 disease during the intervention period - Seroconversion - Hospitalization requirement - Intensive Care Unit Requirement - Safety of the intervention | Not yet recruiting |
| 39 | New Antiviral Drugs for Treatment of COVID-19 | NCT04392427 | Mansoura University | Egypt | Interventional | 100 | - Nitazoxanide, Ribavirin and Ivermectin for a duration of seven days | - Negative test result for COVID-19 | Not yet recruiting |
| 40 | Study the efficacy and therapeutic safety of Ivermectin (SAINTBO) | NCT04836299 | - Universidad Mayor de San Simón - Barcelona Institute for Global Health - Université Catholique de Louvain | Bolivia | Interventional | 90 | - Ivermectin - Placebo | - Evolution of viral load - Clinical remission - Clinical signs of toxicity - Need for supplemental oxygen - Hospital stay - Need for mechanical ventilation | Not yet recruiting |
| 41 | Efficacy, Safety and Tolerability of Ivermectin in Subjects Infected With SARS-CoV-2 With or Without Symptoms | NCT04407507 | Investigacion Biomedica para el Desarrollo de Farmacos S.A. de C.V. | - | Interventional | 66 | - Ivermectin - Placebo | - Participants with a disease control status defined as no disease progression to severe | Not yet recruiting |
| 42 | Worldwide Trends on COVID-19 Research After the Declaration of COVID-19 Pandemic | NCT04460547 | Qassim University | - | Observational, Retrospective Cohort | 200 | - Convalescent Plasma Transfusion - Hydroxychloroquine - DAS181 - Ivermectin, - Interferon Beta-1A | - Geographical distribution of the interventional and observational studies - Monthly Research study completion rate as per geographic distribution of the Research - Statistical correlation of the interventional, Observational, drug based interventional, diagnostic test based interventional and device based interventional studies | Not yet recruiting |
