## Supplemental Figures 1-31 for "An Updated Systematic Review and Meta-Analysis of Mortality, Need for ICU admission, Use of Mechanical Ventilation, Adverse effects and other Clinical Outcomes of Ivermectin Treatment in COVID-19 Patients"

**e-figure 1 Sensitivity analysis excluding Lima-morales et al: Mortality in Clinical trials**

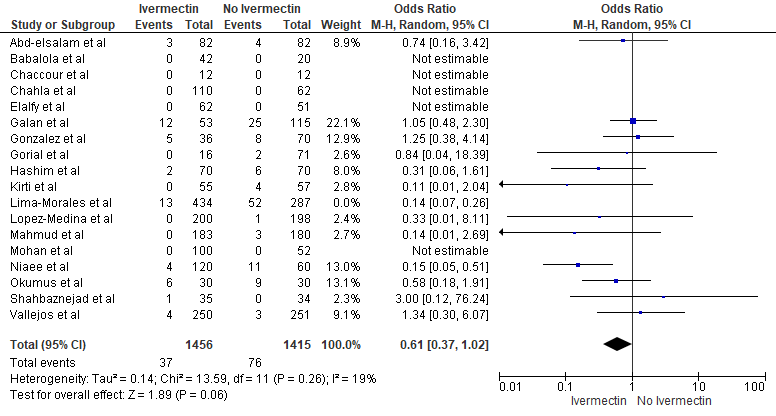

**e-figure 2 Sensitivity analysis excluding Vallejos et al: Mortality in Therapeutic RCTs: Mild-Moderate COVID-19**

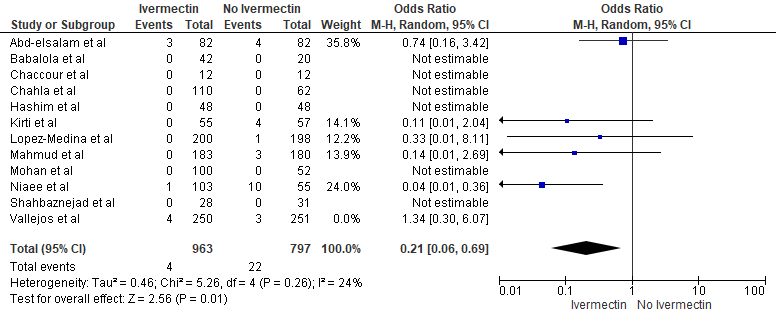

**e-figure 3 Sensitivity analysis excluding Soto-Becerra et al: Mortality in observational studies**

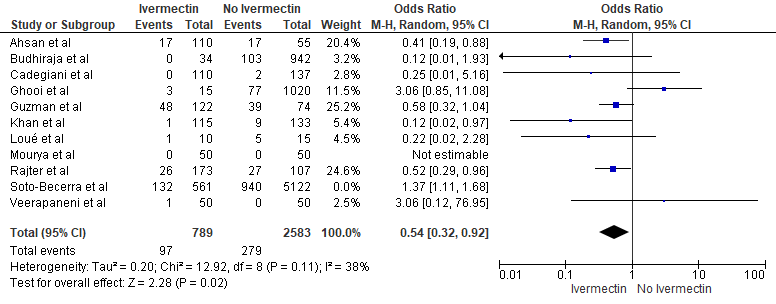

**e-figure 4 Sensitivity analysis excluding Soto-Becerra et al: Mortality in hospitalized patients-Overall**

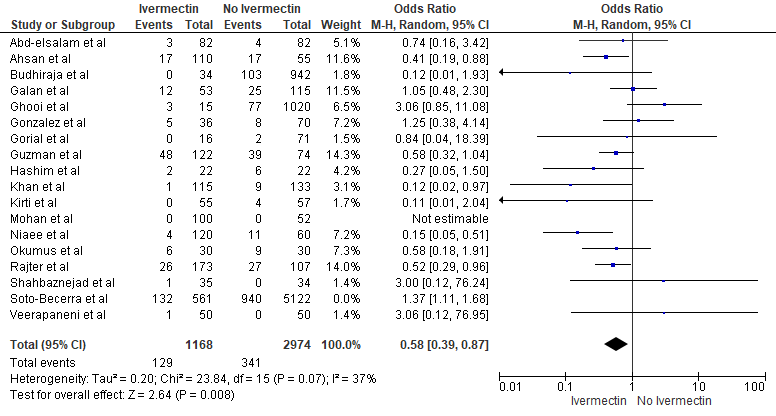

**e-figure 5 Sensitivity analysis excluding Abd-elsalam et al:** **Mortality in hospitalized patients by study design RCTs: Mild-Moderate COVID-19**

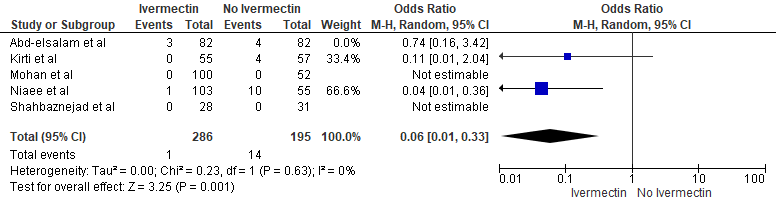

**e-figure 6 Sensitivity Analysis excluding Soto-Becerra et al: Mortality in inpatient observational studies**

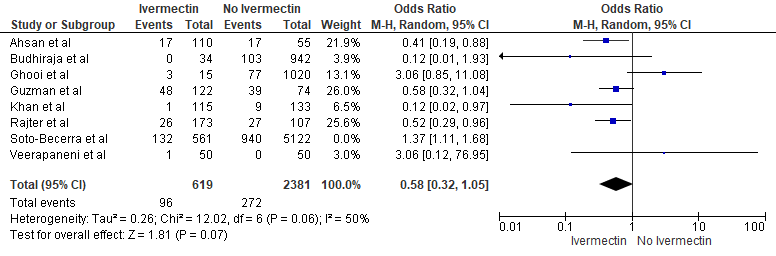

**e-figure 7 Pooled Analysis for Mortality**

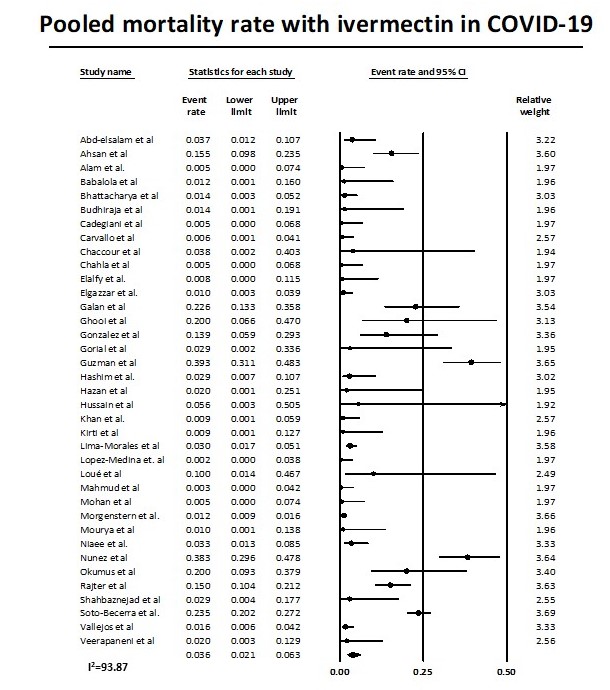

**e-figure 8 Pooled analysis of need for ICU admission**

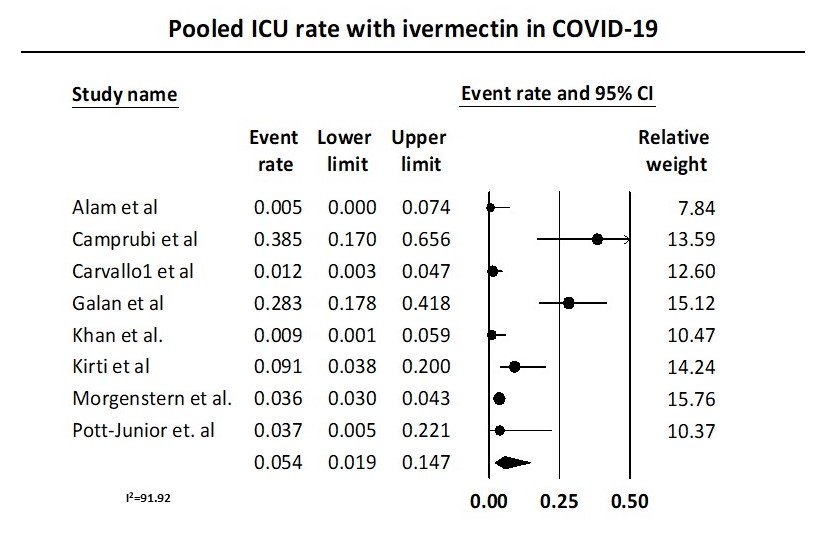

**e-figure 9 Pooled analysis of Mechanical Ventilation**

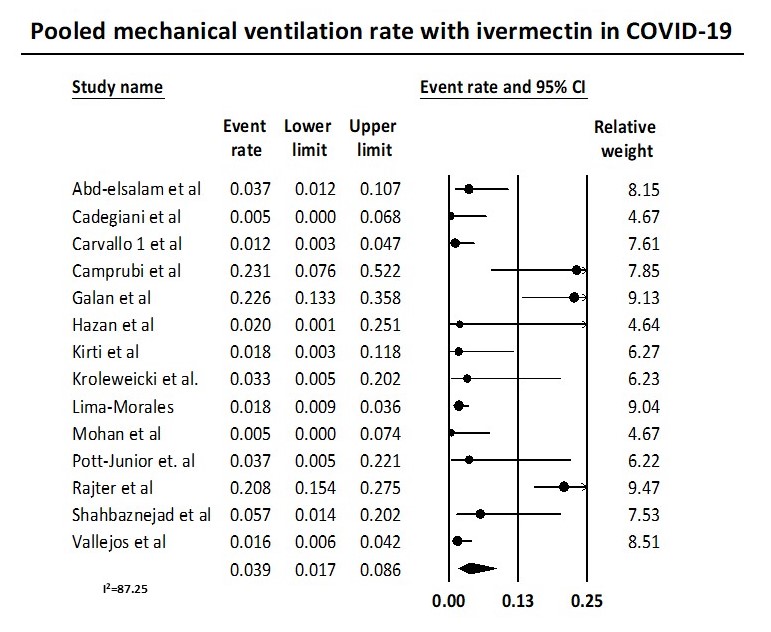

**e-figure 10 Pooled Analysis of Adverse events**

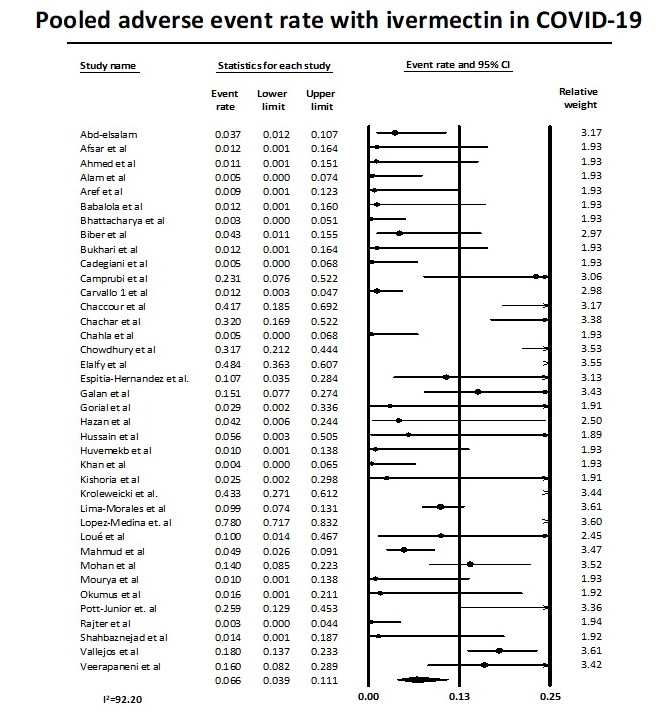

**e-figure 11 Sensitivity Analysis excluding Maurya et al : Viral Clearance**

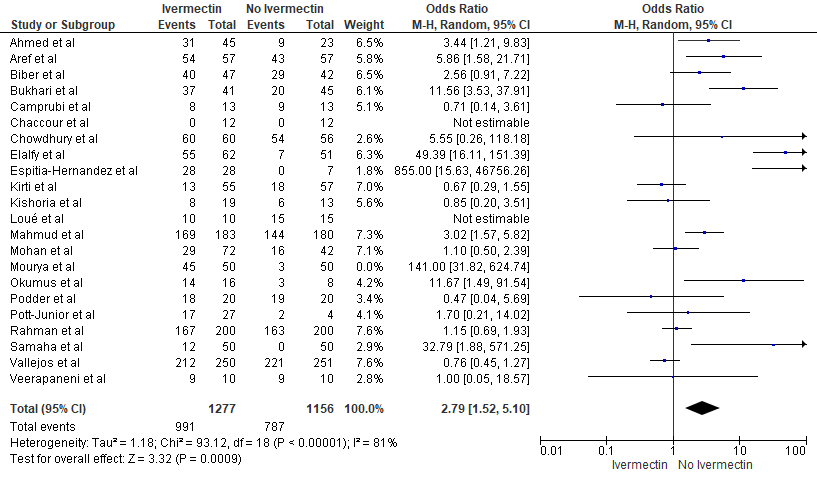

**e-figure 12 Sensitivity Analysis excluding Khan et al : Time to achieve Viral Clearance**

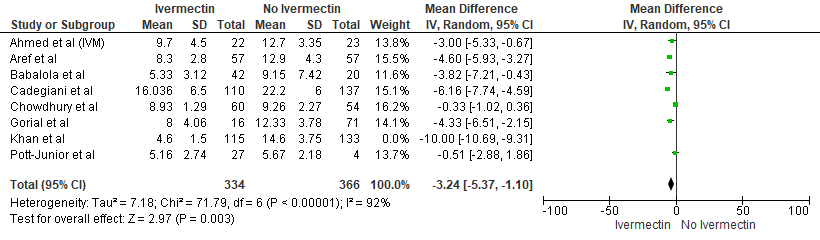

**e-figure 13 Sensitivity Analysis excluding Mahmud et al : Need for hospitalization**

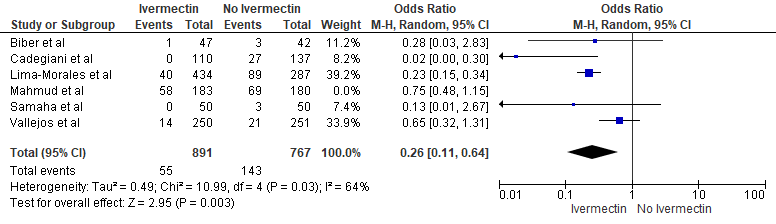

**e-figure 14 Sensitivity Analysis excluding** **Gonzalez et al : Duration of hospital stay**

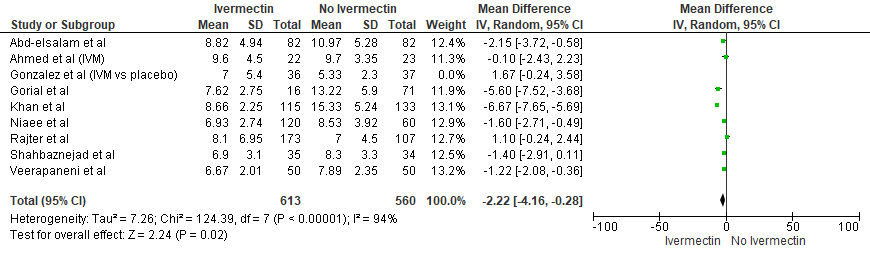

**e-figure 15 Funnel Plot overall mortality**

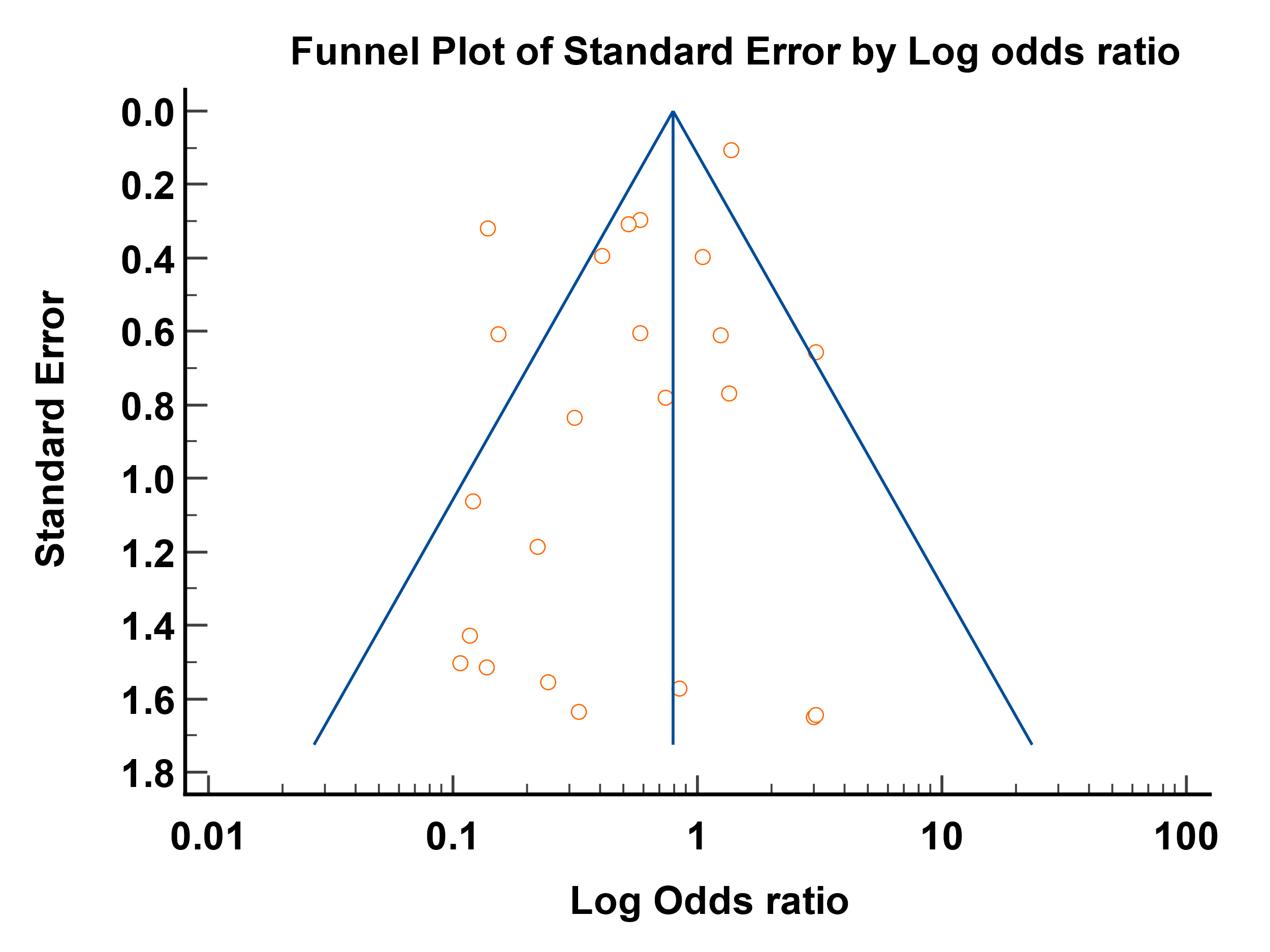

| Egger's test | |
| --- | --- |
| Intercept | -1.2540 |
| 95% CI | -2.3366 to -0.1714 |
| Significance level | P = 0.0253 |

**e-figure 16 Funnel Plot Mortality in Clinical Trials**

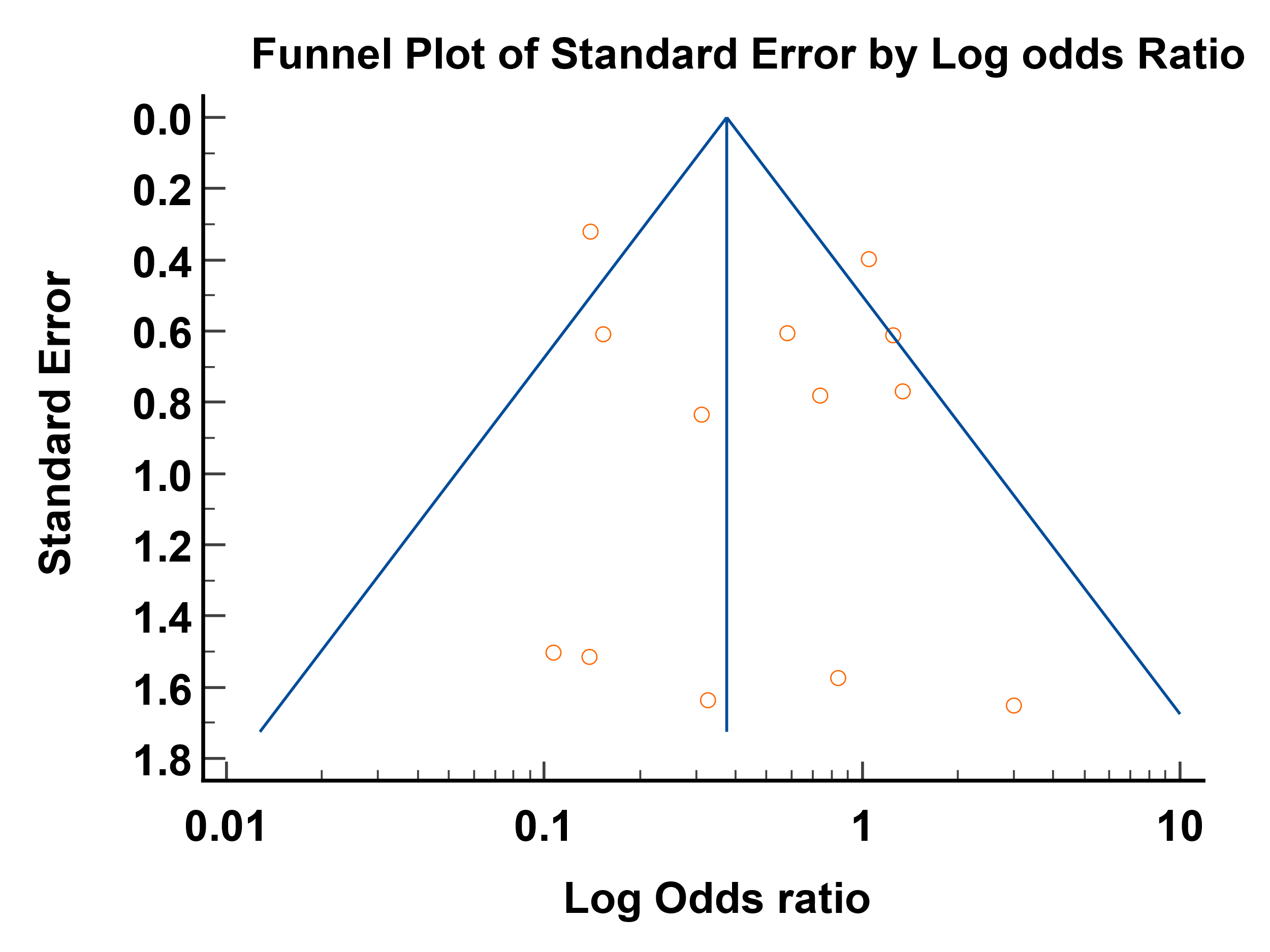

| Egger's test | |
| --- | --- |
| Intercept | 0.6962 |
| 95% CI | -1.2960 to 2.6884 |
| Significance level | P = 0.4580 |

**e-figure 17** **Funnel Plot Mortality in RCTs: Mild/Moderate COVID-19**

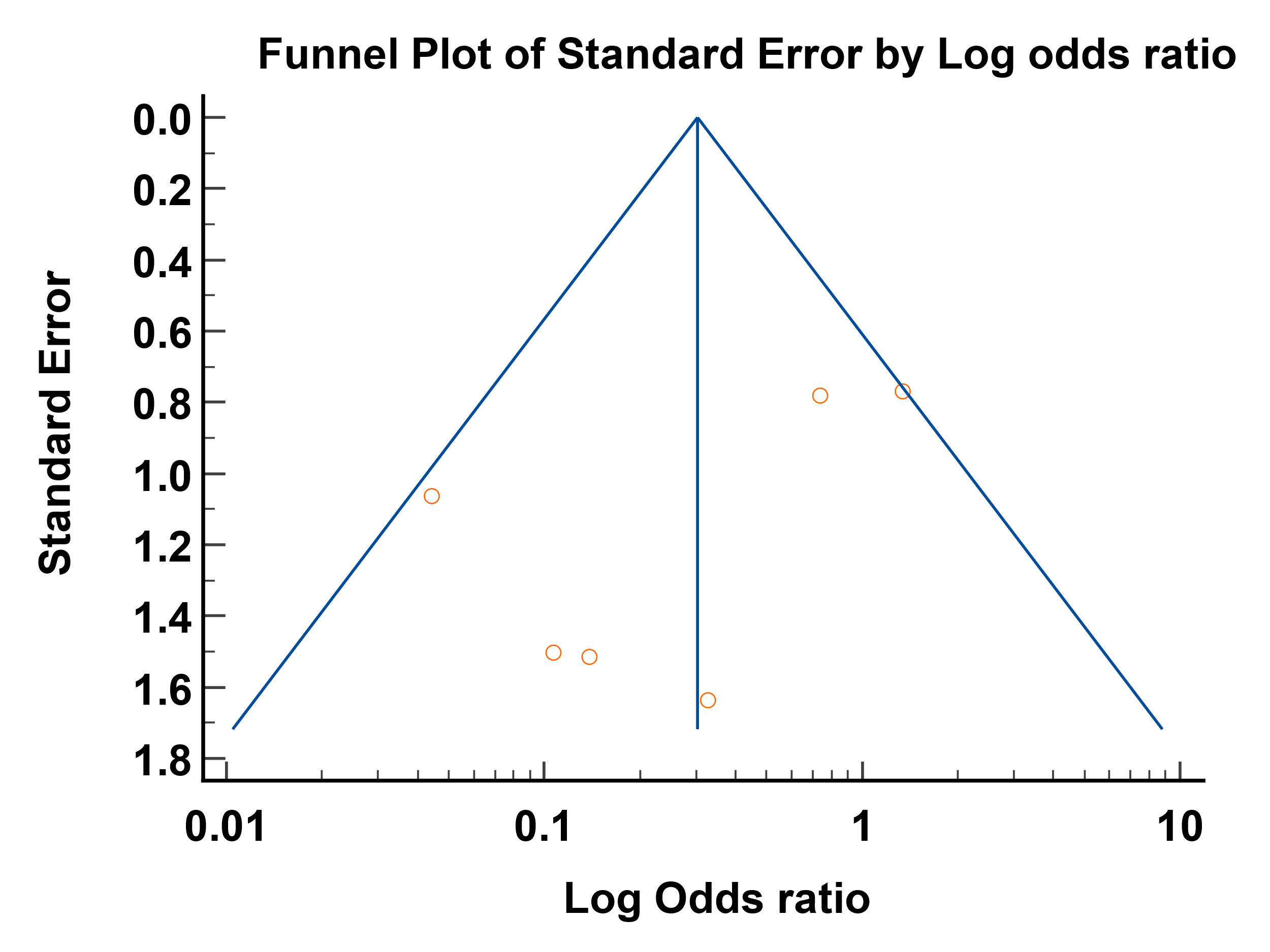

| Egger's test | |
| --- | --- |
| Intercept | -2.5348 |
| 95% CI | -6.7667 to 1.6972 |
| Significance level | P = 0.1716 |

**e-figure 18** **Funnel Plot Mortality in RCTs: Severe/Critical COVID-19**

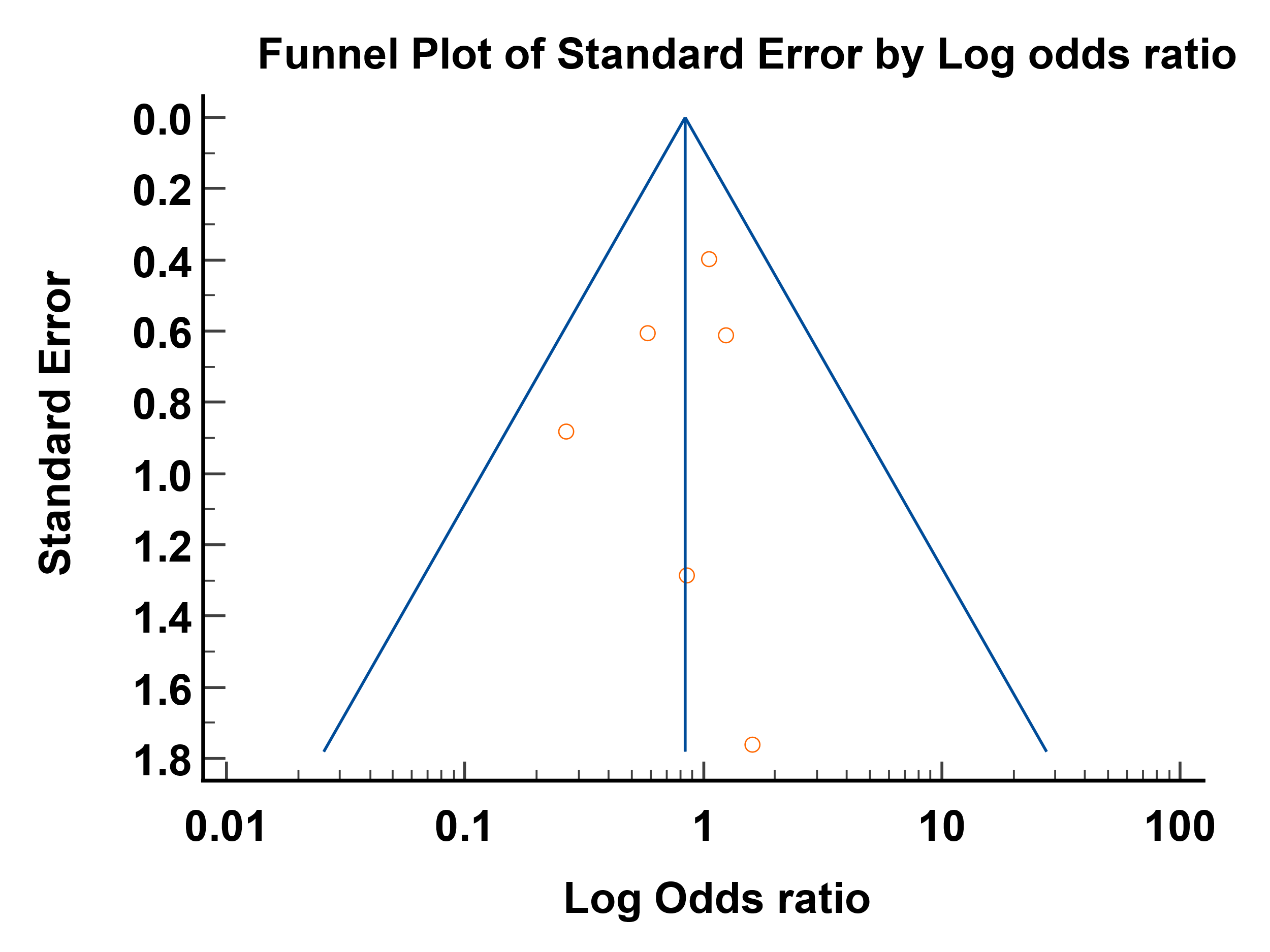

| Egger's test | |
| --- | --- |
| Intercept | -0.4114 |
| 95% CI | -2.6286 to 1.8058 |
| Significance level | P = 0.6336 |

**e-figure 19 Funnel Plot Mortality in Observational studies**

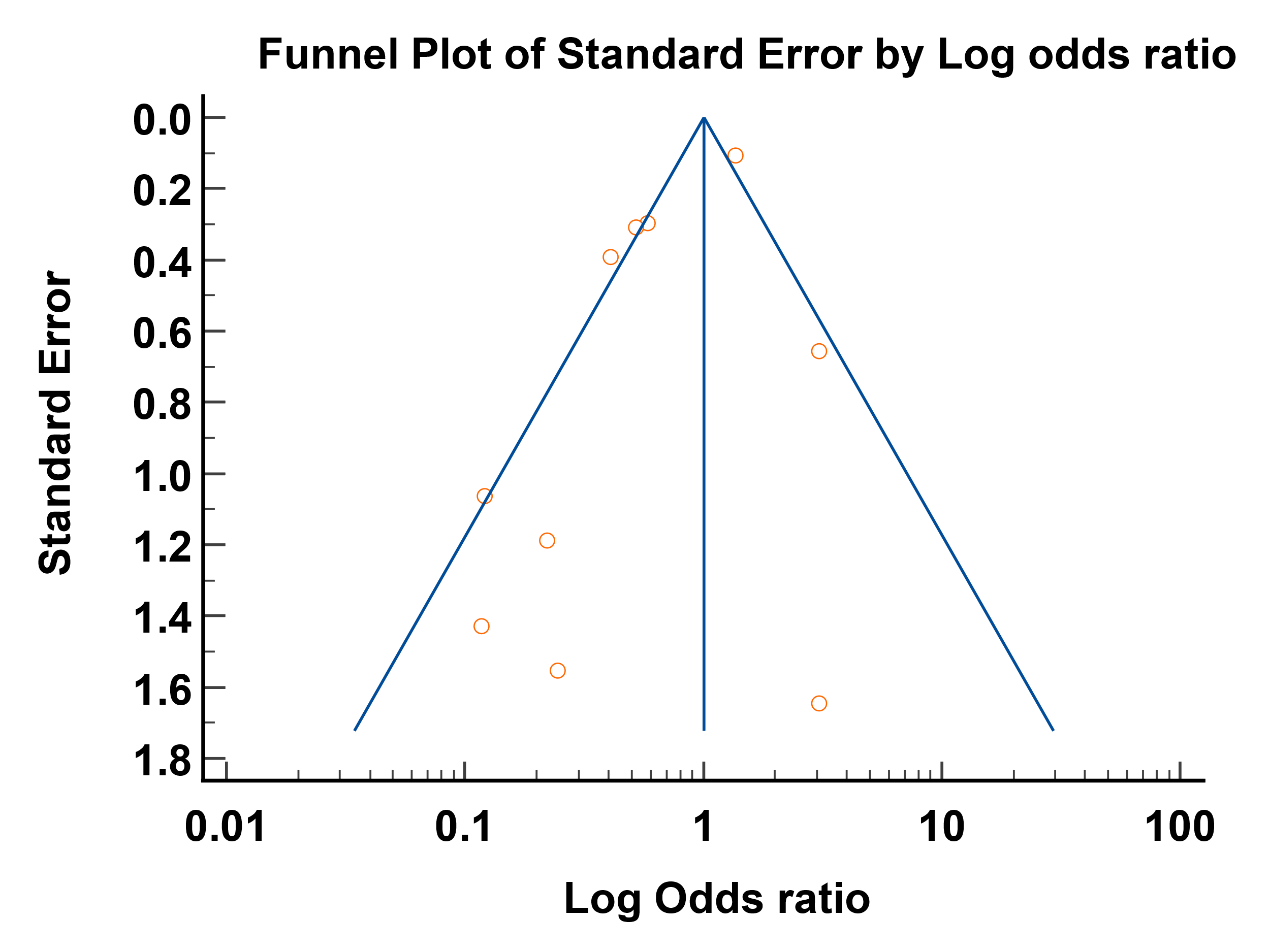

| Egger's test | |
| --- | --- |
| Intercept | -1.4928 |
| 95% CI | -3.1154 to 0.1297 |
| Significance level | P = 0.0667 |

**e-figure 20 Funnel Plot Mortality in Hospitalized patient overall**

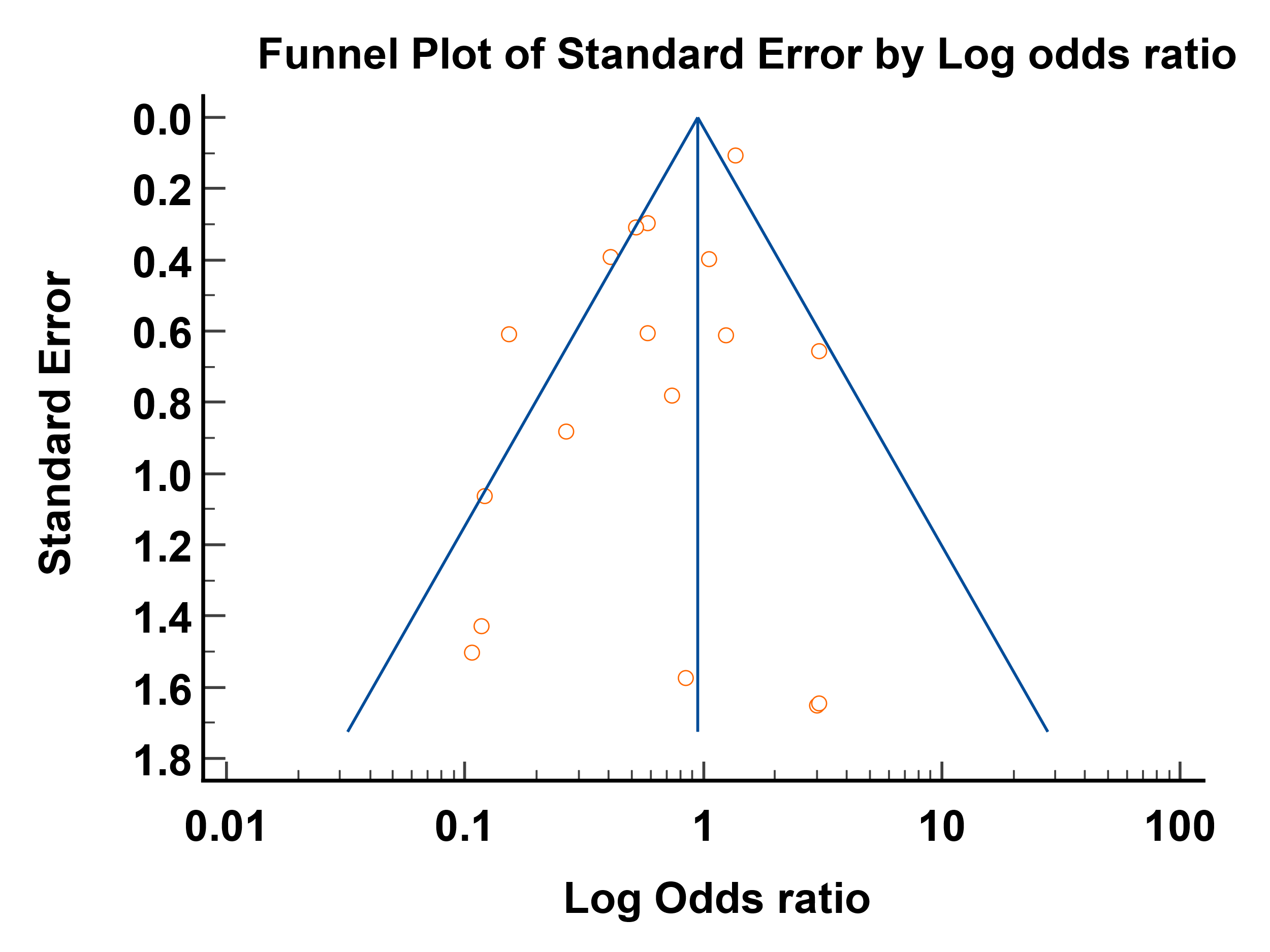

| Egger's test | |
| --- | --- |
| Intercept | -1.2484 |
| 95% CI | -2.3243 to -0.1726 |
| Significance level | P = 0.0258 |

**e-figure 21 Funnel Plot Mortality in Inpatient RCTs: Mild-Moderate COVID-19**
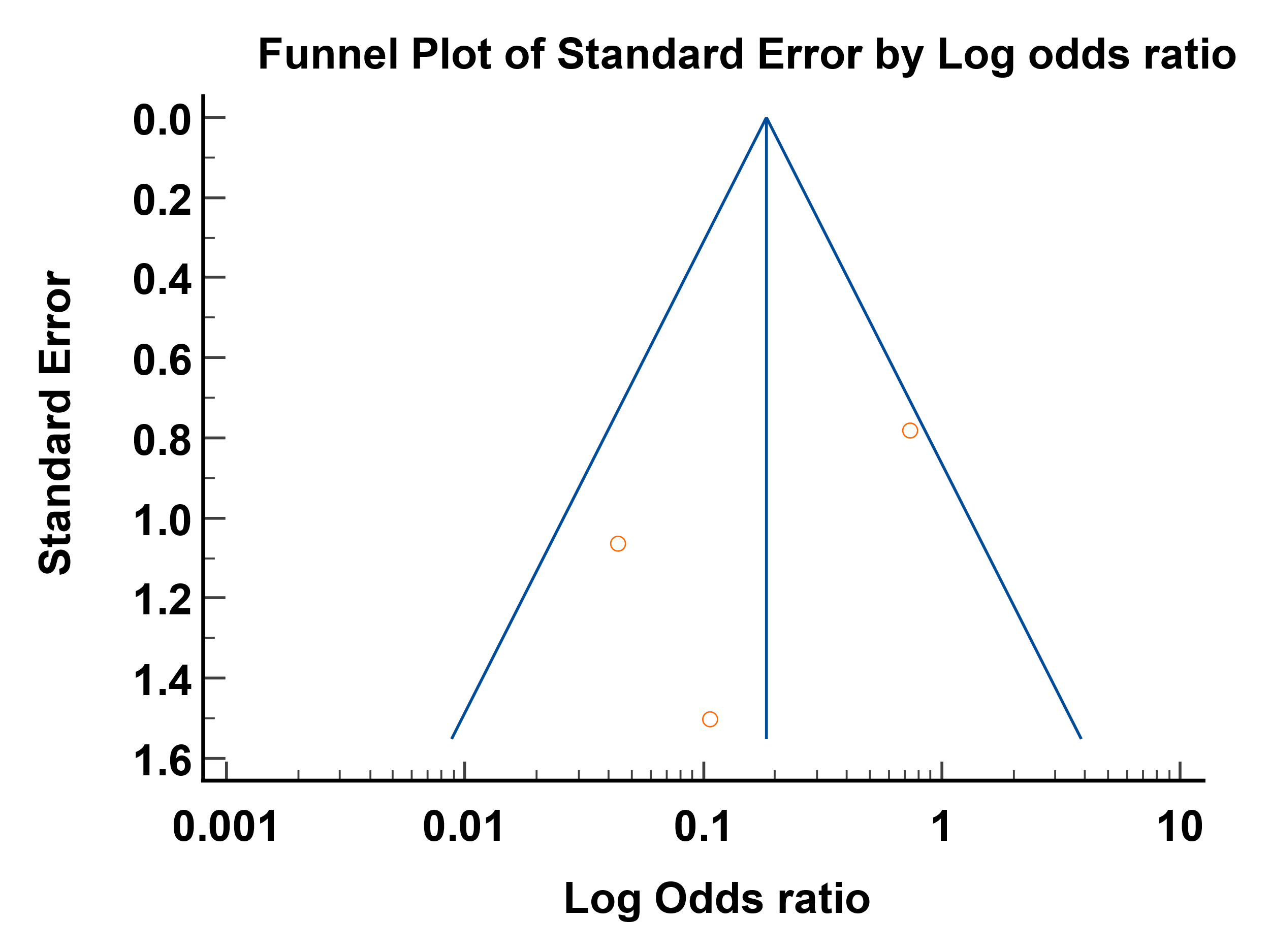

| Egger's test | |
| --- | --- |
| Intercept | -3.5403 |
| 95% CI | -49.2449 to 42.1643 |
| Significance level | P = 0.5051 |

**e-figure 22 Funnel Plot Mortality in Inpatient RCTs: Severe/Critical COVID-19**

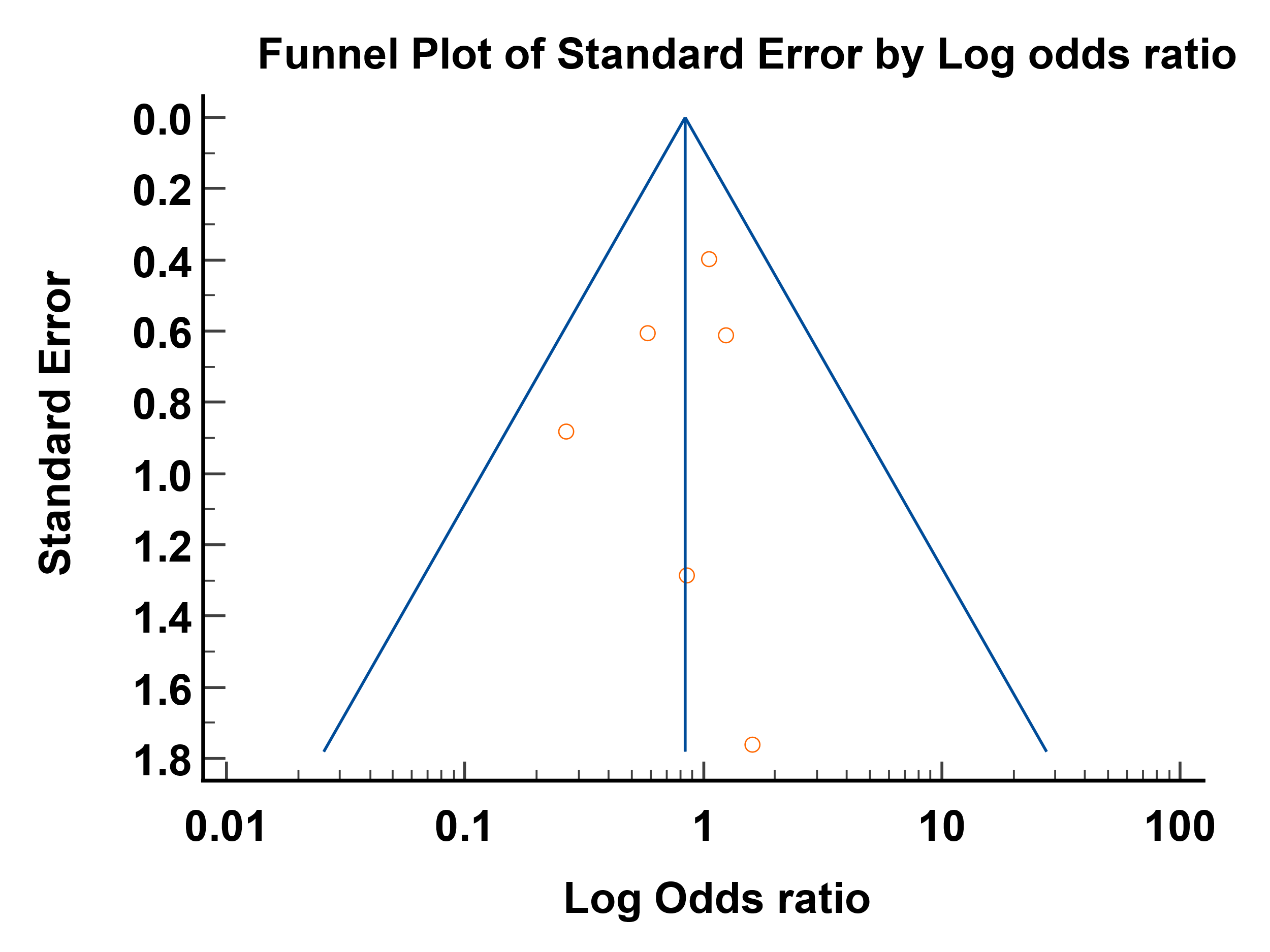

| Egger's test | |
| --- | --- |
| Intercept | -0.4114 |
| 95% CI | -2.6286 to 1.8058 |
| Significance level | P = 0.6336 |

**e-figure 23 Funnel Plot Mortality in Inpatient Observational studies**

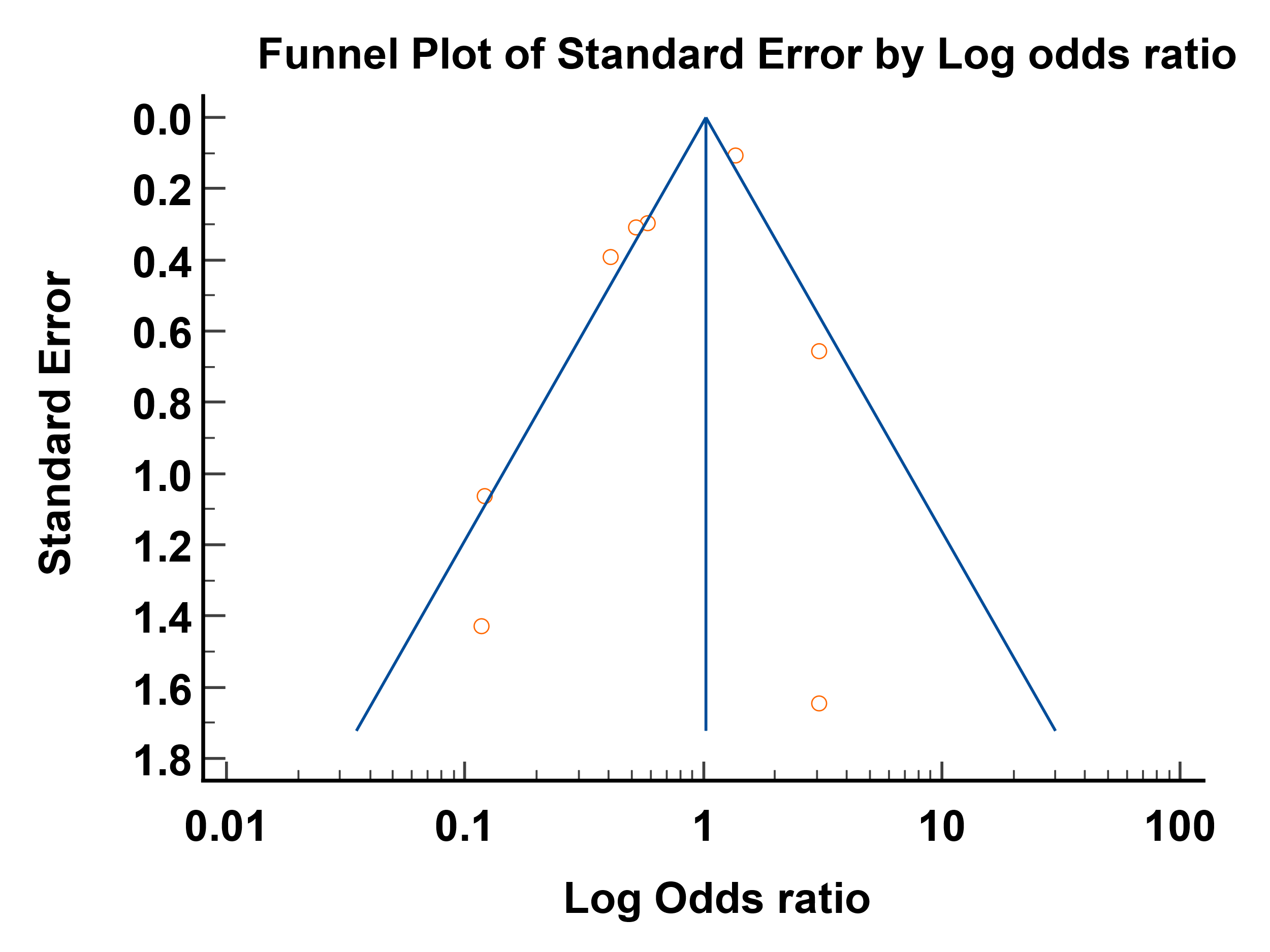

| Egger's test | |
| --- | --- |
| Intercept | -1.5595 |
| 95% CI | -3.8869 to 0.7680 |
| Significance level | P = 0.1522 |

**e-figure 24** **Funnel Plot for Mortality with Ivermectin Monotherapy as per Cochrane’s Criteria**

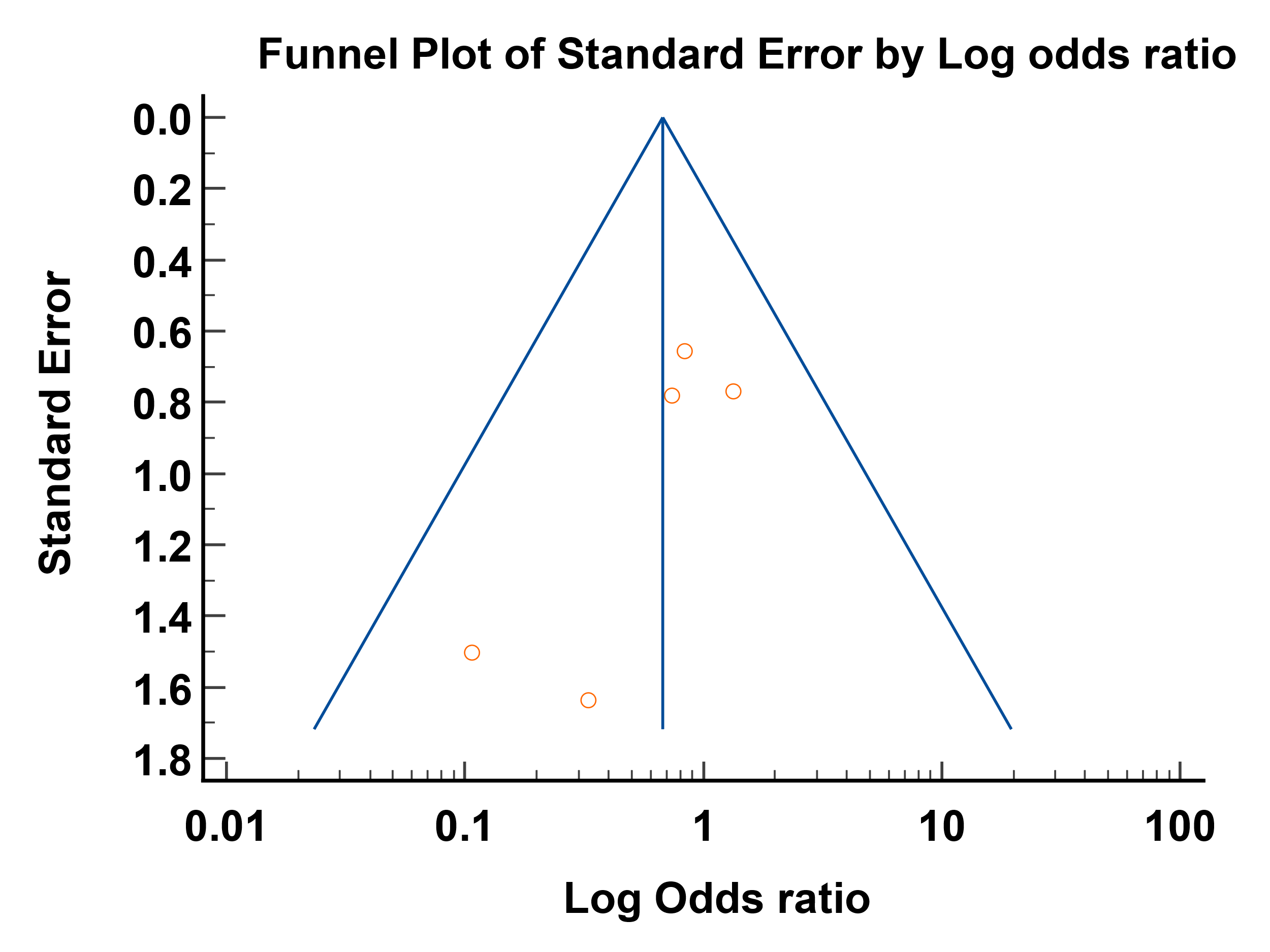

| Egger's test | |
| --- | --- |
| Intercept | -1.7775 |
| 95% CI | -4.1601 to 0.6050 |
| Significance level | P = 0.0981 |

**e-figure 25 Funnel Plot Need for ICU Admission**

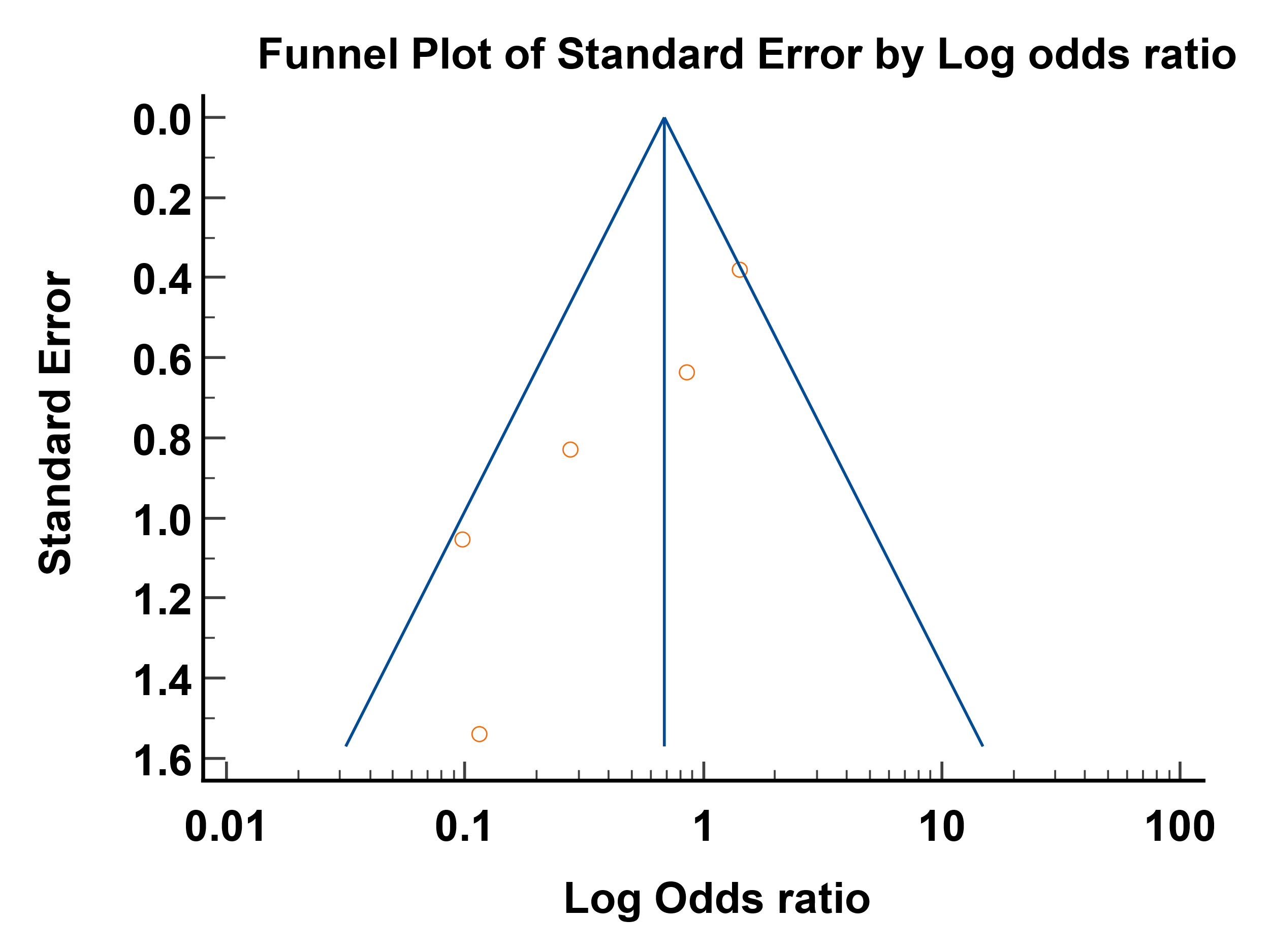

| Egger's test | |
| --- | --- |
| Intercept | -2.9550 |
| 95% CI | -4.8940 to -1.0161 |
| Significance level | P = 0.0167 |

**e-figure 26 Funnel Plot Need for Mechanical Ventilation**

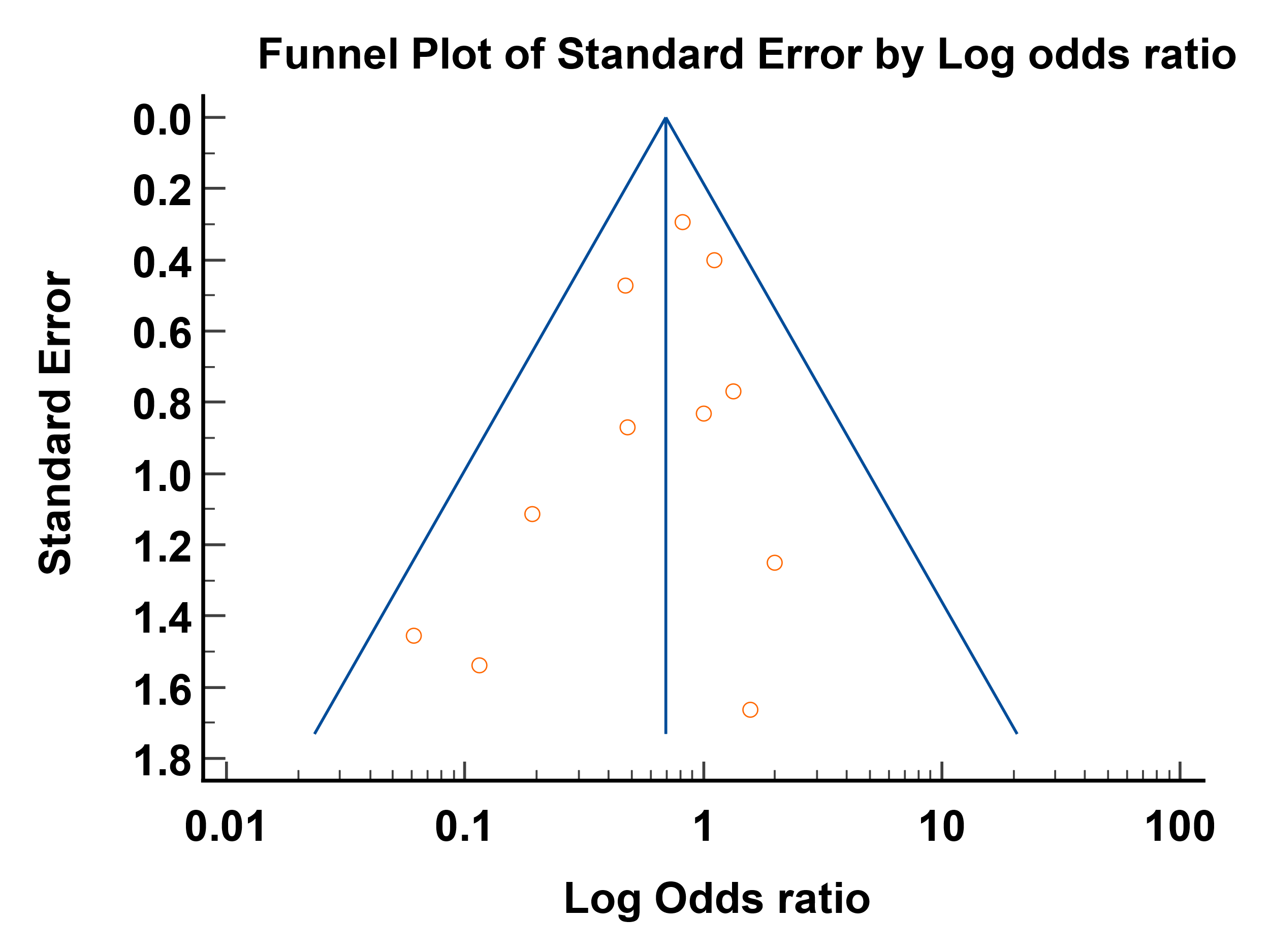

| Egger's test | |
| --- | --- |
| Intercept | -0.6703 |
| 95% CI | -1.9167 to 0.5761 |
| Significance level | P = 0.2547 |

**e-figure 27 Funnel Plot Adverse event**

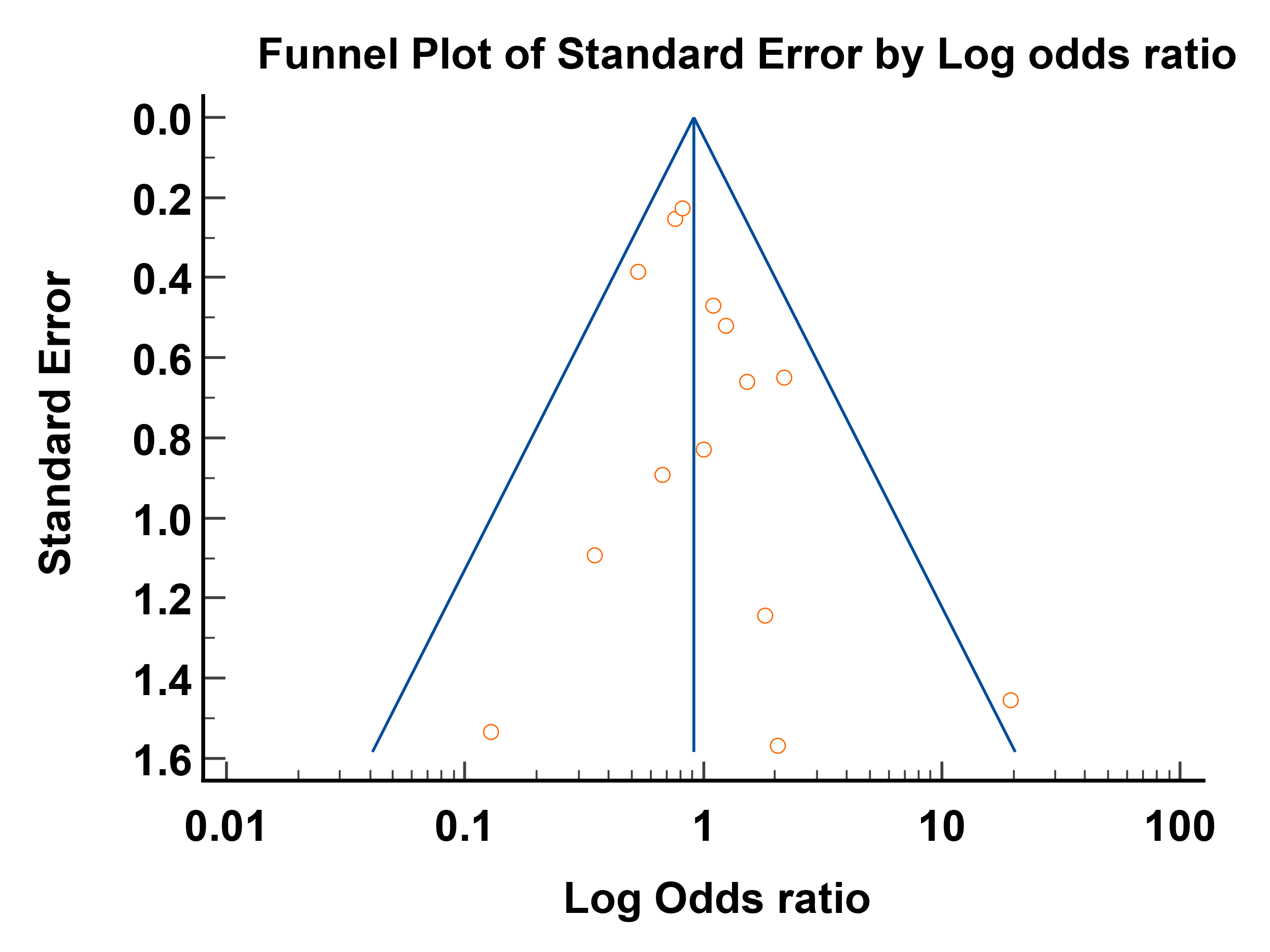

| Egger's test | |
| --- | --- |
| Intercept | 0.5750 |
| 95% CI | -0.4471 to 1.5972 |
| Significance level | P = 0.2438 |

**e-figure 28 Funnel Plot Need for Hospitalization**

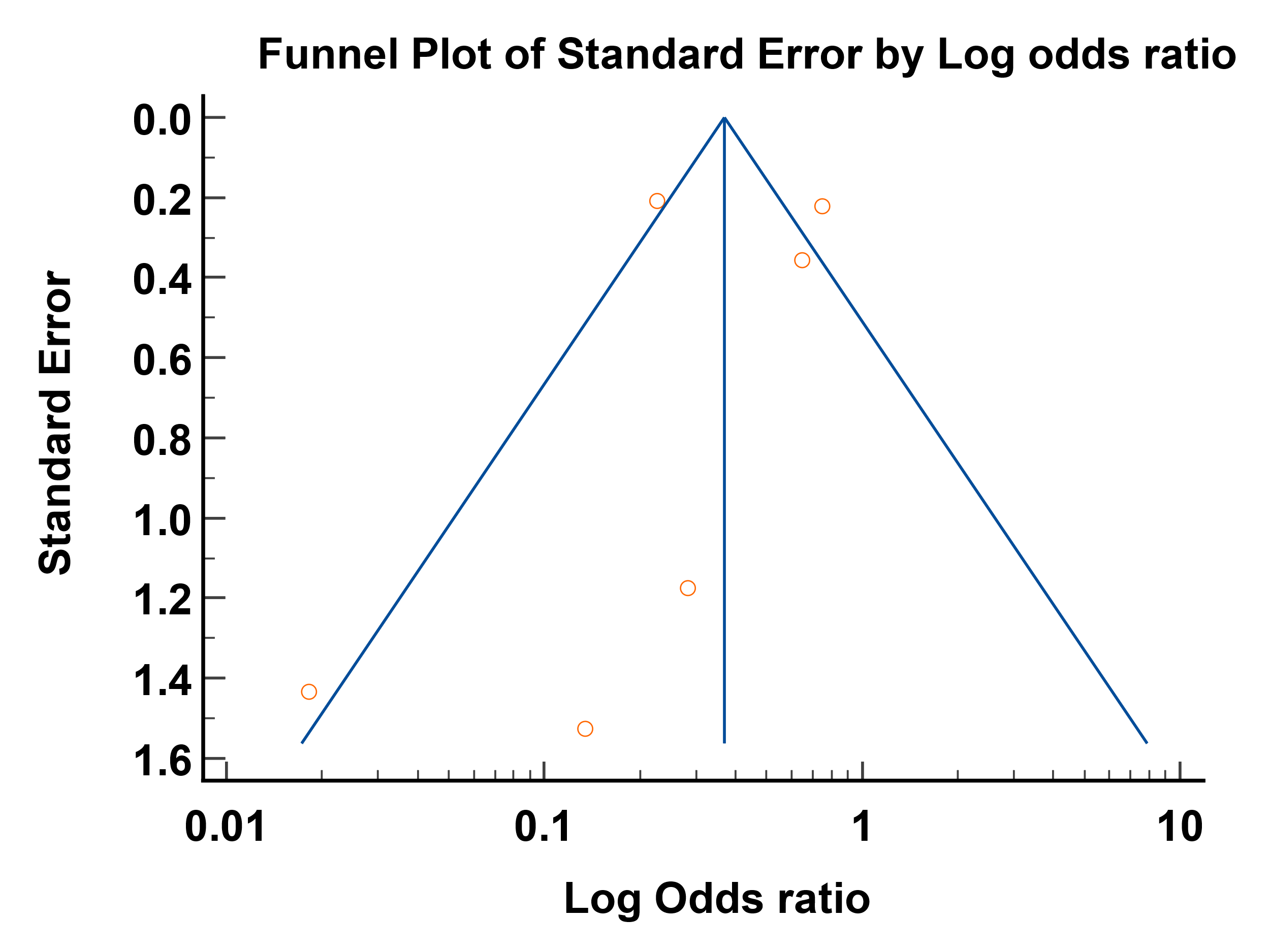

| Egger's test | |
| --- | --- |
| Intercept | -0.9777 |
| 95% CI | -5.2887 to 3.3333 |
| Significance level | P = 0.5631 |

**e-figure 29 Funnel Plot duration of Hospital stay**

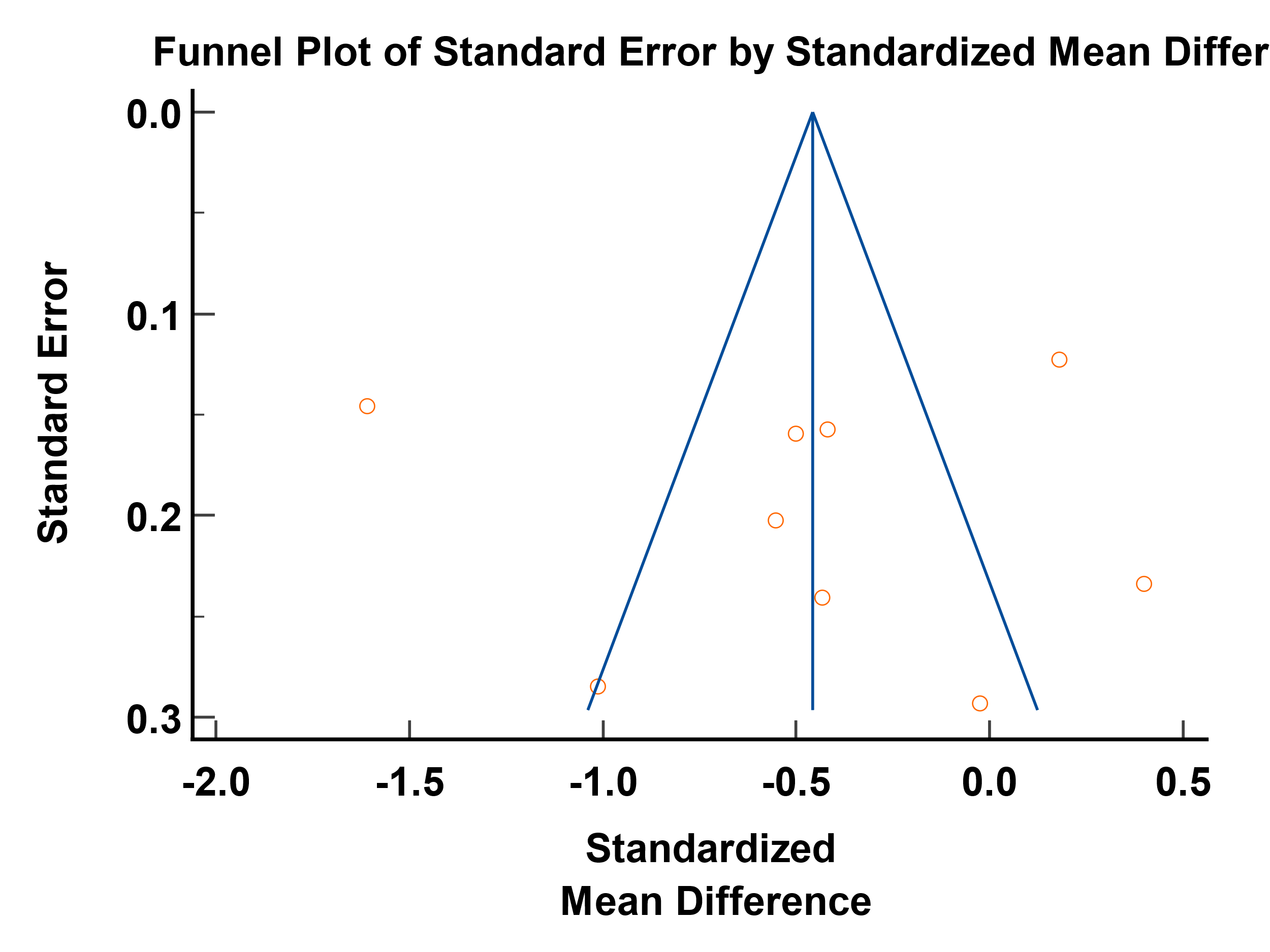

| Egger's test | |
| --- | --- |
| Intercept | 0.1327 |
| 95% CI | -10.9465 to 11.2119 |
| Significance level | P = 0.9782 |

**e-figure 30 Funnel Plot Incidence of Viral Clearance**

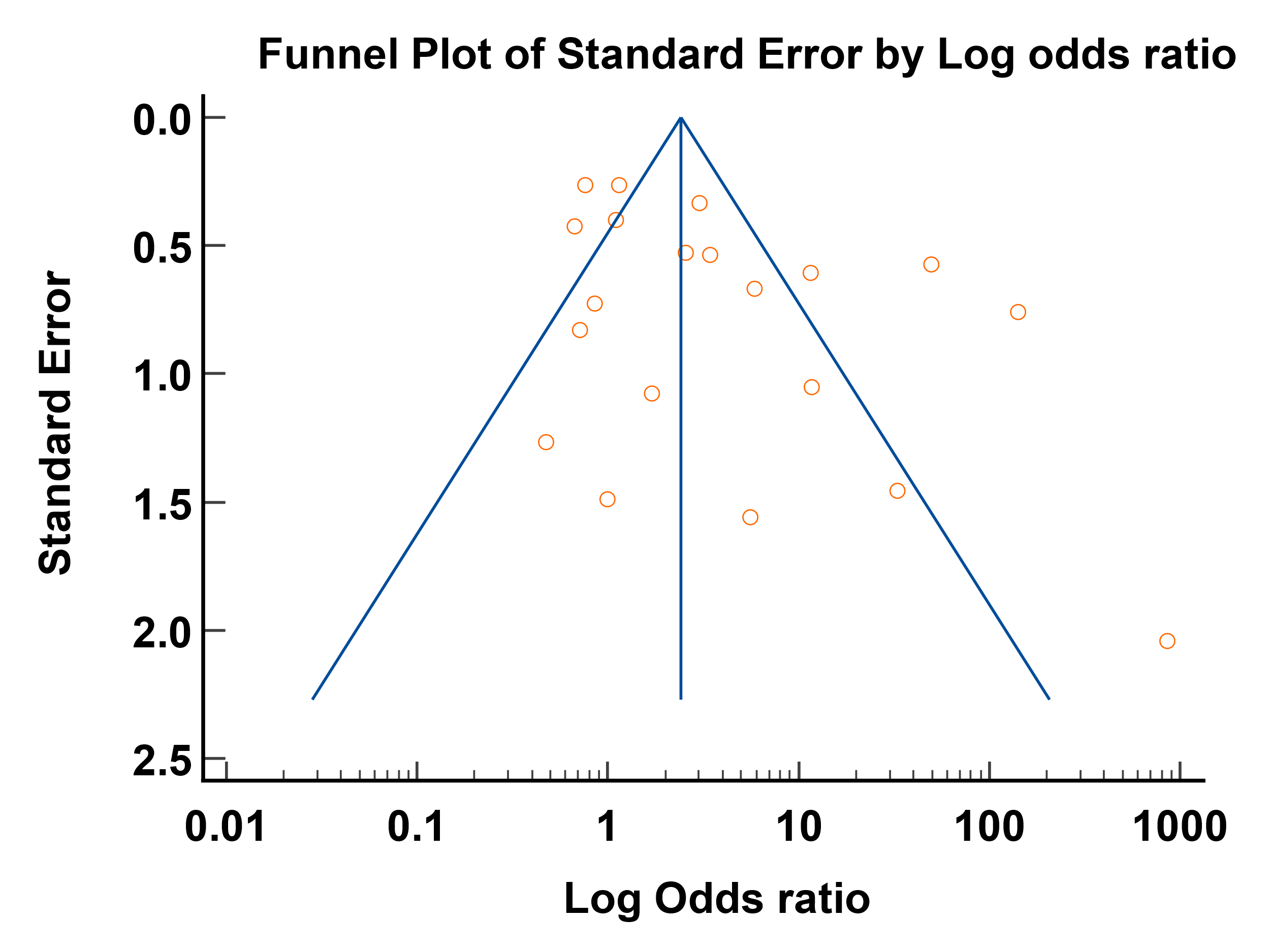

| Egger's test | |
| --- | --- |
| Intercept | 2.3404 |
| 95% CI | 0.1922 to 4.4886 |
| Significance level | P = 0.0344 |

**e-figure 31 Funnel Plot Time to achieve viral clearance**

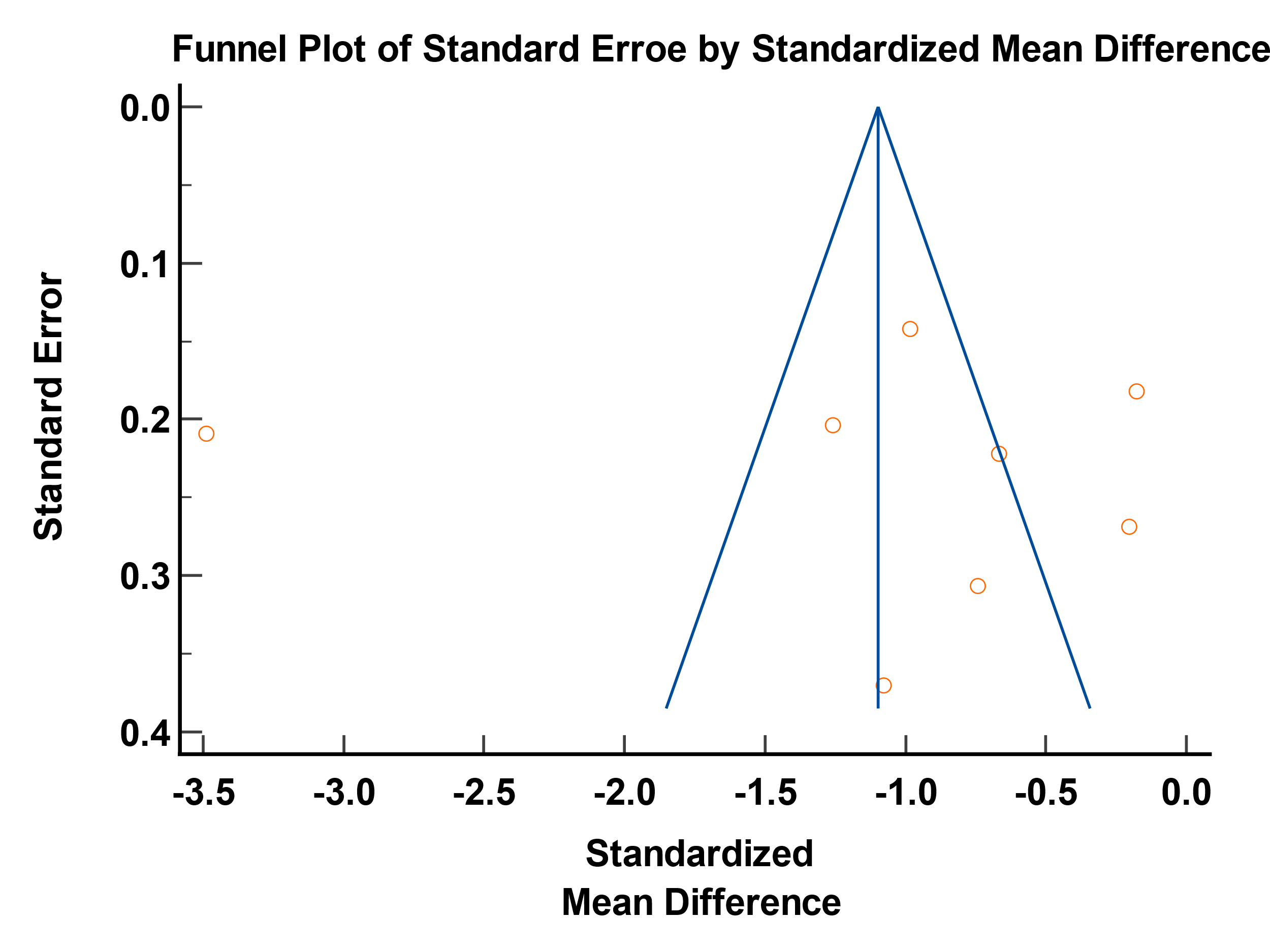

| Egger's test | |
| --- | --- |
| Intercept | 0.2107 |
| 95% CI | -17.0617 to 17.4831 |
| Significance level | P = 0.9772 |
